# The effect of school closures and reopening strategies on COVID-19 infection dynamics in the San Francisco Bay Area: a cross-sectional survey and modeling analysis

**DOI:** 10.1101/2020.08.06.20169797

**Authors:** Jennifer R. Head, Kristin L. Andrejko, Qu Cheng, Philip A. Collender, Sophie Phillips, Anna Boser, Alexandra K. Heaney, Christopher M. Hoover, Sean L. Wu, Graham R. Northrup, Karen Click, Robert Harrison, Joseph A. Lewnard, Justin V. Remais

## Abstract

**Background:** Large-scale school closures have been implemented worldwide to curb the spread of COVID-19. However, the impact of school closures and re-opening on epidemic dynamics remains unclear.

**Methods:** We simulated COVID-19 transmission dynamics using an individual-based stochastic model, incorporating social-contact data of school-aged children during shelter-in-place orders derived from Bay Area (California) household surveys. We simulated transmission under observed conditions and counterfactual intervention scenarios between March 17-June 1, and evaluated various fall 2020 K-12 reopening strategies.

**Findings:** Between March 17-June 1, assuming children <10 were half as susceptible to infection as older children and adults, we estimated school closures averted a similar number of infections (13,842 cases; 95% CI: 6,290, 23,040) as workplace closures (15,813; 95% CI: 9,963, 22,617) and social distancing measures (7,030; 95% CI: 3,118, 11,676). School closure effects were driven by high school and middle school closures. Under assumptions of moderate community transmission, we estimate that fall 2020 school reopenings will increase symptomatic illness among high school teachers (an additional 40.7% expected to experience symptomatic infection, 95% CI: 1.9, 61.1), middle school teachers (37.2%, 95% CI: 4.6, 58.1), and elementary school teachers (4.1%, 95% CI: −1.7, 12.0). Results are highly dependent on uncertain parameters, notably the relative susceptibility and infectiousness of children, and extent of community transmission amid re-opening. The school-based interventions needed to reduce the risk to fewer than an additional 1% of teachers infected varies by grade level. A hybrid-learning approach with halved class sizes of 10 students may be needed in high schools, while maintaining small cohorts of 20 students may be needed for elementary schools.

**Interpretation:** Multiple in-school intervention strategies and community transmission reductions, beyond the extent achieved to date, will be necessary to avoid undue excess risk associated with school reopening. Policymakers must urgently enact policies that curb community transmission and implement within-school control measures to simultaneously address the tandem health crises posed by COVID-19 and adverse child health and development consequences of long-term school closures.

**Funding:** JVR, JRH, QC, PAC, SP, AKH, CMH, and KC were supported in part by National Science Foundation grant no. 2032210, National Institutes of Health grants nos. R01AI125842, R01TW010286 and R01AI148336, and by the University of California Multicampus Research Programs and Initiatives award # 17-446315. JAL received support from the Berkeley Population Center (grant number P2CHD073964 from the National Institute of Child Health & Human Development, National Institutes of Health).

**Research in Context:** *Evidence before this study:* Given the urgent need to enact quick public health interventions to curb transmission of SARS-CoV-2, large-scale school closures were implemented globally. We searched the terms “school”, “children”, “closure”, “coronavirus”, and “COVID-19” in PubMed to assess the current evidence evaluating the role of school closures in mitigating SARS-CoV-2 transmission. Data motivating the decision to close schools remained largely limited to experiences with influenza outbreaks, where children are highly susceptible to infection, are key drivers of transmission, and experience severe outcomes. At the time of writing, no modeling studies to our knowledge have quantified the net impact of COVID-19 related school closures in the United States, and observational studies that documented decreases in COVID-19 incidence associated with statewide school closures are subject to confounding by other concurrently implemented non-pharmaceutical interventions. Further, the scientific consensus remains fragmented in its understanding of key epidemiological parameters, namely the relative susceptibility and infectiousness of children compared to adults, exacerbating uncertainties around the risks of opening schools. As policymakers weigh the negative consequences of school closures on child health and development against the risks of reopening, it becomes critical to discern the range of potential impacts of school reopenings on the COVID-19 epidemic accounting for uncertainty in epidemiological parameters and plausible strategies for risk mitigation.

*Added value of this study:* This study uses an individual-based transmission model parameterized with contact patterns we derived from a web-based contact survey administered to Bay Area (California) households with children during school closures to advance the understanding of the relative impact of Bay Area spring 2020 school closures compared to other non-pharmaceutical interventions, and projects the potential impact of school reopening strategies in the fall 2020 semester. Within the context of our model, we found that school closures averted a similar number of cases as workplace closures in spring 2020, with most of the averted cases attributable to high school closures. We found that COVID-19 risks associated with reopening schools in fall 2020 are highly dependent on the relative susceptibility of children and the level of community transmission at the time of reopening. Strategies necessary to reduce school transmission such that fewer than an additional 1% of teachers would be infected varied across school divisions. Safely reopening high schools may require combining multiple strict contact reduction measures, including staggering school days, halving class sizes, or maintaining small, stable cohorts, while safely reopening elementary schools may be achieved with a more limited set of interventions, including use of stable cohorts and masks.

*Implications of all the available evidence:* Under plausible assumptions regarding the susceptibility and infectiousness of school-aged children and teenagers, this study highlights heterogeneity of COVID-19 risks, and necessary mitigation strategies, associated with reopening across levels of schooling. It also highlights the urgency of resolving uncertain parameters, especially those pertaining to the relative susceptibility and infectiousness of children. Research is needed to quantify the role of children in transmission of COVID-19 in schools or similar settings to enumerate the risk of school-based outbreaks, particularly as transmission remains high in many regions of the United States. To balance both the adverse long-term consequence of school closures on child development and concerns about safe reopening, policy makers must quickly devote resources to ensure schools that choose to reopen amid uncertain evidence can adopt and adhere to strict infection, prevention, and control strategies that are critical to ensuring students, teachers, and community members remain healthy.

## Background

Amid the lack of effective therapeutics or vaccination, large-scale school closures have been instituted globally to reduce transmission of the Severe Acute Respiratory Syndrome Coronavirus 2 (SARS-CoV-2).^1^ Evidence motivating the decision to close schools came primarily from experience with interventions to reduce the transmission of other respiratory pathogens like influenza^2^, where children are key drivers of transmission and highly susceptible to infection and severe outcomes.^3^ School closures present a grave threat to healthy child development^4-6^ and may exacerbate existing racial and socioeconomic gaps in school achievement^7^, or nutrition.^8^ As such, there is an urgent need to assess the impact of school closures on SARS-CoV-2 transmission to date and weigh risks of school reopening amid ongoing SARS-CoV-2 transmission.^9^

To date, children constitute a small fraction of total COVID-19 cases in high-income countries, with estimates from the United States and China suggesting that children under 18 account for less than 2% of cases.^10,11^ Estimates of pediatric susceptibility to infection vary^12^, ranging from less than half as susceptible as adults^13-16^ even after accounting for biases^12^, to equally susceptible.^17,18^ While available data are limited to periods of prolonged school closures where social contact patterns were abnormal^19^, children have tended to be infected by household members in prospective infection studies^20,21^, and infections in children manifest largely as asymptomatic or with mild disease.^22^ It remains unclear to what extent pediatric symptomatic or asymptomatic infections contribute to community transmission^23,24^, though children and adults have been found to shed similar viral loads^25,26^, and contact tracing data from Israel, India, Italy, and South Korea has suggested that children over 10 years of age may be as infectious as adults.^16,17,27,28^

Early evidence from empirical and modeling data is varied with respect to the effectiveness of COVID-19 related school closures. A rapid systematic review found school closures in Asia did not contribute substantially to control^29^, with minimal community transmission in Taiwan even as schools remained open.^28^ Modeling work from the United Kingdom found school closures may avert fewer deaths than other non-pharmaceutical interventions^30^, leading some European countries to cautiously reopen schools while delaying the reopening of other sectors of the economy.^31,32^ Studies conducted before school shutdowns in France found limited evidence of secondary transmission within primary schools^33^, but antibody testing revealed the virus had spread within certain high schools to 38% of students and 43% of teachers.^33^ While some countries that significantly reduced community transmission have attempted to reopen schools, outbreaks in schools, camps, and daycares have persisted and caused reactive closures across various settings^34^, including an outbreak at a middle and high school in Israel where almost 200 students and staff were infected.^35^ Reports of COVID-related deaths among teachers in Sweden, where modifications to reduce class size and enhance social distancing were not made, highlight the urgency in enumerating the risks of reopening strategies.^35^

The objectives of this study were to: 1) estimate social contact patterns among school-aged children during Bay Area (California) COVID-19 related school closures; 2) estimate the cumulative incidence of COVID-19 throughout the 2019-2020 spring semester under counterfactual scenarios had schools or workplaces remained open, or social distancing policies not been enacted; and 3) estimate the effect of various school reopening strategies in Bay Area schools for the 2020 fall semester.

## Methods

We conducted a survey of families with school-aged children to ascertain the contact rates of children and their adult family members during the start of the Bay Area shelter-in-place. We used these contact rates within an individual-based transmission model to examine the impact of spring school closures and fall reopening strategies.

### Survey methodology

We designed and fielded a survey on social contacts of school-aged children in nine Bay Area (California) counties (Alameda, Contra Costa, Marin, Napa, San Francisco, San Mateo, Santa Clara, Solano, Sonoma) during county-wide shelter-in-place orders. Survey respondents were asked to report the number and location of non-household contacts made within six age categories (0-4, 5-12, 13-17, 18-35, 36-64, and 65+ years) throughout the day prior to survey completion. A contact was defined as an interaction within six feet with a non-household member lasting over five seconds.

A first sample was obtained using a web-based contact diary distributed in English via social networks (Nextdoor, Berkeley Parents Network) via a survey conducted between May 4 and June 1, 2020 of households containing at least one school-aged child (preK - grade 12). A second sample was procured between May 18 and June 1, 2020 via an online panel provider (Qualtrics) to be representative of the Bay Area by race/ethnicity and income. In both samples, surveys asked one respondent from each sampled household to respond on their own behalf and for all school-aged children in their household. The survey recorded household demographic information as well as the number and location of non-household contacts throughout the day prior to survey completion. A copy of the survey tool is included in the Supporting Information.

### Survey analysis

To adjust for potential selection bias, we calculated post-stratification weights reflecting joint distributions of race/ethnicity and income of the source population using the 2018 one-year American Community Survey Public Use Microdata Sample (PUMS) from the nine Bay Area counties. To account for potential bias due to occasional missing data on the location where contacts occurred, we applied a second set of weights equal to the inverse of the probability that an individual indicated the location of contacts, conditional on race and income (fixed effect) and household ID (random effect). Weighted and unweighted survey data yielded similar results (Supporting Information; Figure S2).

To determine whether the mean number of contacts differed across demographic strata, contact matrices generated using weighted and unweighted survey data were stratified by income, race, and location of contacts. To determine whether an individual’s total reported contacts varied by key covariates, we fitted a linear regression model accounting for random effects at the household level and fixed effects for age, race, household income, number of household members, single parent household, weekday of reported contact, school type, and a binary indicator of whether more adults work at home during shelter-in-place compared to prior to shelter-in-place.

We conducted all statistical analyses using R (version 3.2.2; R Foundation for Statistical Computing; Vienna, Austria), and fit random effects models using the *lme4* package.^36^

### Ethics statement

Ethical approval was obtained from the Institutional Review Board at the University of California, Berkeley (Protocol Number: 2020-04-13180). Prior to taking the anonymous survey, parents were provided a description of the survey and were asked to provide written informed consent.

### Transmission model

Using survey-derived estimates of contact patterns, we developed a transmission model to estimate the number of cases, hospitalizations, and deaths that would have occurred under various counterfactual situations, such as if schools had remained open, and used this model to simulate the impact of various school reopening strategies in the fall.

First, we generated 1,000 synthetic populations representative of the demographic composition of Oakland, California, following previous methods (see Supporting Information).^2^ Each individual was assigned an age, household, and occupation status (student, teacher, school staff, other employment, not employed) upon which membership in a class or workplace was based. Each individual represented 25 individuals in the real population. All possible pairings of individuals were partitioned into one of six types of interactions, according to a hierarchy of highest shared membership: household > classroom or workplace > grade > school > community.^37^ We defined elementary schools as grades K-5, middle schools as grades 6-8 and high schools as grades 9-12.

We then developed a discrete-time, age-structured individual-based stochastic model to simulate COVID-19 transmission dynamics in the synthetic population (Figure 1A). At each time increment, representative of one day, each individual is associated with an epidemiological state: susceptible (S), exposed (E), asymptomatic (A), symptomatic with non-severe illness (C), symptomatic with severe illness (H1, D1) resulting in eventual hospitalization before recovery (H2) or hospitalization before death (D2), recovered (R), or dead (M). A full description of the transmission model methodology is provided in the Supporting Information.

Based on their type of interaction (e.g. household, class, community), the daily contact rate between individuals *i* and *j* on day *t, K_ij,t_*, was estimated for pairs of individuals following previous study.^37^ Contact rates were scaled by a time-dependent factor between 0 (complete closure) and 1 (no intervention) representing a social distancing intervention to reduce contact between individual pairs. Pairs with a school or workplace interaction were reassigned as community interactions under closures. Because symptomatic individuals mix less with the community^38^, we simulated a 100% reduction in daily school or work contacts and a 75% reduction in community contacts for a proportion of symptomatic individuals, and an additional proportion of their household members.^30^ We assumed that individuals were in the infectious class for up to three days prior to observing symptoms^39^, during which time they did not reduce their daily contacts.

Transmission was implemented probabilistically for contacts between susceptible (S) and infectious individuals in the asymptomatic (A) or symptomatic and non-hospitalized states (C, H1, D1). Movement of individual *i* on day *t* from a susceptible to exposed class is determined by a Bernoulli random draw with probability of success given by the force of infection, *λ_i,t_*:

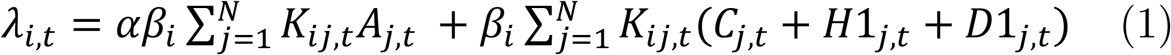

where *N* is the number of individuals in the synthetic population (N=16,000), *α* is the ratio of the force of infection between asymptomatic and symptomatic individuals, and *β_i_* is calculated from 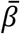, the population mean transmission rate of the pathogen. 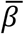 is determined using the next-generation matrix method.^40^ We represent age-varying susceptibility^13^ using an age-stratified *β_i_* that incorporates varying relative susceptibility by age (Supporting Information). Due to uncertainty in the relative susceptibility of children to SARS-CoV-2 infection compared with adults, we model scenarios where children under 10 years are half as susceptible as older children and adults, children under 20 years are half as susceptible as adults, and all individuals are equally as susceptible (see the Supporting Information for justification of this choice). Following previous work^13,41^, we assumed *α* to be less than one, as asymptomatic individuals may be less likely to spread infectious droplets by sneezing or coughing.^24^ Using this method, we calculated the secondary attack rate among household members to be between 9.6% and 11.1%, in agreement with prior studies.^15,17,18,27^

The duration of the latent period, d_L_, for each individual transitioning from the exposed class was drawn from a Weibull distribution with mean 5.4 days (95% CI: 2.4, 8.3).^42-44^ Whether an individual remained asymptomatic, or was hospitalized, or died was determined via Bernoulli random draws from age-stratified conditional probabilities (Figure 1B, Table 1). The time to recovery for non-hospitalized cases (mean: 13.1 days, 95% CI: 8.3, 16.9)^45^, the time to hospitalization for severe cases (mean: 10.3, 95% CI: 6.5, 13.3)^46^, and time to recovery or death for hospitalized cases (mean: 14.4, 95% CI: 11. 3, 16.6) were sampled from Weibull distributions (Table 1).^47^ Simulations were initiated on January 17, two weeks before the first known case in Santa Clara County, assuming a fully susceptible population seeded with a random number (range: 5-10) of exposed individuals.^48^ We averaged results over 1,000 independent realizations, using one random draw from the synthetic population, and estimated confidence intervals as the 2.5^th^ and 97.5^th^ percentile of all realizations.

**Table 1.** Parameters of the susceptible-exposed-infected-recovered model

| Parameter | Ages (i) | Values | References |
| --- | --- | --- | --- |
| Basic reproduction number, $R_0$ | all | 2.5 | Kucharski, et al<sup>55</sup><br>Wu, et al<sup>56</sup> |
| Average incubation period, $d_L$ (95% CI) | all | 5.4 (2.4, 8.3) | Guan, et al<sup>42</sup><br>Li, et al<sup>43</sup><br>Lauer, et al<sup>44</sup> |
| Average duration of infection, non-hospitalized individuals, $d_I$ (95% CI) | all | 13.1 (8.3, 16.9) | Huang, et al<sup>45</sup> |
| Average time from infection to hospitalization, $d_H$ (95% CI) | all | 10.3 (6.5, 13.3) | Wang, et al<sup>46</sup> |
| Average duration of hospitalization, individuals who recover, $d_R$, or die, $d_M$ (95% CI) | all | 14.4 (11.3, 16.6) | Lewnard, et al<sup>47</sup> |
| Probability case is clinical, $\Pr(\text{clinical}\vert \text{age})$ | $i < 20$<br>$i \geq 20$ | 0.21<br>0.69 | Davies, et al<sup>13</sup> |
| Probability infection is acquired from subclinical transmission, $\alpha$ | all | 0.50 | Davies, et al<sup>13</sup><br>Prem, et al<sup>41</sup> |
| Probability of hospitalization among clinical cases, $\Pr(\text{hospital}\vert \text{age})$ | $i < 10$<br>$10 \leq i < 20$<br>$20 \leq i < 30$<br>$30 \leq i < 40$<br>$40 \leq i < 50$<br>$50 \leq i < 60$<br>$60 \leq i < 70$<br>$70 \leq i < 80$<br>$i \geq 80$ | 0.00001<br>0.000408<br>0.0104<br>0.0343<br>0.0425<br>0.0816<br>0.118<br>0.166<br>0.184 | Verity, et al<sup>57</sup> |
| Probability of death among hospitalized patients, $\Pr(\text{death}\vert \text{age, hospital})$ | $i < 20$<br>$20 \leq i < 30$<br>$30 \leq i < 40$<br>$40 \leq i < 50$<br>$50 \leq i < 60$<br>$60 \leq i < 70$<br>$70 \leq i < 80$<br>$i \geq 80$ | 0.02<br>0.031<br>0.0475<br>0.0785<br>0.1215<br>0.186<br>0.301<br>0.4515 | Lewnard et al<sup>47</sup> |
| Ratio of susceptibility among adults to susceptibility among children, $\beta_{i<20}/\beta_{i\geq 20}$ | all | 0.50 or 1 | Various; (see Supporting Information) |

### Modeled contact rates and interventions

A shelter-in-place order was announced for six Bay Area counties on March 16^49^, following which only essential workers continued in-person work, and schools were closed. Between January 17 and March 16, transmission was simulated as described above, deriving community contact rates from POLYMOD data, national estimates of social mixing data during typical conditions.^50^

We simulated transmission March 17 - June 1, the remainder of the spring semester in the 2019-2020 academic year following issuance of shelter-in-place orders (Figure 1C), first under observed conditions: no school contacts, 28% workforce participation^51^, and community contacts derived from our social contact survey. Modelled output matched well with available data on hospitalizations, deaths, and seroprevalence (Figure S5). We then simulated the cumulative incidence that would have occurred over this period under counterfactual scenarios where: 1) schools remained open; 2) workplaces remained open; and 3) non-essential community contacts (including impersonal encounters on non-essential outings and social gatherings) continued.

Community contact matrices were derived for each intervention based on survey and POLYMOD data (Figure S2 and Table S4). For all counterfactual scenarios, except those permitting non-essential community contacts, we assumed 50% of household members of symptomatic cases reduced their community contacts by 75% and their work or school contacts by 100%.^30^ We estimated the number of cases, hospitalizations, and deaths averted by the intervention as the difference between these outcomes for the counterfactual scenarios minus the modelled observed scenario. Given substantial under-reporting of symptomatic cases^52^, we express findings as the percent increase from observed cases that were averted by the intervention.

Lastly, we simulated the effect of school reopening strategies for the fall semester (August 15 - December 20; Figure 1C). We established initial conditions for these simulations by initiating model runs spanning the spring and summer periods, and then modeled the effect of reopening strategies under two susceptibility assumptions (children <20 half vs. equally as susceptible as adults), and two transmission contexts (high and moderate community transmission). The high transmission context is characterized by 75% of workplaces remaining open and non-essential community contacts double what we observed in our survey; the moderate transmission context is characterized by 50% of workplaces remaining open and non-essential community contacts equal to that observed in our survey after Memorial Day (May 25).

At *t* = Aug 15, 2020, we updated contact rates to simulate six school reopening strategies (Figure 1C; see the Supporting Information for details): 1) schools open without precautions; 2a) classroom groups are enforced, reducing other grade and school contacts by 50% (weak stable cohort), or 2b) 75% (strong stable cohort); 3) class sizes are cut in half, and each half attends two staggered days each week; 4) class sizes maintained, and half the school attends two staggered days each week according to grade groups; 5) students and faculty wear masks; 6a) faculty and/or students are tested with 85% sensitivity on a weekly or 6b) monthly basis^53^, with positive cases isolated and their class quarantined for 14 days. We considered classroom groups to be a stable cohort averaging 20 students. Masks were assumed to reduce both outward and inward transmission by a factor of (1-*η*_i_), where *η*_i_ represents the effectiveness of the mask for agent *i*, allowed to increase across age groups (15% for elementary students, 25% for middle school students, 35% for high school students, 50% for faculty)^54^. We estimate excess infections (symptomatic only and all infections), hospitalizations, and deaths attributable to school-based transmission as the cumulative incidence of infections, hospitalizations, and deaths under each school reopening scenario minus the cumulative incidence under a school closure scenario. We then identify which set of interventions are needed to reduce excess risk of symptomatic illness for teachers (the sub-population determined to be at highest risk) such that less than one additional percent becomes infected.

### Role of Funding Source

Funders had no role in study design, data collection, data analysis, data interpretation, writing of the report, or the decision to submit for publication. The corresponding author had full access to all of the data and the final responsibility to submit for publication.Figure 1. Model schematic A) Schematic of the agent-based susceptible-exposed-infected-recovered (SEIR) model. S = susceptible; E = exposed, A = asymptomatic; C = symptomatic, will recover; H1 = symptomatic and will recover, not yet hospitalized; H2 = hospitalized and will recover; D1 = symptomatic, not yet hospitalized; D2 = hospitalized and will die; R = recovered; M = dead; *λ* = force of infection, which defines movement from S to E. Superscript *i* refers to individual. After an agent enters the exposed class, they enter along their predetermined track, with waiting times between stage progression drawn from a Weibull distribution. B) Schematic of the conditional probabilities by which agents are assigned a predetermined track. C) Schematic of interventions simulated in the SEIR model. The first analysis examines transmission between January 17 and June 1, and tests the effect of several counterfactual scenarios that took place between the enactment of Shelter in Place (March 16) and the original end of the spring semester (June 1). The second analysis examines transmission between January 17 and December 20, and tests the effect of several simulated reopening strategies for the Fall semester, expected to occur between August 15 and December 20, under a high and moderate community transmission scenario. Boxes represent categories of social contacts, including community (red), work (yellow), school (light blue), grade (medium blue) and classroom (dark blue). Percentages in the boxes represent the percentage of the contact rate experienced under a given intervention or counterfactual scenario (e.g. 0% represents a full closure).

**Figure 1.**
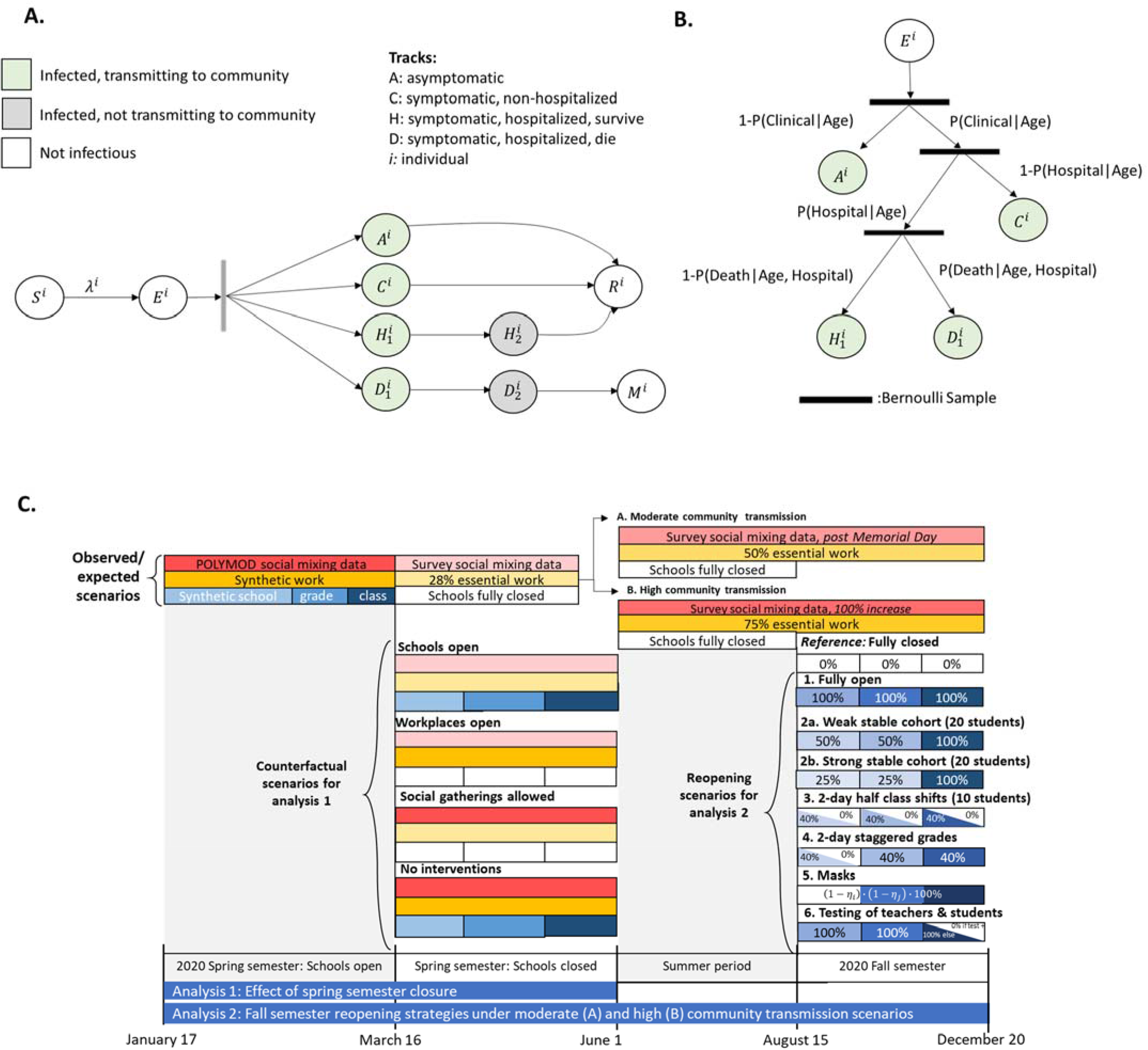
Model schematic. A) Schematic of the agent-based susceptible-exposed-infected-recovered (SEIR) model. S = susceptible; E = exposed, A = asymptomatic; C = symptomatic, will recover; H1 = symptomatic and will recover, not yet hospitalized; H2 = hospitalized and will recover; D1 = symptomatic, not yet hospitalized; D2 = hospitalized and will die; R = recovered; M = dead; *λ* = force of infection, which defines movement from S to E. Superscript *i* refers to individual. After an agent enters the exposed class, they enter along their predetermined track, with waiting times between stage progression drawn from a Weibull distribution. B) Schematic of the conditional probabilities by which agents are assigned a predetermined track. C) Schematic of interventions simulated in the SEIR model. The first analysis examines transmission between January 17 and June 1, and tests the effect of several counterfactual scenarios that took place between the enactment of Shelter in Place (March 16) and the original end of the spring semester (June 1). The second analysis examines transmission between January 17 and December 20, and tests the effect of several simulated reopening strategies for the Fall semester, expected to occur between August 15 and December 20, under a high and moderate community transmission scenario. Boxes represent categories of social contacts, including community (red), work (yellow), school (light blue), grade (medium blue) and classroom (dark blue). Percentages in the boxes represent the percentage of the contact rate experienced under a given intervention or counterfactual scenario (e.g. 0% represents a full closure).

## Results

### Contact patterns

612 households provided contact histories on behalf of 819 school-aged children in the Bay Area (Table S1). Comparison of contact matrices by location revealed that the majority of non-household contacts among survey respondents occurred between individuals in the same age category, and while performing essential activities (such as grocery shopping, laundering clothing or receiving health care), at work, home, or during an outdoor leisure activity (Figure 2A; Figure 2C). Younger children aged 5-12 years had twice as many non-household contacts (1.58 contacts per child per day) than teenagers aged 13-17 years (0.78 contacts per teenager per day) (Figure 2B).

**Figure 2.**
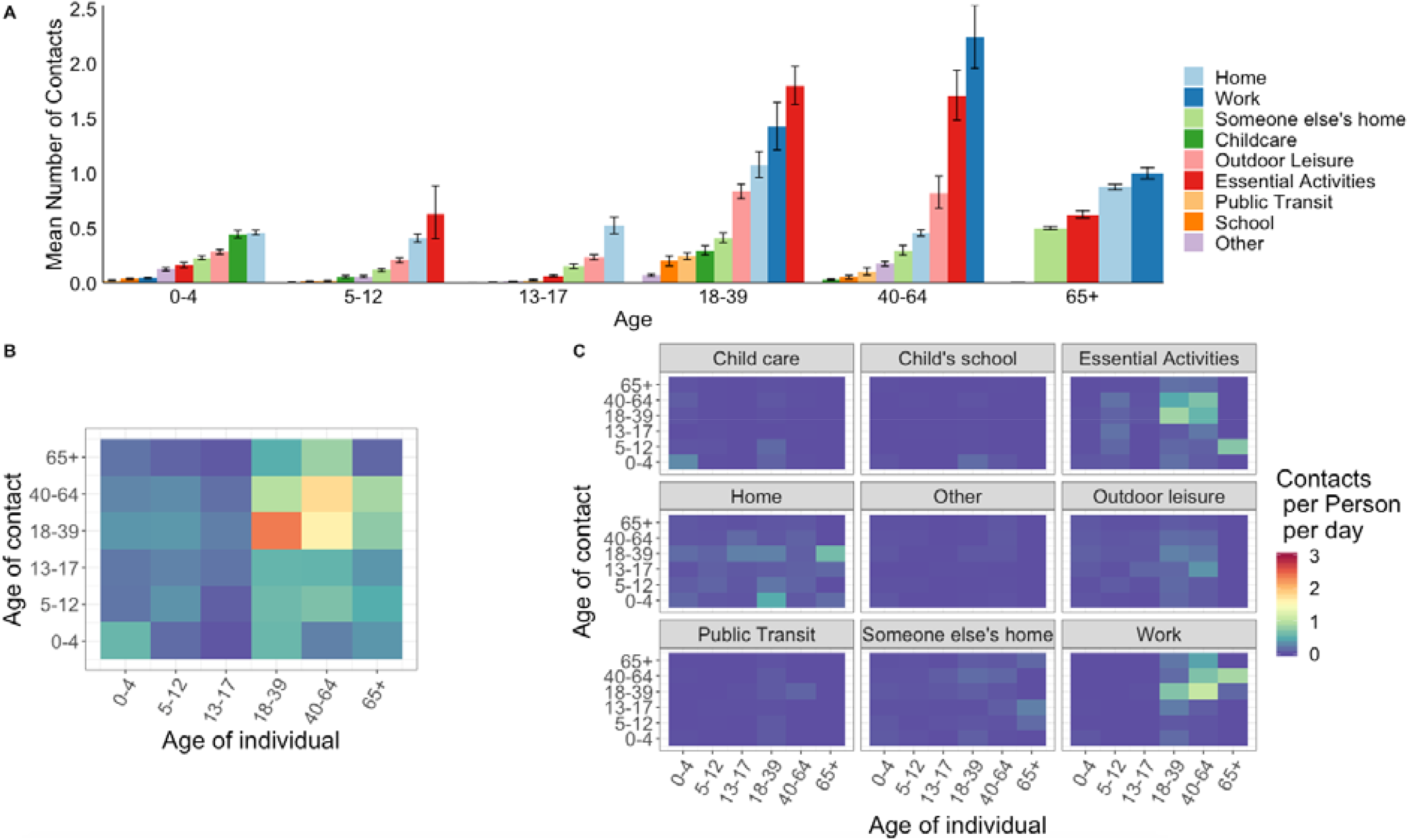
Summary of social contact patterns between children and primary household members of Bay Area (California) households between May 4 and June 1, 2020 following distribution of web-based contact diaries. A) Average number of contacts each age group self-reported at nine pre-specified locations. B) Average number of contacts self-reported per person per day by age category of the survey respondent and reported contact, unweighted. C) Average number of contacts self-reported per person per day at each of the nine locations. Panels B and C share a legend.

Adjusted models revealed disparities in total reported contacts across location of work (outside or inside the home) during shelter-in-place and ethnicity. Households identifying as primarily Hispanic or Latinx had 2.32 (95% CI: 0.08-4.50) more contacts on average compared to non-Hispanic or Latinx households (Table 2). Households that did not indicate an increase in the working from home during shelter-in-place compared to before shelter-in-place had 1.85 (95% CI: 0.16-3.52) more contacts than households with more adults working at home during shelter-in-place.

**Table 2.**
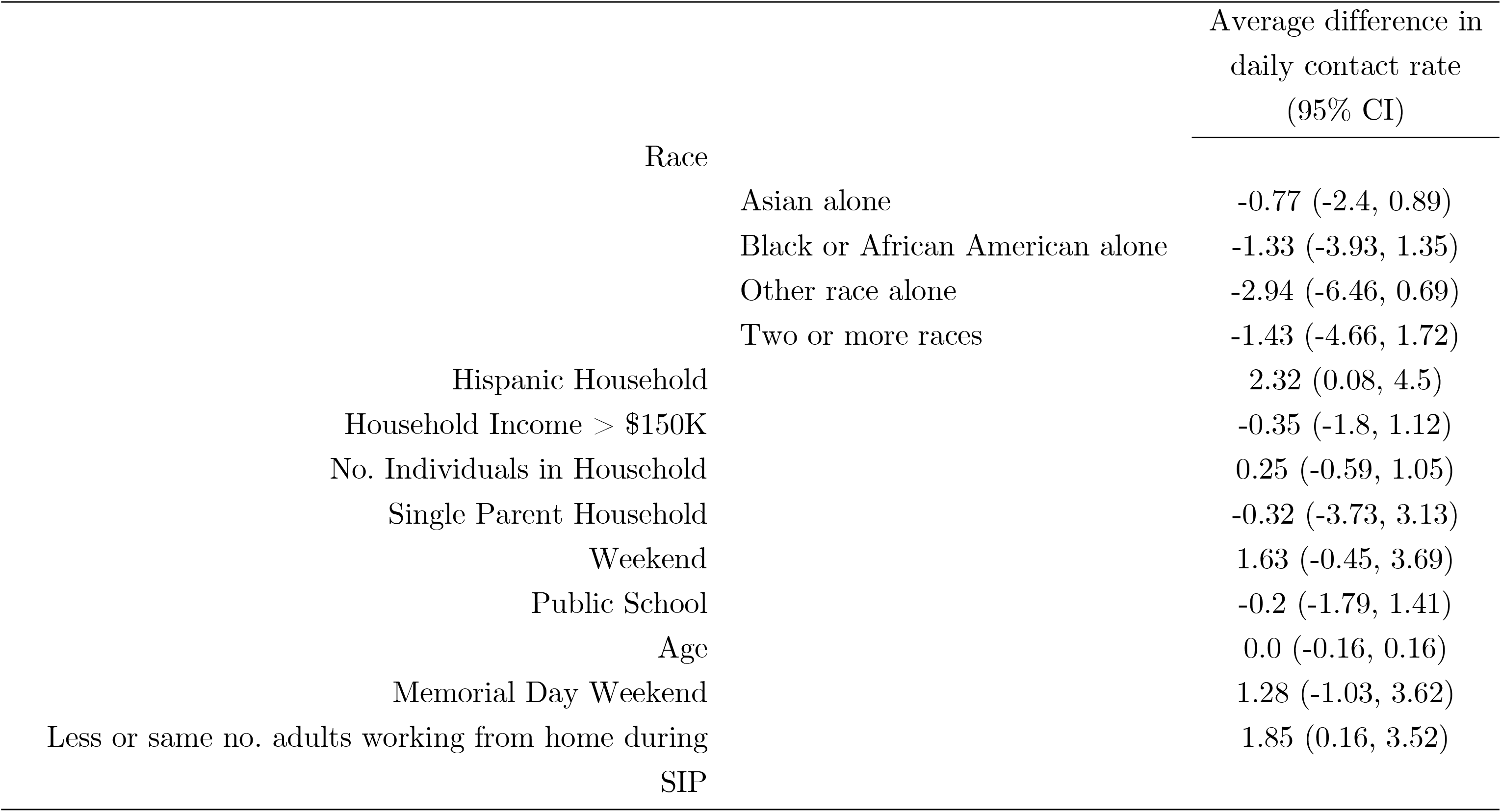
Results from linear mixed model that included random effects for each household. Estimates are adjusted for race (white alone as reference), self-reported household income (less than $150,000 annual income per household as reference), whether household identified as Hispanic (not Hispanic as reference), whether household was a single parent household (multi-parent household as reference), whether date of reported contacts were weekend or weekday (weekday as reference), whether child attended a public or non-public school (non-public school defined as private, charter, homeschool, or other, with non-public as reference), age of individual in years, whether the date of reported contacts occurred over memorial day weekend (defined as any survey completed May 24 - May 26, with not a holiday weekend as reference), and the change in number of adults working at home during shelter in place (with more adults working at home during SIP as reference). SIP = shelter-in-place.

|  | Average difference in<br>daily contact rate<br>(95% CI) |
| --- | --- |
| Race |  |
| Asian alone | -0.77 (-2.4, 0.89) |
| Black or African American alone | -1.33 (-3.93, 1.35) |
| Other race alone | -2.94 (-6.46, 0.69) |
| Two or more races | -1.43 (-4.66, 1.72) |
| Hispanic Household | 2.32 (0.08, 4.5) |
| Household Income > \$150K | -0.35 (-1.8, 1.12) |
| No. Individuals in Household | 0.25 (-0.59, 1.05) |
| Single Parent Household | -0.32 (-3.73, 3.13) |
| Weekend | 1.63 (-0.45, 3.69) |
| Public School | -0.2 (-1.79, 1.41) |
| Age | 0.0 (-0.16, 0.16) |
| Memorial Day Weekend | 1.28 (-1.03, 3.62) |
| Less or same no. adults working from home during<br>SIP | 1.85 (0.16, 3.52) |

### Impact of spring 2020 school-closure policies

#### Assuming children <10 years are half as susceptible to SARS-CoV-2 compared with older children and adults

Had all Bay Area schools remained open between March 17 and June 1, we estimate a higher burden of disease would have been experienced in Bay Area populations in the spring of 2020. For a synthetic population derived from population characteristics of Oakland, California, with a ratio (a) of the force of infection of asymptomatic individuals to symptomatic individuals of 0.5, and susceptibility of children under 10 years set to half that of older children and adults, there would have been 1.98 (95% CI: 0.44, 2.6) times more cases of COVID-19 throughout the nine Bay Area counties than observed (Figure 3), with 3.16 (95% CI: 1.79, 4.89) times more cases among families of students grades K-12. As of June 1, 2020, the Bay Area had 14,202 reported cases of COVID-19^58^, and we estimate that closing schools, especially middle and high schools, averted an additional 13,842 (95% CI: 6,290, 23,040) confirmed cases. We estimate that if elementary schools alone had remained open, the Bay Area would have recorded 2,167 additional cases (95% CI: −985, 5,572), while if only middle schools had remained open, an additional 5,884 cases (95% CI: 1,478, 11,550) would have been observed, and if high schools alone had remained open, an additional 8,650 cases would have been observed (95% CI: 3,054, 15,940). By comparison, had all workplaces remained open, we estimate that, as of June 1, there would have been 15,813 additional confirmed cases (95% CI: 9,963, 22,617), reflecting 2.11 (95% CI: 1.70, 2.59) times more cases than observed. If non-essential outings and social gathering had been permitted, we estimate that we would have seen an additional 7,030 (95% CI: 3,118, 11,676) confirmed cases, reflecting 1.50 (95% CO: 1.22, 1.82) times more cases than observed. All three interventions together helped avert an estimated 49,023 confirmed cases.

We find that both school and workplace closures in the spring of 2020 were necessary to achieve a sustained *R*<1. We estimated that the highest COVID-19 hospitalization rate that would have been observed on any one day during shelter-in-place if schools were open was 10.6 (95% CI: 6.0, 16.0) per 10,000 population, representing an excess of 4.42 individuals per 10,000 from the modelled observed hospitalization rate. As the Bay Area hospital capacity amounts to 22 beds per 10,000 population, school closures permitted more than one-fifth of beds to remain available, but were not necessary to keep the Bay Area healthcare systems under capacity.^59^ As of June 1, 2020, the Bay Area had 3,997 confirmed deaths from COVID-19.^58^ We estimate that school closures averted 0.63 deaths (95% CI:-1.25, 3.75) per 10,000 population, corresponding to 663 deaths across the Bay Area, fewer than workplace closures (estimated 828 deaths averted) and more than other socializing restrictions (estimated 503 deaths averted).

**Figure 3.**
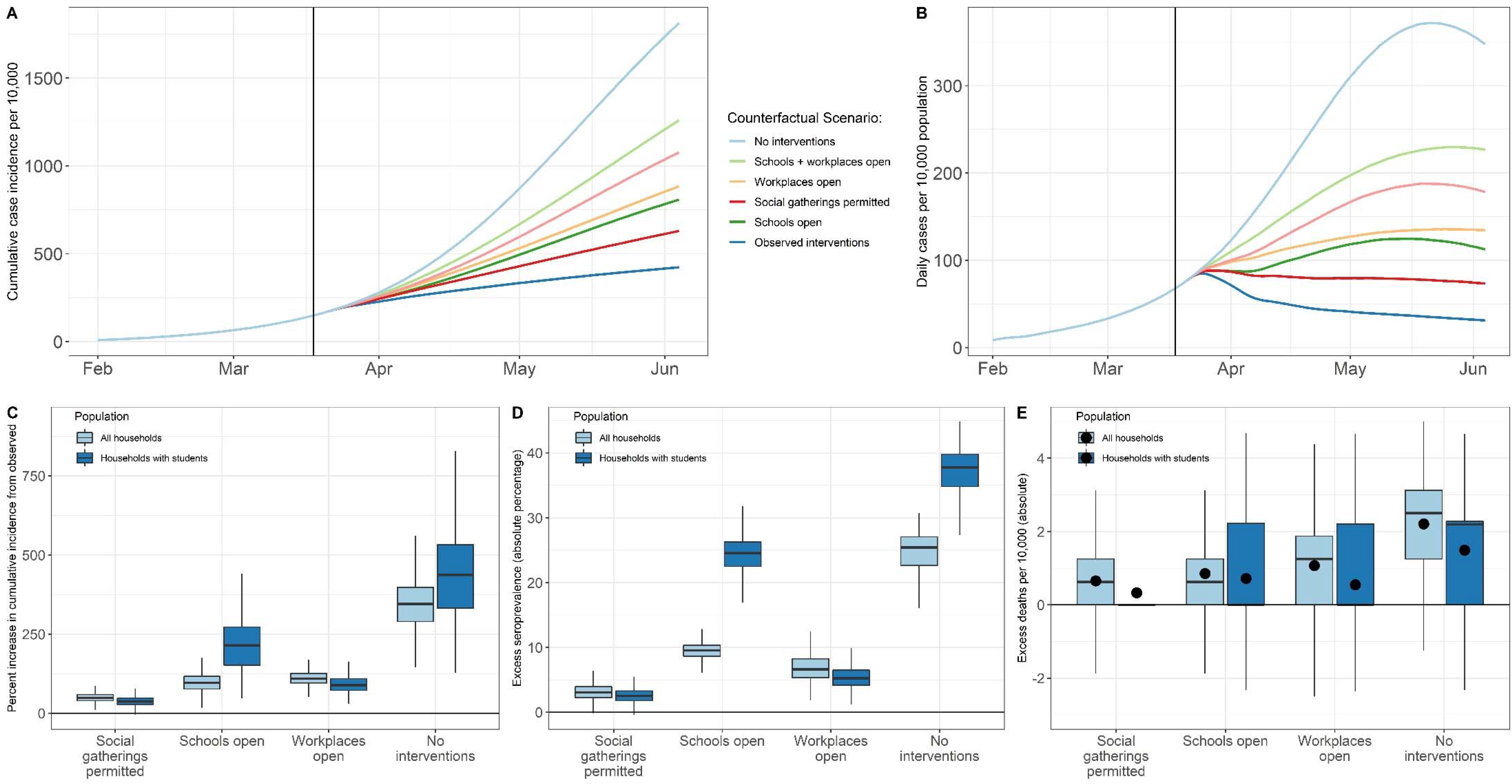
Effect of spring semester interventions. We simulated transmission between February 17 and June 1 assuming children <10 years are half as susceptible to infection compared with older children and adults. Between March 16 (enactment of shelter-in-place orders) and June 1 (the end of the spring school semester), we assessed potential outcomes under various counterfactual scenarios: 1) schools had remained open for the remainder of the school semester; 2) workplaces had remained open; 3) social gatherings were permitted; 4) no interventions were enacted. A) Modelled cumulative incidence according to the counterfactual scenario examined. Modelled predictions are not adjusted for under-reporting, which is expected to be substantial. B) Daily incidence per 10,000 per counterfactual scenario examined. C) The percent increase in cumulative incidence from observed incidence between February 17 and June 1, stratified by counterfactual scenario and population sub-group. D) The absolute difference in the percent of population seropositive for each counterfactual scenario compared to the modelled, observed seroprevalence between February 17 and June 1, stratified by population sub-group. E) The percent increase in deaths per 10,000 from observed between February 17 and June 1, stratified by counterfactual scenario and population sub-group. The distribution of estimated death rate across 1,000 realizations was skewed, so black dots representing the mean number of excess deaths per 10,000 are added.

#### Assuming individuals <20 years are half as susceptible to SARS-CoV-2

The estimated impact of school closures in spring 2020 strongly depended on the relative susceptibility of children to adults (Figure 4A). Under the assumption that all individuals under 20 years are half as susceptible to SARS-CoV-2 compared with adults, school closures would be the least effective intervention when compared to workplace and social distancing strategies, avoiding an estimated 4,179 cases (95% CI: 308, 10,583) and 202 deaths (0.26 deaths per 10,000 population, 95% CI: −1.25, 2.50) between March 17 - June 1 across the Bay Area.

**Figure 4.**
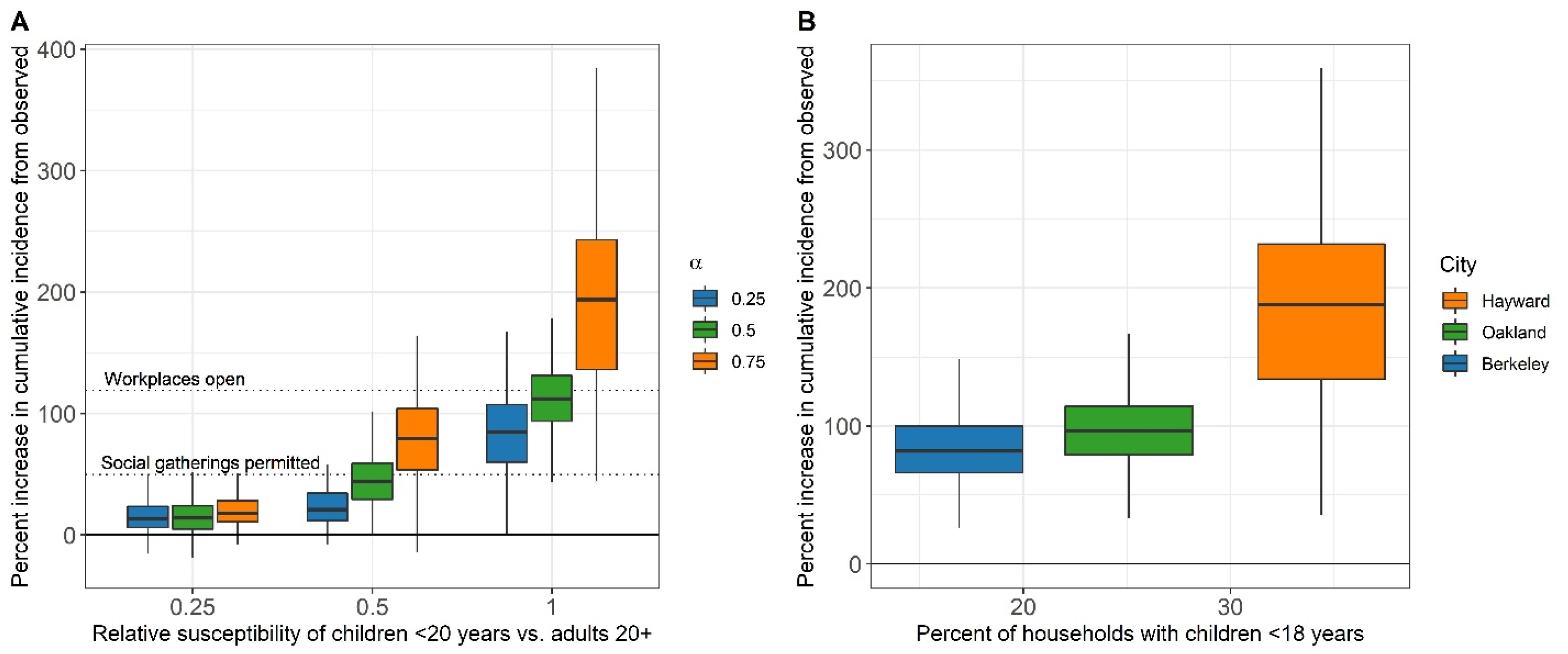
Influence of key epidemiological parameters on the effectiveness of school closures. The percent increase in cumulative incidence from observed incidence over the period February 17 - June 1 had schools remained open between March 17 and June 1. A) Results are reported for modelling scenarios that varied the ratio of the susceptibility of individuals under 20 years to adults 20 or older, and the ratio of the force of infection for asymptomatic infections to symptomatic infections (α). Dashed lines indicate the percent increase in incidence from observed that would have been expected if workplaces had remained open, and if social gatherings were permitted. B) Results are reported for synthetic populations with varying levels of the proportion of households with children under 18 years of age, reflecting three major Bay Area cities (Berkeley, Oakland, Hayward, assuming children under 10 are half as susceptible as older children and adults.

#### Assuming equal susceptibility across all ages

Under the assumption of equal susceptibility among all ages, the estimated impact of school closures quadruples, from 4,179 averted cases to 16,348 (95% CI: 8,325, 25,363) averted cases, making school closures the most effective intervention. Likewise, under equal susceptibility of all individuals, the estimated number of deaths averted by school closures in the nine Bay Area counties between March 17 and June 1 more than triples, from 202 to 655 averted deaths, corresponding to an excess death rate of 0.84 (−1.25,3.13) per 10,000 population. The excess death rate from workplace closures is only slightly higher, at 0.90 excess deaths per 10,000 (95% CO: −1.25, 3.13) between March 17 - June 1. At low levels of susceptibility (i.e., one-fourth that of adults) among children, the impact of school closures was small, and the ratio of the force of infection of asymptomatic individuals to symptomatic individuals (a) had little influence on the impact of spring school closure policies (Figure 4A). As children increase in susceptibility relative to adults, the influence of *a* becomes more pronounced (Figure 4a).

We found a significant positive relationship between the number of cases averted by school closures and the proportion of households in the population with children under 18 years (Figure 4B). For each 1% increase in the proportion of total households that have children under 18, we estimate an additional 5.8% increase over observed incidence had schools remained open throughout the spring semester.

### Simulated impact of fall 2020 reopening strategies

The estimated risk of symptomatic infection during the fall 2020 semester—across moderate to high transmission contexts—is highest for teachers and all other school staff, followed by students and other household members of students and teachers/staff (Figure 5A). Owing to larger average school sizes, high schools are at higher risk, followed by middle schools, then elementary schools. Staggered 2-day school weeks with halved class sizes provided the largest reduction in risk, followed by strong stable cohorts of class groups, then mask wearing. In the absence of other interventions, testing and isolation/quarantine strategies have low effectiveness, but when combined with strict social distancing measures, a modest reduction in community cases is possible as infectious individuals and their contacts identified in the school environment are quarantined (i.e., have their community contacts reduced by 75% for 14 days). Excess seroprevalence, hospitalizations, and deaths associated with fall school reopening, as they vary across child susceptibility and transmission contexts, are detailed in Tables S5-S8.

#### Assuming individuals < 20 years are half as susceptible

At moderate levels of community transmission, and with no precautions taken within school settings, we estimate that an additional 21.0% (95% CI: 0, 46.0%) of high school teachers, 13.4% (95% CI: −2.2, 38.6%) of middle school teachers, and 4.1% (95% CI: −1.7, 12.0%) of elementary school teachers would experience symptomatic illness between August 15 and December 20, compared to expectations if schools were closed (Figure 5). We estimate that the daily hospitalization rate would increase by an average of 0.53 (95% CI: −0.58, 1.73) hospitalizations per 10,000 individuals (roughly 2.5% of Bay Area hospital bed capacity), of which 0.13 (95% CI: −0.29, 0.58) and 0.33 (95% CI: −0.58, 1.30) per 10,000 would be among household members of students and other community members, respectively (Figure 6B). We estimate an excess total death rate of 0.56 (95% CI: −1.88, 3.13) per 10,000, corresponding to 434 (95% CI: −1,451, 2,418) deaths across the Bay Area, of which 287 would be among community members without students in their household, 114 among household members of students, and 31 among teachers; only one death was expected among students. At high community transmission (similar to observed in July 2020), and with no precautions taken within school settings, we estimate that an additional 33.3% (95% CI: 11.1, 53.6%) of high school teachers, 24.4% (95% CI: 4.3, 44.4%) of middle school teachers, and 9.1% (95% CI: 0.9, 20.0%) of elementary school teachers would experience symptomatic illness (Figure 5). We estimate that the daily hospitalization rate would increase by an average of 1.65 (95% CI: −0.17, 3.38) hospitalizations per 10,000 individuals, of which 0.37 (95% CI: −0.22, 1.01) and 1.17 (95% CI: −0.36, 2.70) per 10,000 would be among household members of students or teachers and other community members, respectively (Figure 6B). We estimate an excess total death rate of 1.73 (95% CI: −2.50, 6.25) per 10,000, corresponding to 1,341 (95% CI: −1,934, 4,837) deaths across the Bay Area, of which 1,026 would be among community members, 254 among household members, 60 among teachers, and one death among students.

At moderate community transmission, we estimate that strict adherence to staggered school weeks (either as half classes or grades), or combining stable cohorts (weak or strong) with masks (with at least 35% effectiveness in students and 50% in teachers) and monthly testing, is needed to reduce excess risk of symptomatic illness for teachers to less than 1% (see Table 3, which also details interventions necessary in high transmission contexts). Interventions such as strong stable cohorts, 2-day staggered grades, or strong stable cohorts combined with masks and testing would decrease the expected total number of excess deaths by 85%, 95%, and 95%, respectively.

**Table 3.**
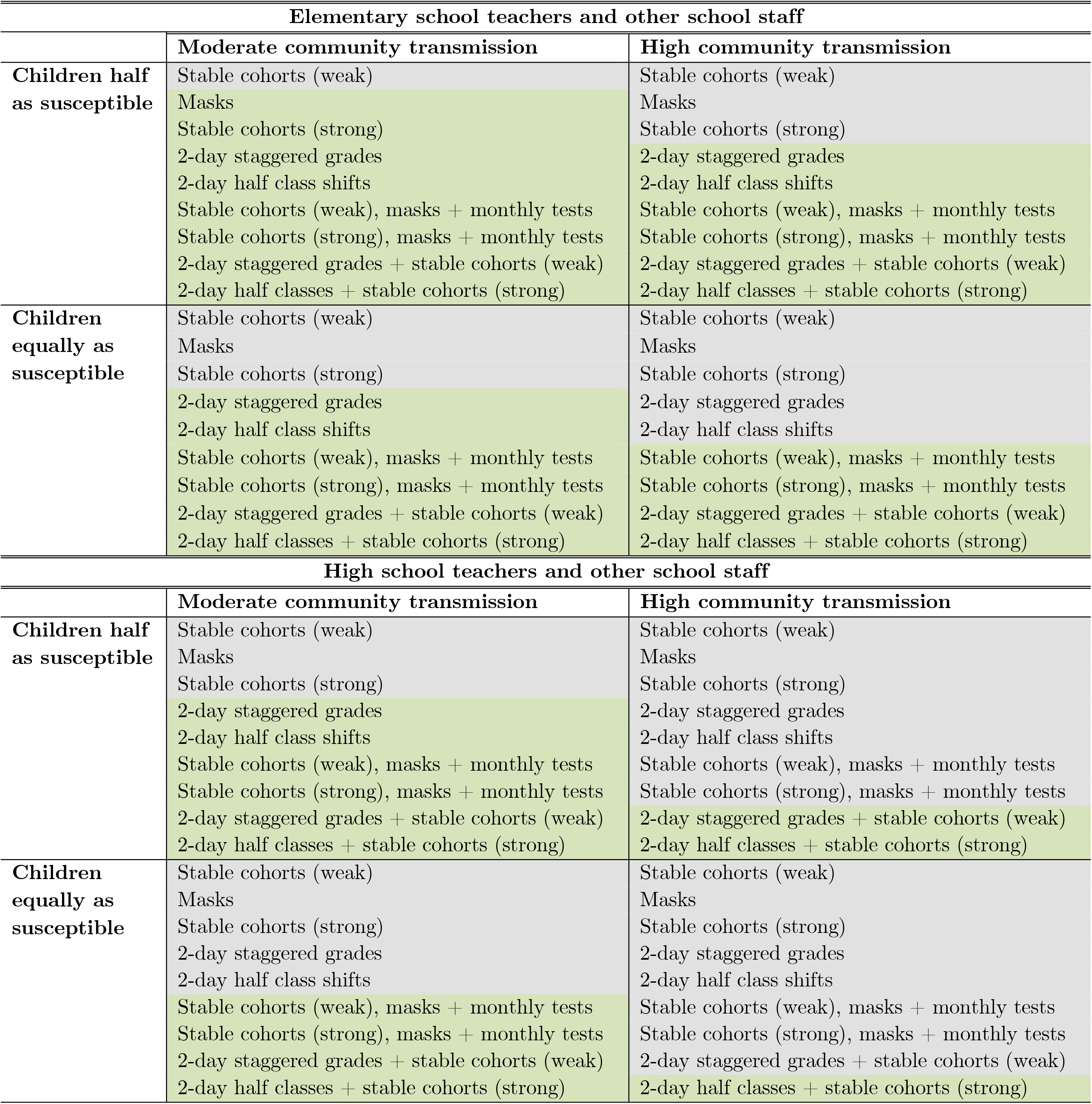
School-based interventions to reduce risk. This table colors the reopening strategies examined for the fall 2020-2021 academic year by whether or not they are sufficient to reduce the additional proportion of teachers and other school staff experiencing symptomatic illness between August 15 and December 20 to <1% of teachers. Strategies colored in green are strategies which reduce the excess number of teachers with symptomatic illness to <1%. Strategies colored in gray are strategies which do not reduce the excess number of teachers with symptomatic illness to <1%. Results are stratified by high school and elementary school teachers.

We estimate that reducing community transmission significantly reduces the excess risk to teachers across all grades, from 18.4% (95% CI: 7.7, 27.9%) to 10.3% (95% CI: 0.4, 20.7%), with the influence of community transmission levels minimized as school-based interventions become stronger. The level of community transmission strongly determines whether the effect of school reopenings will be associated with increased incidence among the general community (non-students, teachers or family members). In high transmission settings where schools open without precautions, we estimate that the majority (59%) of the excess cases will be among community members, whereas in moderate transmission settings, fewer than half (45%) of the excess cases will be among community members (Figure 6A),In scenarios evaluating both moderate and high community transmission, when susceptibility to infection is assumed constant across all ages, we estimate a higher proportion of additional clinical infections amongst all sub-populations and reopening strategy compared to the reopening scenario where children were half as susceptible (Figure 5). Notably, if no precautions are taken within school settings, at moderate levels of community transmission, we estimate nearly four times as many elementary school teachers will experience additional clinical infections if children are equally susceptible (17.3%, 95% CI: 4.4, 30.0%) compared to the equivalent scenario where children are half as susceptible (4.1%, 95% CI: −1.7, 12.0). Similarly, over three times as many middle school teachers (37.2%, 95% CI: 4.6, 58.1% vs. 13.4%, 95% CI: −2.2, 38.6%), and nearly two times as many high school teachers (40.7%, 95% CI: 1.9, 61.1% vs. 21.0%, 95% CI: 0, 46.0%) will experience symptomatic illness when comparing the relative susceptibility of children at moderate levels of community transmission if no additional precautions are taken in school settings. At moderate levels of community transmission, increasing the relative susceptibility of children to adults also quadrupled the excess daily hospitalization rate in moderate transmission scenarios from 0.53 hospitalizations per 10,000 individuals when children are half as susceptible to 2.00 (95% CI: 0.36, 3.67) hospitalizations per 10,000 individuals if children are equally susceptible, leading to more than four times the number of absolute deaths amongst community members (287 community member deaths if children are half as susceptible vs. 1,159 community member deaths if children are equally as susceptible) (Figure 6B).

We found that regardless of the relative susceptibility of children to adults, across both moderate and high community transmission settings, a strict adherence to a combination of strong distancing interventions (e.g., combining staggered half classes or staggered grades with stable cohorts; combining stable cohorts with mask wearing and monthly testing) is needed to reduce the excess risk of symptomatic illness for high school teachers and all other school staff to less than 1% (Table 3). The benefit of having a strong (75%) versus a weak (50%) reduction in non-classroom (non-cohort) contacts is most notable when children are highly susceptible. For instance, in a high transmission context, reducing non-classroom contacts by 50% and 75% lowers the excess risk to all teachers from 32.1% to 15.3% and 5.3%, respectively. If children are half as susceptible, the excess risk to all teachers is lowered from 18.4% to 5.2% and 3.4%, respectively (Figure 5).

**Figure 5.**
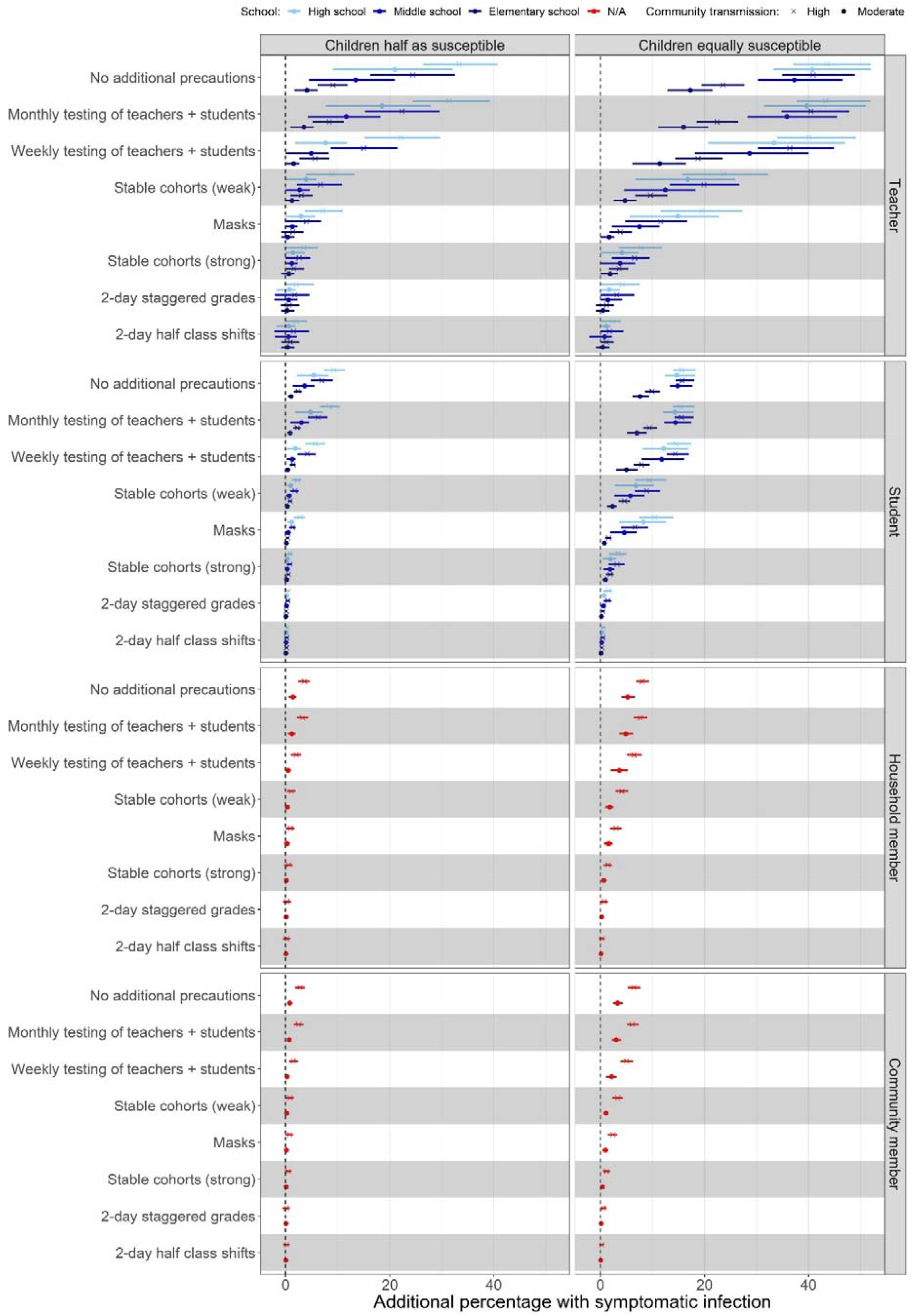
Excess risk by subgroup associated with school reopening strategies for the 2020 Fall semester. Panel A shows the additional proportion (mean and IQR) of each subgroup expected to experience clinical infection over the course of the fall 2020 semester (August 15 - December 20) compared to if schools were closed under each reopening scenario and the four transmission contexts: children half and equally as susceptible as adults crossed with moderate and high community transmission. Colors indicate the transmission across levels of schooling (elementary, middle, and high) while the shape of the mean point indicates the level of community transmission (circle = moderate, cross = high). “Teachers” include teachers and all other school staff.

**Figure 6.**
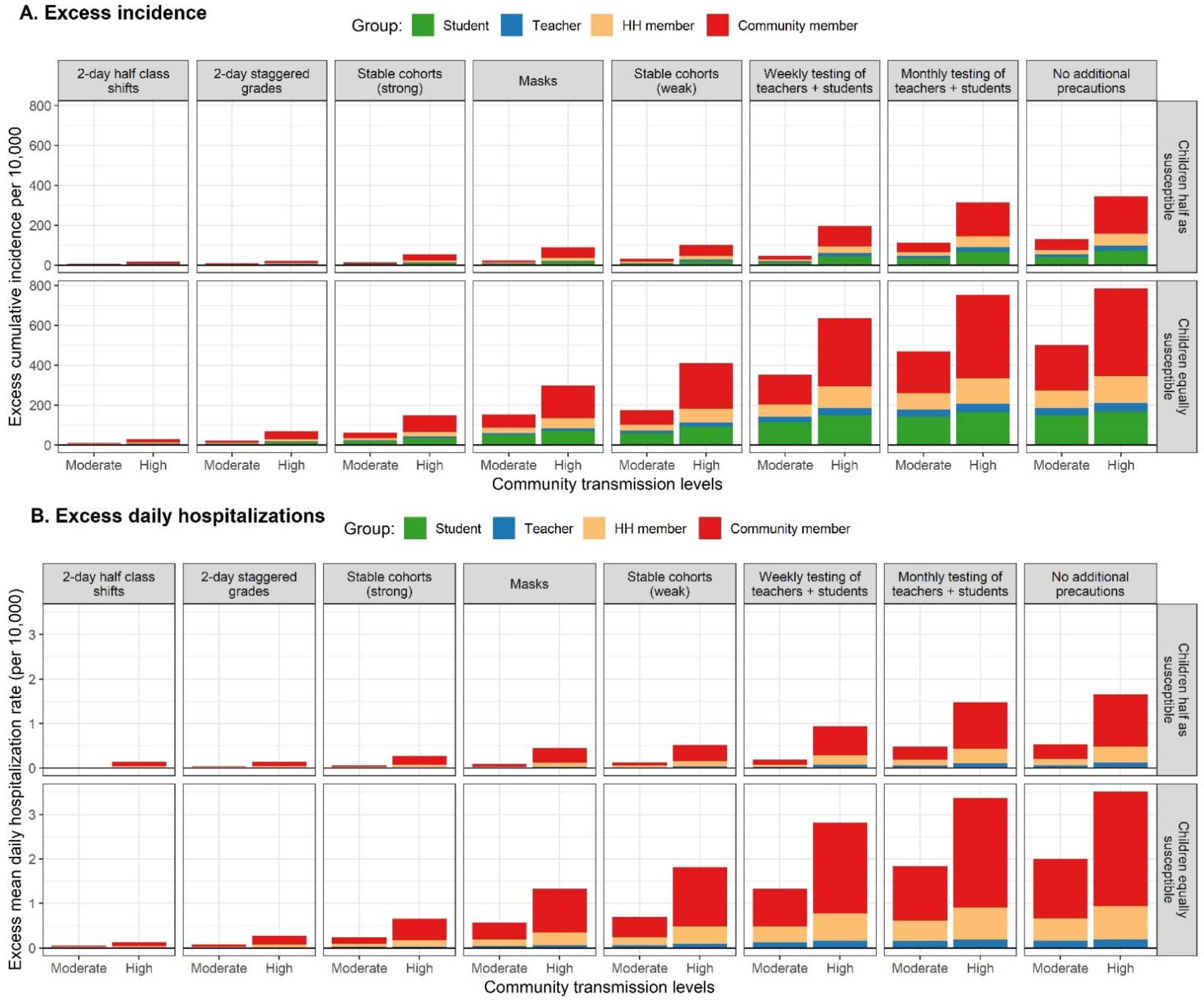
Population level excess incidence and hospitalizations association with reopening strategies for the 2020 fall semester. Panel A shows the excess cumulative incidence per 10,000 that would be expected between August 15 and December 20 for each reopening strategy compared to if schools were closed. Bars are stratified by the moderate and high community transmission scenario and colored according to the subgroup contributing cases. Panel B shows the excess daily hospitalizations, on average, per 10,000 that would be expected between August 15 and December 20 for each reopening strategy compared to if schools were closed. Bars are stratified by the moderate and high community transmission scenario and colored according to the subgroup contributing cases. “Teachers” include teachers and all other school staff.

## Discussion

Gaps in our understanding of contact patterns among US schoolchildren have limited previous efforts to estimate the effect of school closures on COVID-19 transmission in a community of demographically heterogeneous households. Consistent with reports that systemic health and social inequities have disproportionately increased the risk of COVID-19 infection and death among low-income communities and racial and ethnic minorities^60,61^, our contact survey found evidence of a higher average community contact rate among low income and Hispanic children during shelter-in-place orders, which supports the growing body of evidence that physical distancing measures pose lesser benefit in low-income communities.^62^ The largest increases in contacts of Hispanic respondents compared to non-Hispanic respondents occurred among working-aged adults (18-65 years) and young children (0-12 years). As Hispanic individuals make up a disproportionate number of essential workers in the Bay Area^51^, these findings may reflect both higher working contacts as well as a need to find child care for young children during the caretaker’s working hours. Such challenges associated with school closures may be experienced more strongly among households with essential workers, even as we expect that school closures will avert more cases among these populations due to higher transmission levels. While our survey found higher contact rates in elementary students during shelter-in-place as compared to high school students, prior social mixing data report higher community contact rates among high school students.^50^ Elementary students were more likely than high school students to accompany their adult family members in performing essential activities, and had more daycare contacts. This may suggest that young children are less able to reduce non-household contacts during school closures and shelter-in-place orders than are older children.

In the March 17 - June 1 spring 2020 semester period, we estimate that school closures averted 13,842 confirmed cases, and 663 deaths in the Bay Area, with most of the averted cases and deaths due to high school and middle school closures. These estimates depend, in part, on epidemiologic parameters that remain imperfectly understood, such as the susceptibility of children and the degree to which asymptomatic individuals contribute to transmission. We present modeling results across four transmission contexts, varying community transmission levels and the relative susceptibility of children compared to adults in order to explore their impact on estimated transmission across fall reopening policies. To understand the full impact of closures and reopening policies, contact tracing studies that seek to capture the relative susceptibility and infectiousness of both symptomatically and asymptomatically infected children across ages are urgently needed. Policy makers should acknowledge that uncertainty around susceptibility and infectiousness of children exist when making decisions about school closure and act with precaution.

We find that the safest school reopening strategy involves controlling community transmission by enforcing other non-pharmaceutical interventions such as closure of non-essential workplaces and minimization of social gatherings. To prioritize opening of schools, policies should limit the opening of nonessential services for adults if opening such services will force schools to remain closed.^4^ We estimate that when workplace and social controls are minimal, the excess risk for high school and elementary school teachers and staff increases by 50%, and the risk to elementary school teachers more than doubles. What is more, in a high transmission context, our model estimates that the majority of excess cases attributable to opening schools will be experienced by individuals from households unaffiliated with schools, rather than by students, teachers, or household members of students and teachers/staff combined. Concerns have been raised that school-based outbreaks can spill over into the general population where subsequent transmission can propagate readily, as documented in Israel when the opening of middle and high schools was associated with high and sustained community transmission.^35^ Heterogeneity in risk of infection across community transmission contexts falls sharply as school-based interventions increase in effectiveness—that is, strong infection control measures in schools minimizes the influence of community transmission on school-based transmission. School reopening guidelines should include contingency plans for situations where community transmission increases, potentially phasing in additional safeguards, or temporary closures if the school cannot feasibly implement such measures. However, this could lead to a situation in which well-resourced schools in low transmission areas remain open, putting unsafe pressure on low-resourced public schools in high transmission areas to stay open, or further increasing inequities caused by school closures. Accordingly, resources directed to schools for control measures should first go to public schools in high transmission areas that may not have the resources to implement and maintain an effective set of interventions.

Even as community transmission is lowered, adherence to some set of school-based interventions is needed to reduce the excess risk of symptomatic illness at all levels of education. Under the lowest risk scenario examined (moderate community transmission, 79% of infected children as asymptomatic carriers, and low susceptibility of children relative to adults), we find that reopening for the fall semester without any precautions will yield substantial risk for students (an additional 3.0% of students across all grades levels infected over the fall semester), family members of students (an additional 1.4% infected), and especially teachers/staff (an additional 10.3% across all grade levels). This is consistent with evidence of high transmission among a summer camp where children interacted in large cohorts^34^ as well as high seroprevalence among teachers and students from a high school setting with limited safety measures.^33^ We estimate that school reopening without any precautions would increase hospitalizations such that an additional 2.5 - 16% of Bay Area hospital bed capacity (22 beds per 10,000 residents^59^) is occupied.

While the effectiveness of specific protective measures during reopening depended strongly on the level of community transmission and the relative susceptibility of children and adults, the relative impact of interventions remained consistent across all scenarios. To reduce the increase in symptomatic illness to below 1% in each population sub group is most feasible in elementary schools (using, for instance, mask wearing and weak stable cohorts). These findings are consistent with modelling results from the UK, which support a cautious reopening of elementary schools when *R* is less than one^63^, as well as other reviews.^4^ Achieving the same protection within high schools, by comparison, would require combining and maintaining two or more strict social-distancing interventions, such as staggered 2-day school weeks, mask wearing, and stable cohorts, which may present a challenge as high school students often interact with several different classroom groups across a single school day. However, a staggered school schedule is likely more feasible for high school families as compared to elementary school families, as teenagers may be more amenable to self-remote instruction.

Children’s social contact networks vary substantially by age—both within and outside of the school community—with critical consequences for the development of safe school reopening policies. High school environments have larger student, teacher and staff populations, resulting in significantly increased risks of transmission attributable to reopening. Even if younger children are as susceptible as older children, reopening high schools without precautions yields an estimated 3-5 times more risk of symptomatic infection to teachers/staff when compared with reopening of elementary schools, depending on the level of community transmission. If susceptibility increases with age, as some evidence suggests^13^,^57^,high school teachers may experience as much as 5-10 times more risk of symptomatic infection when compared with elementary school teachers, depending on the level of community transmission (moderate-high).

Age-structured contact rates captured by our survey have limited generalizability, as the composition of households in the Bay Area differs from the broader United States in several dimensions: the average household income is higher, there is higher overall educational attainment, a larger workforce, smaller household sizes, a smaller relative proportion of African Americans, and higher levels of social distancing^64^ and mask use.^65^ However, the age-structured contact rates from the Bay Area are similar to those captured from households with children from other major cities, including New York, Atlanta, Phoenix, and Boston.^66^ During the spring semester, the Bay Area had a higher proportion of essential workers (28%) than the national average,^51^ which could translate into a larger impact of workplace closures in cities outside the Bay Area. The impact of school closures also varies by the proportion of households that have school aged children, as well as the average school and class size of local public schools. Accordingly, the risk associated with school-based transmission will be higher in cities with a greater proportion of school aged children, as well as larger school or classroom sizes. Nevertheless, we expect that several findings pertaining to school reopening may translate to other settings—such as teachers experiencing the highest risks; high schools being at higher risk than elementary schools; high community transmission increasing risk; and the relative ranking of interventions. After all, key epidemiologic parameters (e.g., susceptibility of children, asymptomatic transmission, mask effectiveness) likely apply across locations, and several population-level parameters (e.g., average household size, birth spacing, mother’s age at first parity) likely apply to urban areas outside the Bay Area.

Another limitation relates to the social contact survey sample, which provided incomplete representation of the racial and economic diversity of the Bay Area. Selection bias likely arose from its administration in English, and because survey respondents were less likely to be essential workers. Discrepancies observed in the number of contacts by work location (outside vs. inside the home) and ethnicity (Hispanic or Latinx vs. non-Hispanic or Latinx) are thus expected to be biased towards the null. Since the survey was only administered to households with children, our community contact matrix was unable to capture contact patterns among and between households who do not have children, particularly missing those of young adults (18-29) or older adults (65+). However, our results are similar to estimates captured in another contact survey implemented in the Bay Area with a target population of households with and without children.^66^ Community contacts under modelled school closure scenarios account for increases in daycare contacts only at the rates observed in our community survey, when fewer adults were permitted to have in-person work. Therefore, modelled school closures or staggered weeks during the fall semester may not adequately account for increases in community contacts from daycare settings.

In addition to uncertainties in child susceptibility and community transmission, uncertainty in how the per-contact risk of transmission varies across settings (e.g. household environment vs. classroom) is a limitation. Similarly, we do not capture how setting (indoor vs. outdoor) and duration may increase the risk of COVID-19 transmission, which have direct implications for standard classrooms with poor ventilation and high durations of contacts.^4^ In 2013, over half of California elementary school classrooms did not meet state standards for classroom ventilation.^67^ Our modeled interventions were also limited, and did not include infection control measures, such as increased handwashing, desk spacing, or reduced sharing of classroom supplies, which may further reduce transmission in a school environment, and should be included in school reopening plans.

Given the myriad individual and societal consequences of school closures, policymakers must urgently dedicate financial resources to support the package of interventions necessary to mitigate risk in schools, and take immediate action to reduce community transmission while considering whether opening other sectors of society will increase community transmission to a point where it precludes school reopening. Focus should be placed first on elementary schools, where a more limited set of interventions may be required, and risk of school-associated transmission lower.

## Data Availability

COVID-19 data used in models are publicly available. Social contact data is available by request in aggregate form.

## Acknowledgements

We are grateful to Laura Kurre for valuable input in the discussion and conversations regarding the concerns of teachers and Dr. Naomi Bardach for sharing her expertise on the most up-to-date understanding of children and COVID-19 as well as the feasibility of school-based interventions. We thank Dr. Jon Krosnik for advice regarding survey questionnaire development. We thank Shelley Facente and Dr. Stephanie Holm for assistance in distributing the survey.

## SUPPORTING INFORMATION

The effect of school closures and reopening strategies on COVID-19 infection dynamics in the San Francisco Bay Area: a modeling study based on a survey of children’s social contact structure

Contents

Notice of approval for human research 2

Community Contact Survey 3

Inverse probability weighting of survey responses 17

Construction of synthetic population for transmission model 17

Transmission model details 17

Description of reopening strategies 20

Choice of susceptibility parameters based on available literature 22

Figure S1. Validation of synthetic population 26

Figure S2. Location stratified contact matrices adjusted for non-response 27

Figure S3. Age-specific contact matrices used for each counterfactual scenario 28

Figure S4. Community contact matrices by household characteristics 29

Figure S5. Comparison of model to observed data 30

Table S1: Synthetic population parameters 32

Table S2. Representativeness of survey sample to Bay Area population 33

Table S3. Characteristics of survey respondents 34

Table S4. Composition of community matrices 35

Table S5. Effect of fall semester reopening strategies: children half as susceptible, moderate community transmission 36

Table S6. Effect of fall semester reopening strategies: children half as susceptible, high community transmission 38

Table S7. Effect of fall semester reopening strategies: children equally as susceptible, moderate community transmission 40

Table S8. Effect of fall semester reopening strategies: children equally as susceptible, high community transmission 42

Notice of approval for human research

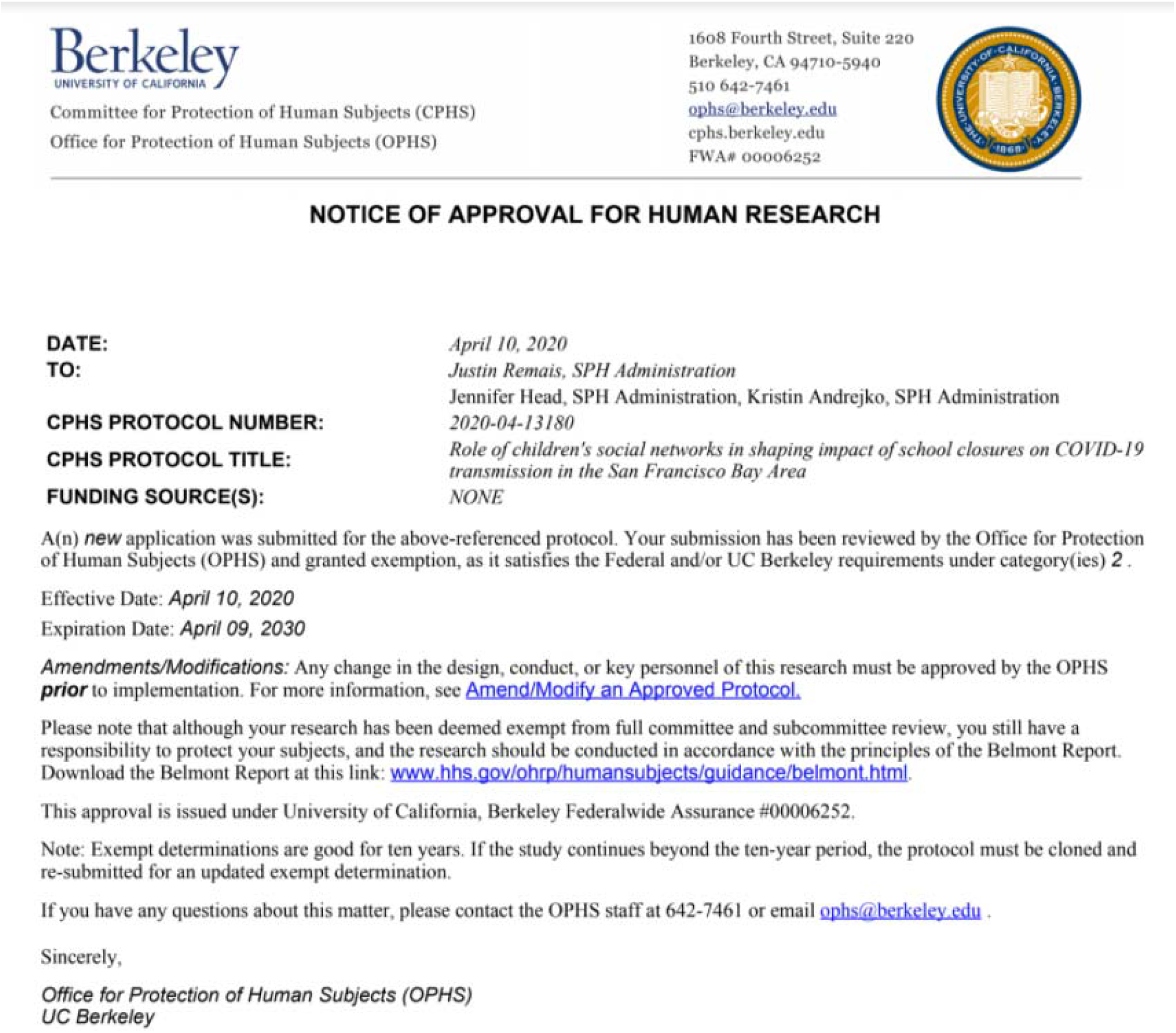

## Community Contact Survey

### Section 0: Consent

#### DESCRIPTION

You are invited to participate in a research study to understand the effect of school closures in your community. Our team is assisting public health agencies to develop computational models that understand how school closures have affected the spread of COVID-19 in your community. These models will be useful in knowing when to re-open schools and when to close schools under future outbreaks. These models depend on knowing the contact patterns of children and their families following school closures. You will be asked to fill out a form describing the number of people and their ages that you have been within 6 feet of yesterday. We ask families with children in pre-school through 12th grade to also fill out information about their children’s contacts.

#### PROJECT TEAM

We are a team of epidemiologists, mathematicians, and engineers at UC Berkeley School of Public Health who are assisting public health professionals in their COVID-19 planning and response efforts.

#### TIME COMMITMENT

Your participation will take approximately 5-10 minutes to provide information about your own contact history, and about 5 additional minutes per child to provide information about the contact history of your children.

#### RISKS AND BENEFITS

We foresee no risks associated with this study. The benefits which may reasonably be expected to result from this study are better epidemiological models that lead to more informed school closure policies. We cannot and do not guarantee that you will receive any benefits from this study.

#### PAYMENTS

This is a volunteer effort; no payments are involved. Thank you for your time!

#### PARTICIPANT’S RIGHTS

Your participation is voluntary and you have the right to withdraw your consent or discontinue participation at any time. The alternative is not to participate. Responses are confidential and anonymous. We do not collect personally identifying information and thus cannot identify you from your responses in the data. You have the right to refuse to answer particular questions. The results of this research study may be presented at scientific or professional meetings or published in scientific journals. Your individual privacy will be maintained in all published and written data resulting from the study.

#### CONTACT INFORMATION

Questions: If you have any questions, concerns or complaints about this research, its procedures, risks and benefits, contact the UC Berkeley PI contact, Justin Remais

Independent Contact: If you are not satisfied with how this study is being conducted, or if you have any concerns, complaints, or general questions about the research or your rights as a participant, please contact the UC Berkeley Office for Protection of Human Subjects to speak to someone independent of the research team at (510)-642-7461, or by. The study was approved by UC Berkeley’s Institutional Review Board with protocol ID 2020-04-13180.

Having read the information above, please select one of the two options below:

[] I CONSENT to take the survey

[] I DO NOT CONSENT to take the survey

### Section 1. Screening

1. Do you have at least one child grade preK-12 in your household?
  - Yes
  - No

### Section 2: Demographics

1. Choose one or more races that you identify as:
  - White
  - Black or African American
  - American Indian or Alaska Native
  - Asian
  - Native Hawaiian or Pacific Islander
  - Prefer not to say
  - Other
2. Do you identify as Hispanic, Latinx, or Spanish origin?
  - Yes
  - No
3. Information about income is very important to understand. Would you please give your best guess? Please indicate the answer that includes your entire household income between January 1, 2019 and December 31, 2019 before taxes.
  - Less than $19,999
  - $20,000 to $39,999
  - $40,000 to $59,999
  - $60,000 to $79,999
  - $80,000 to $99,999
  - $100,000 to $149,999
  - $150,000 or more
  - Prefer not to say
4. What is your zip code? [WRITE IN]
5. How did you hear about our survey?
  - My child’s school
  - Online forum (e.g. Berkeley Parents Network, Nextdoor)
  - Social Media
  - Friend
  - Local public Health Department
  - Other

### Section 3: State and County

1. In which state do you currently live?
  ▼ Drop down with US States
2. In which county do you currently live?
  ▼ Drop down with California counties, only display if State = California
3. Where do you live in [XX] County
  ▼ Drop down with PUMS districts in California, only display if State = California

### Section 4: Household Composition

1. How many people (including yourself) are in your household?

#### INCLUDE

- everyone who is living or staying at this address for more than 2 months
- anyone else staying at this address who does not have another place to stay, even if they are at this address for 1 month or less (ex. college student who has returned home due to university/ dorm closure)

#### DO <u>NOT</u> INCLUDE

- anyone who is living somewhere else for more than 2 months, such as a college student living away or someone in the Armed Forces on deployment
- 1
- 2
- 3
- 4
- 5
- 6
- More than 6

1. Please fill out the following information about your household:

2. In the past two weeks, have you, or anyone in your household, experienced a fever or dry cough?
  - Yes
  - No
  - Not sure/ prefer not to say
3. **BEFORE** COVID-19 related school closures, how many adults (18 years or older) typically spent the majority of school hours (8am - 3pm) at home? INCLUDE anyone who typically works from home, is unemployed, or retired. [WRITE IN NUMBER]
4. **AFTER** COVID-19 related school closures, how many adults (18 years or older) typically spend the majority of school hours (8am - 3pm) at home? [WRITE IN NUMBER]

|  | **Age**<br>**(years)** |
| --- | --- |
| Household member 1 (YOU!) | [WRITE IN] |
| Household member 2 | [WRITE IN] |
| Household member 3 | [WRITE IN] |
| Household member 4 | [WRITE IN] |
| Household member 5 | [WRITE IN] |
| Household member 6 | [WRITE IN] |
| Household member 7 | [WRITE IN] |

### Section 5: Adult Contact Diary

The following questions will ask about yesterday [INSERT DATE]. We know it’s hard to remember exactly what happened yesterday, but please give your best guess.

1. **Where did you spend the majority of your day yesterday, [INSERT YESTERDAY’S DATE]?**
  - In my home
  - At my place of work (if your place of work is your home during shelter in place, select ‘In my home’)
  - At someone else’s home who does not run a commercial daycare
  - At a commercial daycare location
  - At an outdoor leisure location
  - Performing essential activities, such as grocery shopping, laundering clothes, or receiving health care
2. Think about people **that you do not live with** that you were **within 6 feet of for more than 5 seconds** yesterday ([YESTERDAY’S DATE]). How many of these people were <u>infants, toddlers, or pre-school aged children (0-4 years)</u> [WRITE IN]
  2a. **[IF 2 > 0]** In the boxes below, write the number of infants, toddlers, or pre-school aged children (0-4 years) that you were within 6 feet of for more than 5 seconds at each location

3. Think about people **that you do not live with** that you were **within 6 feet of for more than 5 seconds** yesterday ([YESTERDAY’S DATE]). How many of these people were <u>young children (5-12 years)</u> [WRITE IN]
  3a. **[IF 3 > 0]** In the boxes below, write the number of <u>young children (5-12 years)</u> that you were within 6 feet of for more than 5 seconds at each location

|  | Number of infants, toddlers, or pre-school aged children (0-4 years) |
| --- | --- |
| In my home | [WRITE IN] |
| At my place of work | [WRITE IN] |
| At someone else's home | [WRITE IN] |
| At a childcare center that has remained open during Shelter in Place | [WRITE IN] |
| At an outdoor leisure location | [WRITE IN] |
| Performing essential activities, such as grocery shopping, laundering clothes, or receiving health care | [WRITE IN] |
| Riding or waiting for public transit | [WRITE IN] |
| My child's school | [WRITE IN] |
| Other | [WRITE IN] |

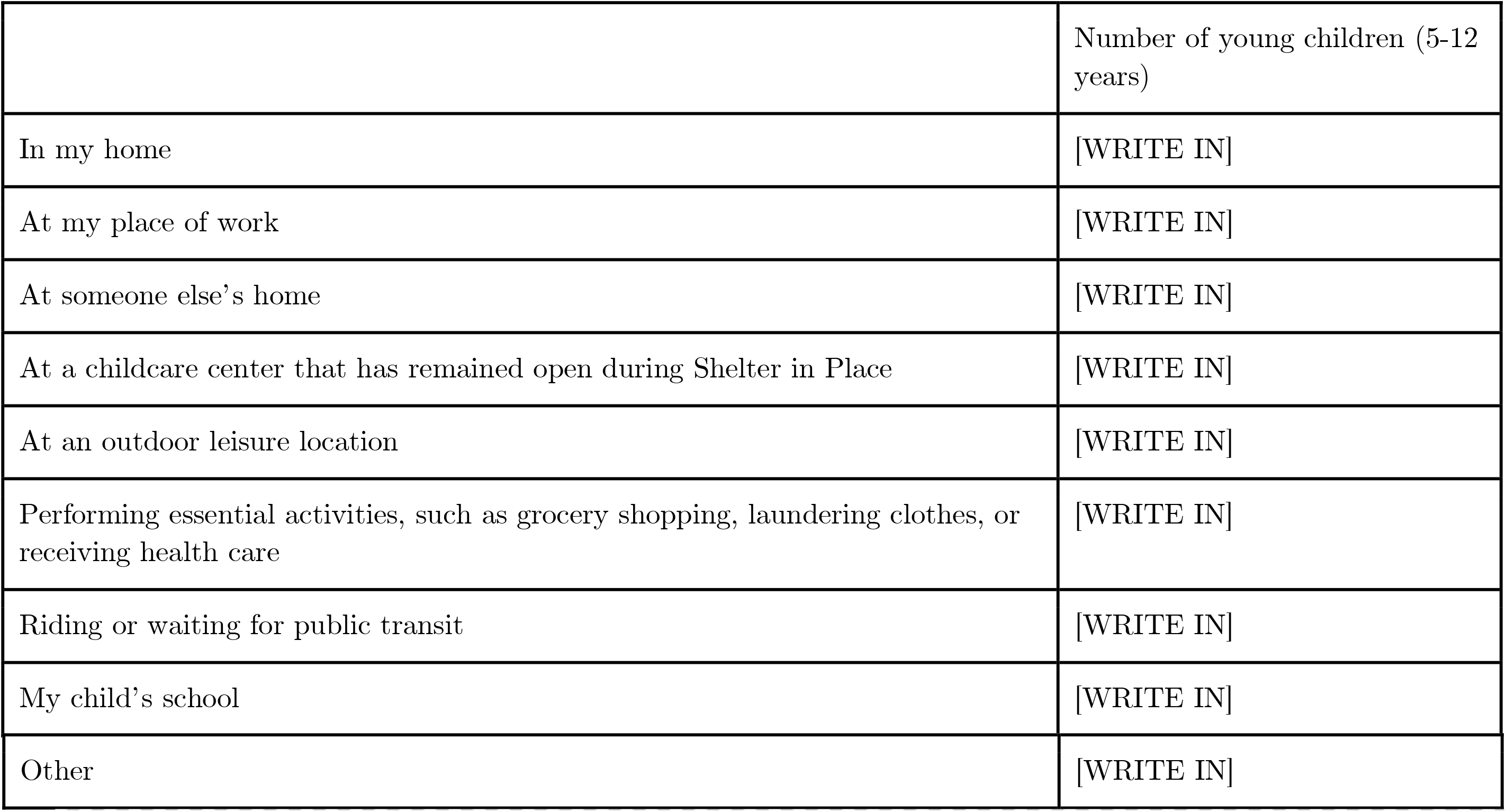 [WRITE IN]
4. Other Think about people **that you do not live with** that you were **within 6 feet of for more than 5 seconds** yesterday ([YESTERDAY’S DATE]). How many of these people <u>were teenagers (13-17 years)</u> [WRITE IN]
  4a. **[IF 4 > 0]** In the boxes below, write the number of teenagers (13-17 years) that you were within 6 feet of for more than 5 seconds at each location

5. Think about people **that you do not live with that** you were **within 6 feet of for more than 5 seconds** yesterday ([YESTERDAY’S DATE]). How many of these people were <u>young adults (18-39 years)</u> [WRITE IN]
  5a. **[IF 5 > 0]** In the boxes below, write the number of young adults (18-39 years) that you were within 6 feet of for more than 5 seconds at each location

|  | Number of teenagers (13-17 years) |
| --- | --- |
| In my home | [WRITE IN] |
| At my place of work | [WRITE IN] |
| At someone else's home | [WRITE IN] |
| At a childcare center that has remained open during Shelter in Place | [WRITE IN] |
| At an outdoor leisure location | [WRITE IN] |
| Performing essential activities, such as grocery shopping, laundering clothes, or receiving health care | [WRITE IN] |
| Riding or waiting for public transit | [WRITE IN] |
| My child's school | [WRITE IN] |
| Other | [WRITE IN] |

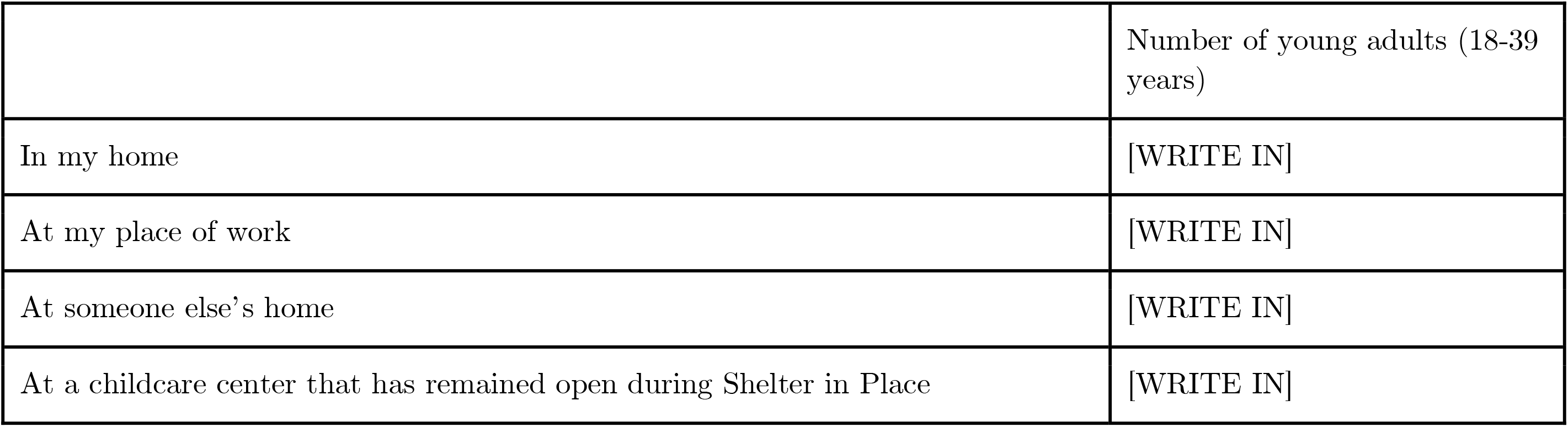
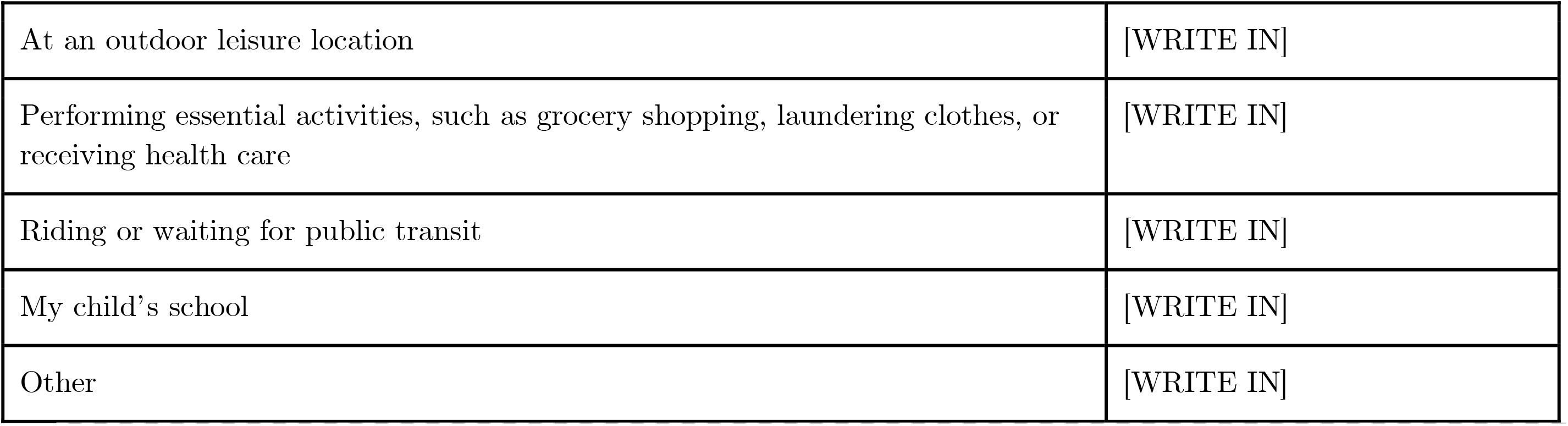
6. Think about people **that you do not live with** that you were **within 6 feet of for more than 5 seconds** yesterday ([YESTERDAY’S DATE]). How many of these people were <u>middle aged adults (40-64 years)</u> [WRITE IN]
  6a. **[IF 6 > 0]** In the boxes below, write the number of middle aged adults (40-64 years) that you were within 6 feet of for more than 5 seconds at each location

|  | Number of young adults (18-39 years) |
| --- | --- |
| In my home | [WRITE IN] |
| At my place of work | [WRITE IN] |
| At someone else's home | [WRITE IN] |
| At a childcare center that has remained open during Shelter in Place | [WRITE IN] |
| At an outdoor leisure location | [WRITE IN] |
| Performing essential activities, such as grocery shopping, laundering clothes, or receiving health care | [WRITE IN] |
| Riding or waiting for public transit | [WRITE IN] |
| My child's school | [WRITE IN] |
| Other | [WRITE IN] |

7. Think about people **that you do not live with** that you were **within 6 feet of for more than 5 seconds** yesterday ([YESTERDAY’S DATE]). How many of these people were <u>older adults (65 +)</u> [WRITE IN]
  7a. **[IF 7 > 0]** In the boxes below, write the number of older adults (65+ years) that you were within 6 feet of for more than 5 seconds at each location

|  |  |
| --- | --- |
|  | Number of middle aged adults (40-64 years) |
| In my home | [WRITE IN] |
| At my place of work | [WRITE IN] |
| At someone else's home | [WRITE IN] |
| At a childcare center that has remained open during Shelter in Place | [WRITE IN] |
| At an outdoor leisure location | [WRITE IN] |
| Performing essential activities, such as grocery shopping, laundering clothes, or receiving health care | [WRITE IN] |
| Riding or waiting for public transit | [WRITE IN] |
| My child's school | [WRITE IN] |
| Other | [WRITE IN] |

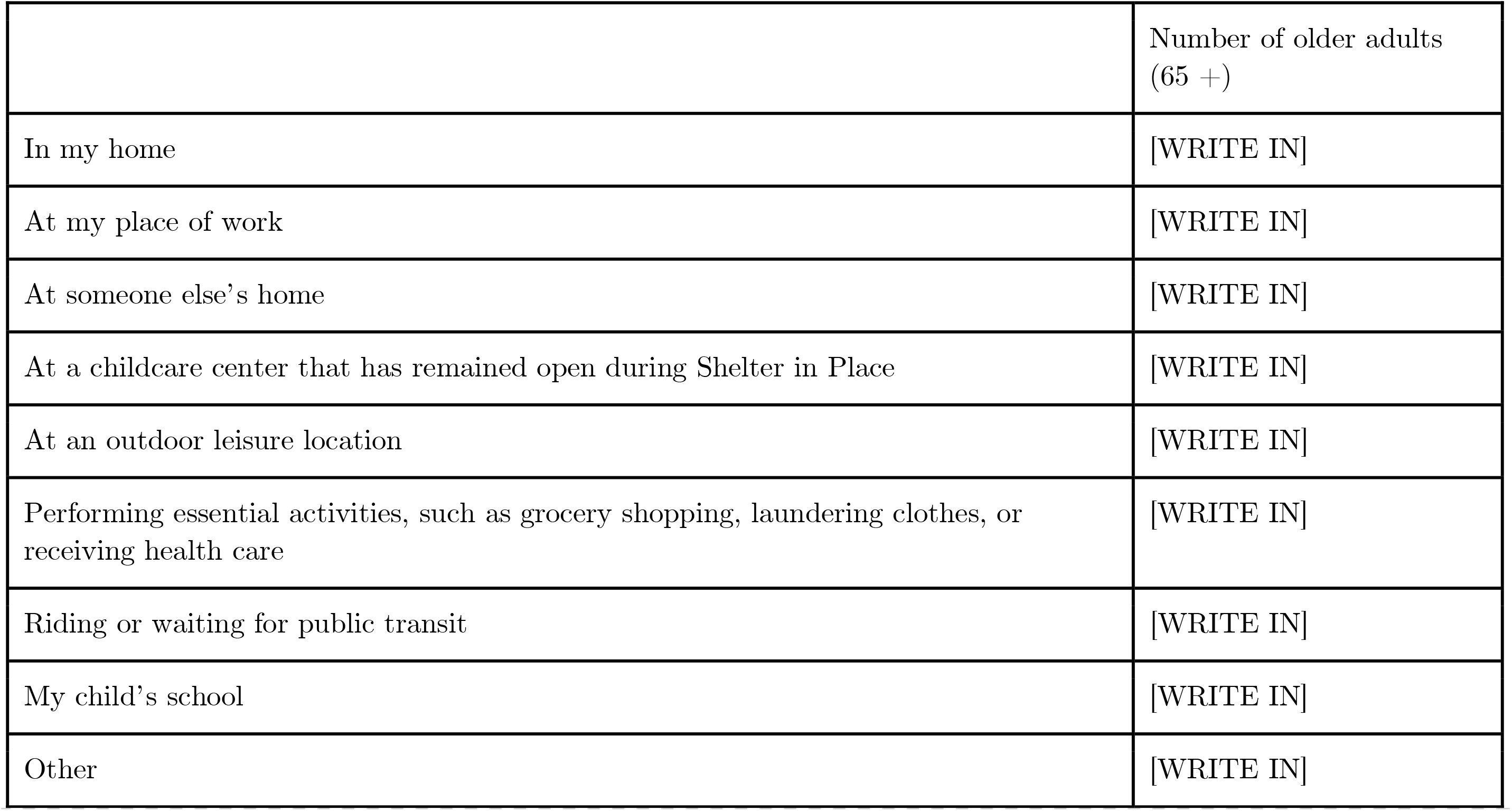
8. [IF “At my place of work” is selected for ANY of 2a. - 7a]: Where do you work?
  - Office building
  - Grocery store
  - Restaurant
  - Health care facility
  - Various locations, as a delivery driver or postal employee
  - Various locations, as a law enforcement officer
  - Construction site
  - Retail store
  - Public park
  - Gas station or garage
  - Child care/daycare center
  - School or tutoring agency
  - Food processing facility
  - Warehouse or manufacturing facility
  - Other

|  |  |
| --- | --- |
|  | Number of older adults<br>(65 +) |
| In my home | [WRITE IN] |
| At my place of work | [WRITE IN] |
| At someone else's home | [WRITE IN] |
| At a childcare center that has remained open during Shelter in Place | [WRITE IN] |
| At an outdoor leisure location | [WRITE IN] |
| Performing essential activities, such as grocery shopping, laundering clothes, or receiving health care | [WRITE IN] |
| Riding or waiting for public transit | [WRITE IN] |
| My child's school | [WRITE IN] |
| Other | [WRITE IN] |

### Section 6: Children Screening Questions

1. We are hoping to get information on all members of the household, especially children in pre-school - 12th grade. Are you willing to help by answering these questions for one or more of your children? Only answer YES if someone else in your household has not already filled out a survey for your children.
  - Yes
  - No
  - Someone else in my household has already completed the survey for my children
  - I do not have children in prek-12th grade in my household
2. *[only display the next series of questions about kids if they answer YES above; this series will display for the number of children that they selected above]*
3. How many children will you complete the survey for?
  - 1
  - 2
  - 3
  - 4
  - 5
4. Do you think school closures have helped reduce the number of covid-19 cases in your community (flatten the curve)?
  - Yes
  - No
5. Do you think school closures are necessary to flatten the curve?
  - Yes
  - No
6. Has your child missed any routine pediatrician appointments during the Shelter in Place order (ex. well-child check ups, yearly physical, routine childhood immunizations), either because you were unable to or unwilling to attend?
  - Yes-I was unable to attend a visit
  - Yes-I was unwilling to attend a visit
  - No-My child has not missed any appointments
  - No-My child has not had any pediatrician visits scheduled, but if they did, I would be willing to attend
  - No-My child has not had any pediatrician visits scheduled, but if they did, I would be unwilling to attend

### Section 7: Children Contact Diary

Please answer these questions for the [first/second/third/fourth/fifth] of your school aged children.

1. How old is your child (in years?) [Write in]
2. What type of school does your child attend?
  - Private
  - Public
  - Charter
  - Home-school
  - Other
3. [write in]
4. Where did your child spend the majority of your day yesterday, [INSERT YESTERDAY’S DATE]?
  - In my home
  - At my place of work (if your place of work is your home, select ‘In my house’)
  - At someone else’s home who does not run a commercial daycare
  - At a commercial daycare location
  - At an outdoor leisure location
  - Performing essential activities, such as grocery shopping, laundering clothes, or receiving health care
5. Think about people **that you do not live with** that your child was **within 6 feet of for more than 5 seconds** yesterday ([YESTERDAY’S DATE]). How many of these people were <u>infants, toddlers, or pre-school aged children (0-4 years)</u> [WRITE IN]
  a. **[IF 4> 0]** In the boxes below, write the number of infants, toddlers, or pre-school aged children (0-4 years) that your child was **within 6 feet of for more than 5 seconds** at each location

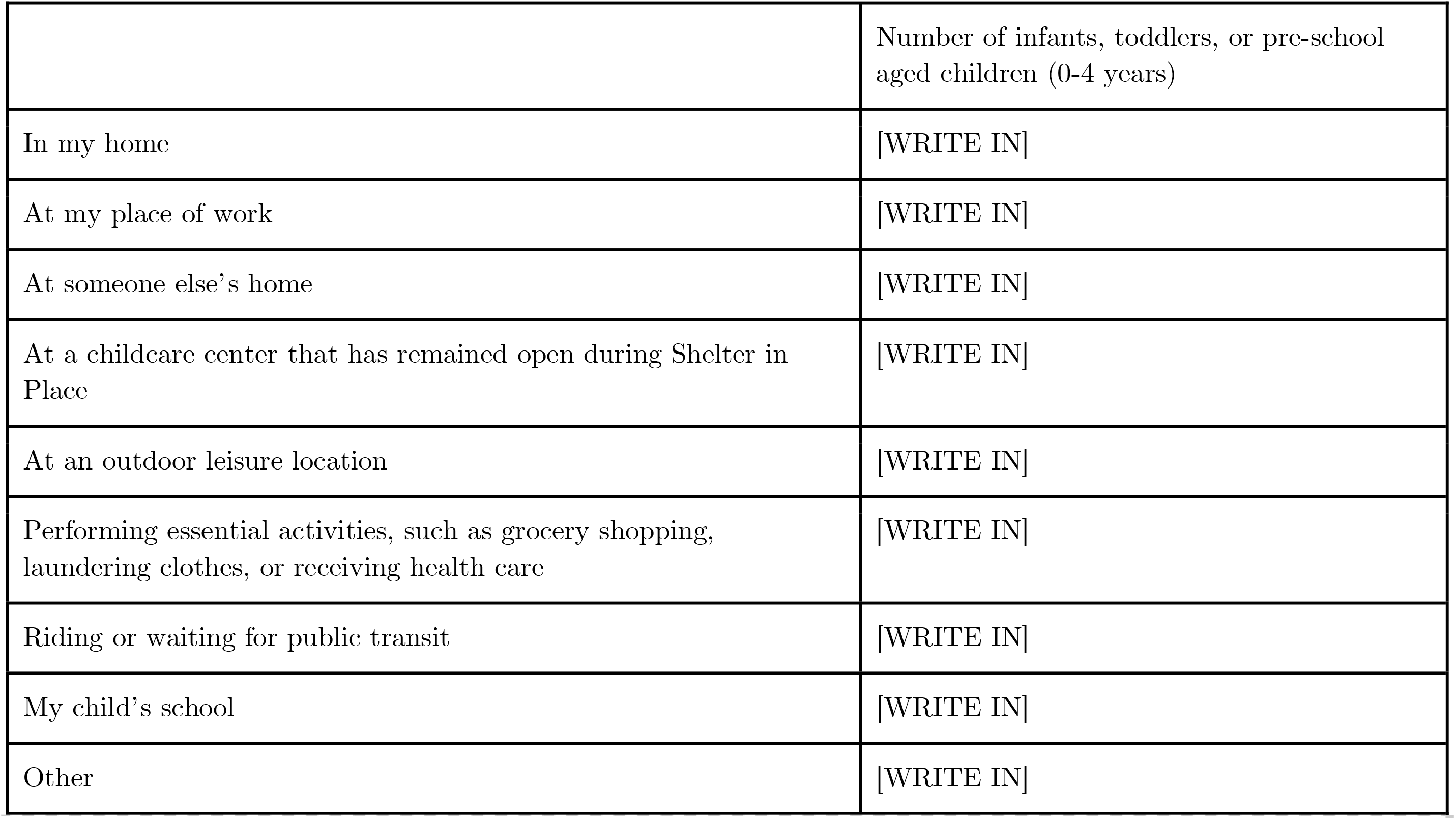
6. Think about people **that you do not live with** that your child was **within 6 feet of for more than 5 seconds** yesterday ([YESTERDAY’S DATE]). How many of these people were <u>young children (5-12 years)</u> “ [WRITE IN]”
  a. **[IF 5> 0]** In the boxes below, write the number of <u>young children (5-12 years)</u> that that your child was within 6 feet of for more than 5 seconds at each location

6. Think about people **that you do not live with** that your child was **within 6 feet of for more than 5 seconds** yesterday ([YESTERDAY’S DATE]). How many of these people were <u>teenagers (13-17 years)</u> [WRITE IN]
6a. **[IF 6 > 0]** In the boxes below, write the number of teenagers (13-17 years) that that your child was **within 6 feet of for more than 5 seconds** at each location

|  | Number of young children (5-12 years) |
| --- | --- |
| In my home | [WRITE IN] |
| At my place of work | [WRITE IN] |
| At someone else's home | [WRITE IN] |
| At a childcare center that has remained open during Shelter in Place | [WRITE IN] |
| At an outdoor leisure location | [WRITE IN] |
| Performing essential activities, such as grocery shopping, laundering clothes, or receiving health care | [WRITE IN] |
| Riding or waiting for public transit | [WRITE IN] |
| My child's school | [WRITE IN] |
| Other | [WRITE IN] |

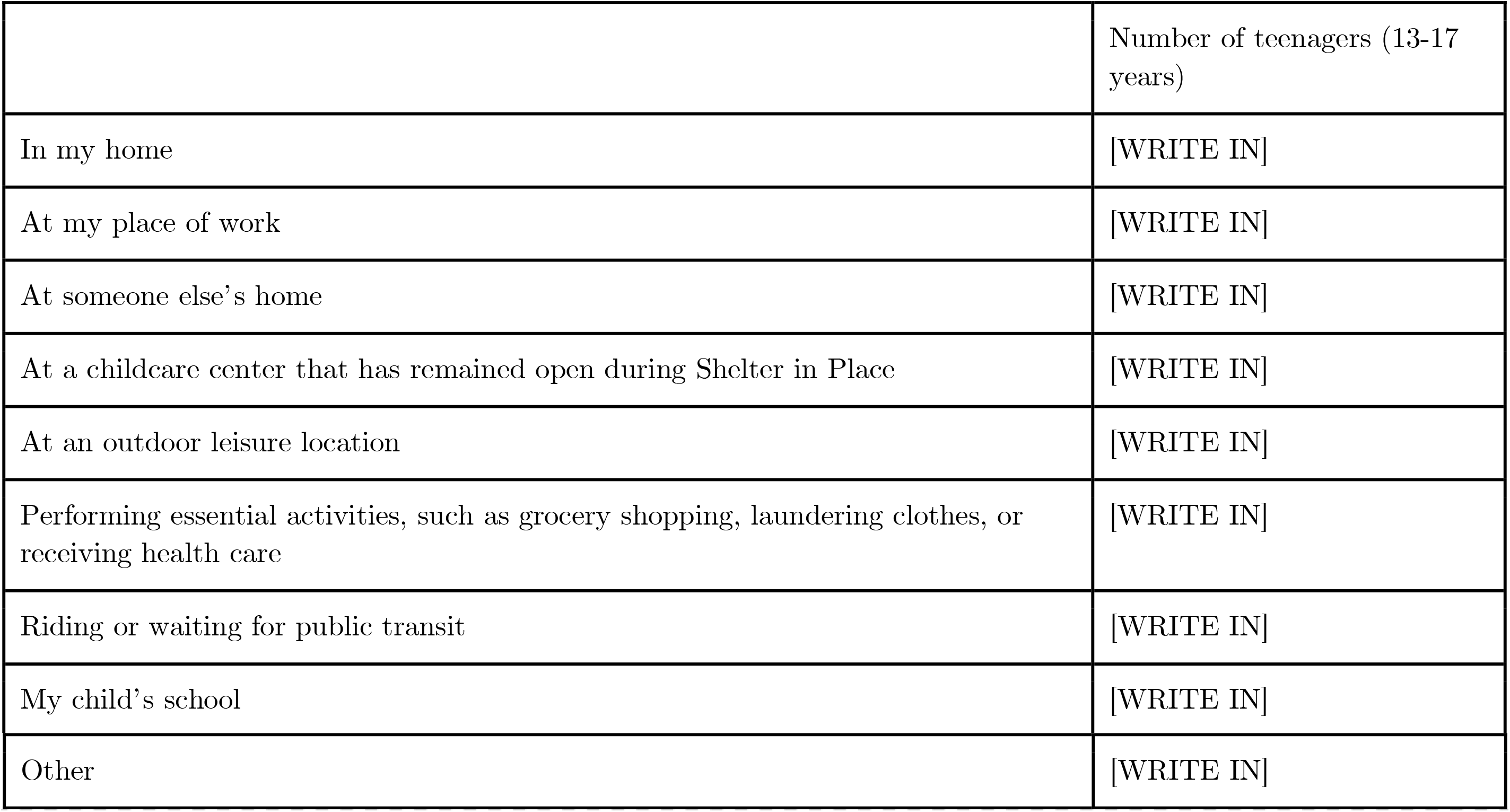 [WRITE IN]
7. Think about people **that you do not live with** that your child was **within 6 feet of for more than 5 seconds** yesterday ([YESTERDAY’S DATE]). How many of these people were <u>young adults (18-39 years)</u> [WRITE IN]
7a. **[IF 57> 0]** In the boxes below, write the number of young adults (18-39 years) that your child was within 6 feet of for more than 5 seconds at each location

8. Think about people **that you do not live with** that your child was **within 6 feet of for more than 5 seconds** yesterday ([YESTERDAY’S DATE]). How many of these people were middle <u>aged adults (40-64 years)</u> [WRITE IN]
8a. **[IF 8 > 0]** In the boxes below, write the number of middle aged adults (40-64 years) that your child was within 6 feet of for more than 5 seconds at each location

|  | Number of young adults (18-39 years) |
| --- | --- |
| In my home | [WRITE IN] |
| At my place of work | [WRITE IN] |
| At someone else's home | [WRITE IN] |
| At a childcare center that has remained open during Shelter in Place | [WRITE IN] |
| At an outdoor leisure location | [WRITE IN] |
| Performing essential activities, such as grocery shopping, laundering clothes, or receiving health care | [WRITE IN] |
| Riding or waiting for public transit | [WRITE IN] |
| My child's school | [WRITE IN] |
| Other | [WRITE IN] |

9) Think about people **that you do not live with** that your child was within 6 feet of for more than 5 seconds yesterday ([YESTERDAY’S DATE]). How many of these people were <u>older adults (65 +)</u> [WRITE IN]
9a. **[IF 9 > 0]** In the boxes below, write the number of older adults (65+ years) that your child was within 6 feet of for more than 5 seconds at each location

|  | Number of middle aged adults (40-64 years) |
| --- | --- |
| In my home | [WRITE IN] |
| At my place of work | [WRITE IN] |
| At someone else's home | [WRITE IN] |
| At a childcare center that has remained open during Shelter in Place | [WRITE IN] |
| At an outdoor leisure location | [WRITE IN] |
| Performing essential activities, such as grocery shopping, laundering clothes, or receiving health care | [WRITE IN] |
| Riding or waiting for public transit | [WRITE IN] |
| My child's school | [WRITE IN] |
| Other | [WRITE IN] |

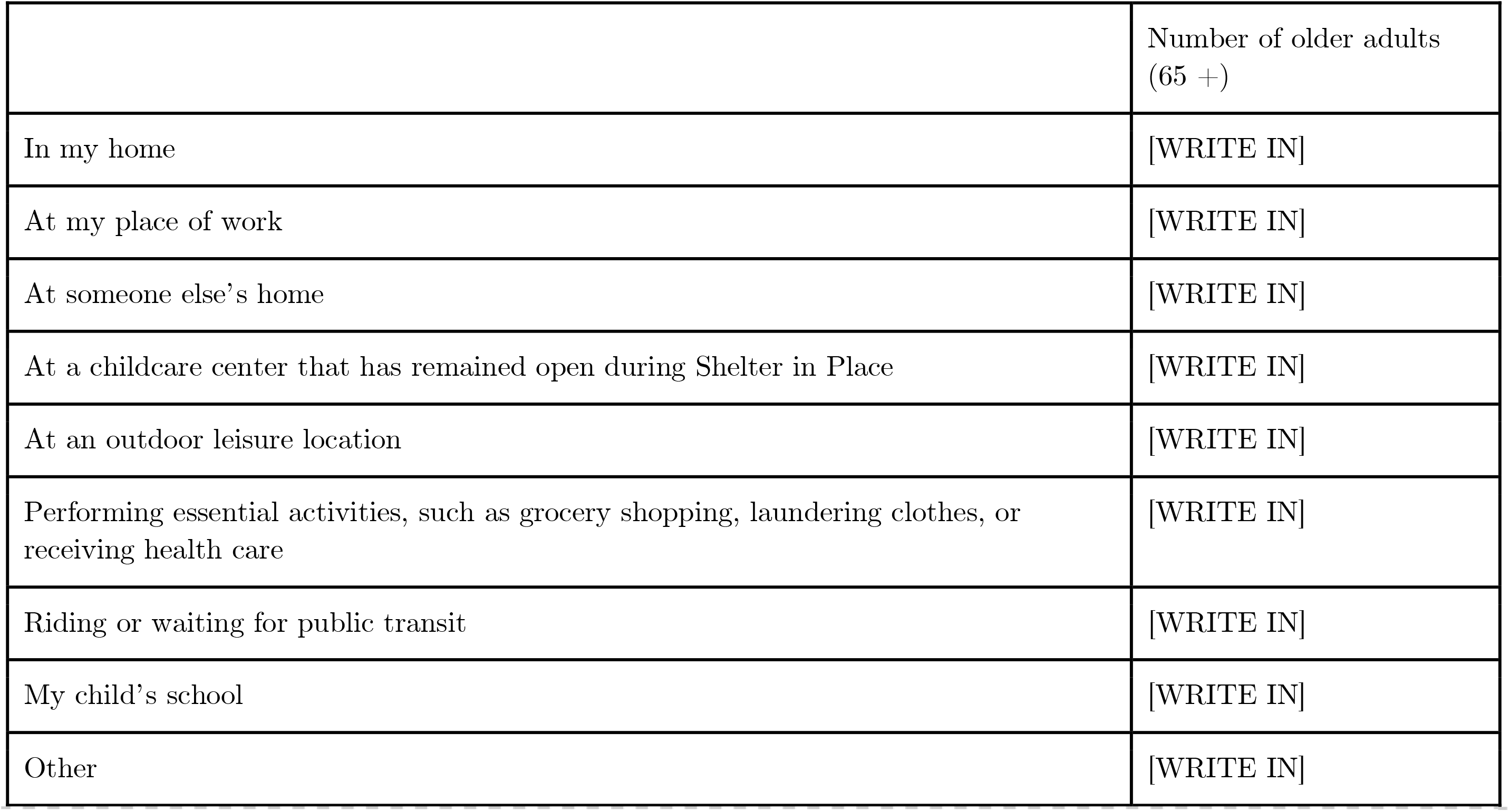 *[Repeat Q’s 1-9 in Section 7 depending on how many children said they would answer for]*

### Section 8: Thank you message

Thank you! Your response will help schools understand the impact of school closures on COVID-19 transmission in your community!

**Do you have another family member who has not taken the survey?** Please invite them to participate by sharing the link here: [custom referral link]

**Do you know other families who have not taken the survey?** Please invite them to participate by sharing the link here: [custom referral link]

#### FAQ

What will you do with this data?

Leading epidemiologists are assisting public health agencies to develop computational models that understand how school closures have affected the spread of COVID-19 in your community. These models will be useful in knowing when to re-open schools and when to close schools under future outbreaks. These models depend on knowing the contact patterns of children and their families following school closures.

We urgently need volunteers to help us understand the effect of school closures in your community.

Who is behind this project?

We are a team of epidemiologists, mathematicians, and engineers at UC Berkeley School of Public Health who are assisting federal and state officials in their COVID-19 planning and response efforts.

## Inverse probability weighting of survey responses

To account for the fact that some respondents did not indicate the locations where they had contact with a given age group, we created a linear mixed model accounting for a random effect at the household level and fixed effects for race and income to model the probability that the individual filled out a location matrix. A binary indicator of whether the individual filled out the location matrix correctly was calculated—the individual was assigned a 0 (indicated incorrect) if the respondent indicated that they have more than zero contacts in a given age category but did not indicate the location where these contacts occurred. We applied a weight defined as the inverse probability of filling out a location matrix correctly when calculating the average number of contacts per location. Weights ranged from 0 to 24 and were not truncated. Figure S2 displays the weighted contact matrices by location.

### Construction of synthetic population for transmission model

Household membership and age were drawn from a distribution based on census data on average household size (for households with and without children), proportion of households with children ages <18 years, proportion of single parent households, proportion of multi-generation households, and age of mother at first parity (Table S1). Individuals between 5-18 years old were assigned membership in a school, grade and class, using school district data on school and class sizes. Adults 18-65 years old were assigned membership in a workplace, using census data on employment, with some adults being assigned to schools (staff) and classes (teachers). College students were treated as belonging to non-essential workplaces. We validated the composition of the synthetic population by comparing household age-stratified contact patterns between our synthetic population, the 2018 one-year American Community Survey PUMS from the 9 Bay Area counties, and our household survey (Figure S1). The synthetic population had 16,000 individuals, such that each agent in the synthetic population represented 25 individuals in the real population.

### Transmission model details

We developed a discrete-time, age-structured individual-based stochastic model to simulate COVID-19 transmission dynamics in the synthetic population (Figure 1A). At each point in time, representative of one day, each individual is associated with an epidemiological state: susceptible (S), exposed (E), asymptomatic (A), symptomatic with non-severe illness (C), symptomatic with severe illness (H1, D1) resulting in eventual hospitalization before recovery (H2) or hospitalization before death (D2), recovered (R), or dead (M).

The daily contact rate between individuals *i* and *j* on day *t*, *K_ij,t_*, was estimated for pairs of individuals,

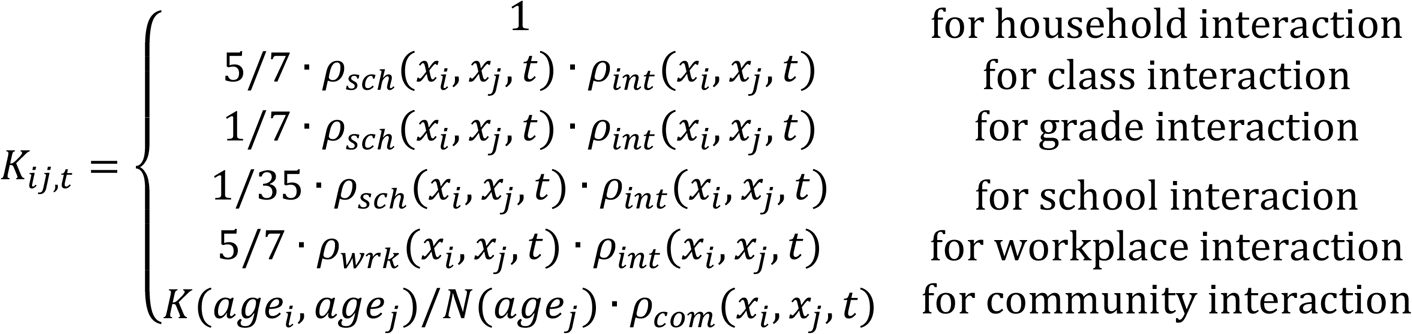

where the scaling ratios between classes, grades, and schools were obtained from previous study on transmission in various settings.^37^ Community interaction represents the number of contacts expected between individuals from age groups of individuals *i* and *j* scaled by the number of individuals in the age group of individual *j. ρ_int_(x_i_, x_j_, t)* is a factor between 0 and 1 representing a social distancing intervention to reduce contact between individual pairs, and is equal to one under a no-intervention scenario. Because symptomatic individuals mix less with the community^38^, we simulated a 100% reduction in daily school or work contacts and a 75% reduction in community contacts for a proportion (48%) of symptomatic individuals, and an additional proportion (50%) of their household members.^30^ For these individuals, *ρ_sch_*(*x_i_, x_j_, t*) and *ρ_wrk_(x_i_, x_j_, t)* is equal to 0 and *ρ_com_(x_i_, x_j_, t)* is equal to 0.25, if: 1) either individual *i* or *j* is symptomatic (C, H1, or D1) on day *t* and isolates with some probability, or 2) either individual *i* or *j* is a household member of a symptomatic individual on day *t* and quarantines with some probability; and otherwise equal to 1. We assumed that individuals were in the infectious class for up to 3 days prior to observing symptoms^39^, during which time they did not reduce their daily contacts.

Transmission was implemented probabilistically for contacts between susceptible (S) and infectious individuals in the asymptomatic (A) or symptomatic and non-hospitalized states (C, H1, D1). Movement of individual *i* on day *t* from a susceptible to exposed class is determined by a Bernoulli random draw with probability of infection per day given by the daily force of infection, λ_i,t_:

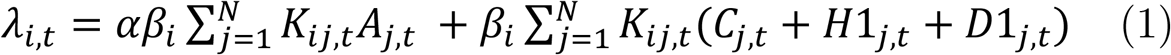

where *a* is the ratio of the force of infection between asymptomatic and symptomatic individuals; and *β_i_* is calculated from *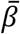*, the population mean transmission rate of the pathogen. *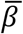* is determined using the next-generation matrix method^40^ as:

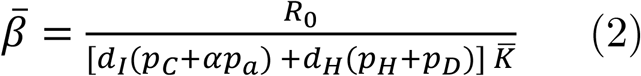

where R_0_ is the basic reproduction number (defined as the expected number of secondary cases from a single infected case in a completely susceptible population); p_s_ is the proportion of agents destined for state s; d_I_ is the average time between infection and recovery for tracks A and C; d_H_ is the average time between infection and hospitalization for tracks H and D; and *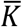* is the mean number of contacts an individual makes daily under no interventions, weighted by their probability of being contacted.^68^ We represent age-varying susceptibility^13^ using an age-stratified *β_i_* that incorporates the ratio of the susceptibility of adults to children and jointly solves equations (3) and (4):

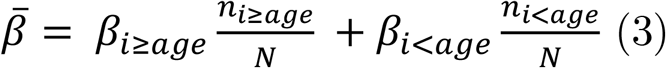

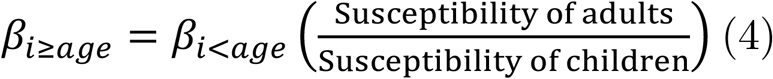

Using this method, we calculated the secondary attack rate among household members to be between 9.6% and 11.1%, in agreement with prior studies.^15,17,18,27^

The duration of the latent period, d_L_, for each individual transitioning from class E was drawn from a Weibull distribution with mean 5.4 days (95% CI: 2.4, 8.3).^42-44^ Whether an individual remained asymptomatic, or was hospitalized, or died was determined via Bernoulli random draws from age-stratified conditional probabilities (Figure 1B, Table 1). The time to recovery for non-hospitalized cases (mean: 13.1 days, 95% CI: 8.3, 16.9)^45^, the time to hospitalization for severe cases (mean: 10.3, 95% CI: 6.5, 13.3)^46^, and time to recovery or death for hospitalized cases (mean: 14.4, 95% CI: 11.3, 16.6) were sampled from Weibull distributions (Table 1).^47^ Simulations were initiated on January 17, two weeks before the first known case in Santa Clara County, assuming a fully susceptible population seeded with a random number (range: 5-10) of exposed individuals^48^. We averaged results over 1,000 independent realizations and estimated confidence intervals as the 2.5^th^ and 97.5^th^ percentile of all realizations.

## Description of reopening strategies

1. Schools open without precautions In this scenario, schools are open under a business-as-usual scenario. For all interactions, *ρ_int_*(*x_i_, x_j_, t*) = 1. The average class size is 20 students, the average sizes of elementary (K - 5), middle (6-8), and high schools (9-12) are 383, 414, and 619 students.
2. Stable cohorts: *classroom groups are enforced, reducing other grade and school contacts by 50% (weak) or 75% (strong)* In this scenario, we assume that students reduce their contacts with other teachers and students outside of their class group (or cohort) by a given proportion. We model both reductions of outside-class contacts by 50% (“weak” cohort approach) or 75% (“strong” cohort approach). The size of the class group is 20 students, on average. This may be equivalent to reductions in lunchroom or recess contacts, while still permitting chance interactions in the hallways or bathrooms. Here, we update *ρ_int_*(*x_i_, x_j_, t*) such that for the weak cohort (2a):

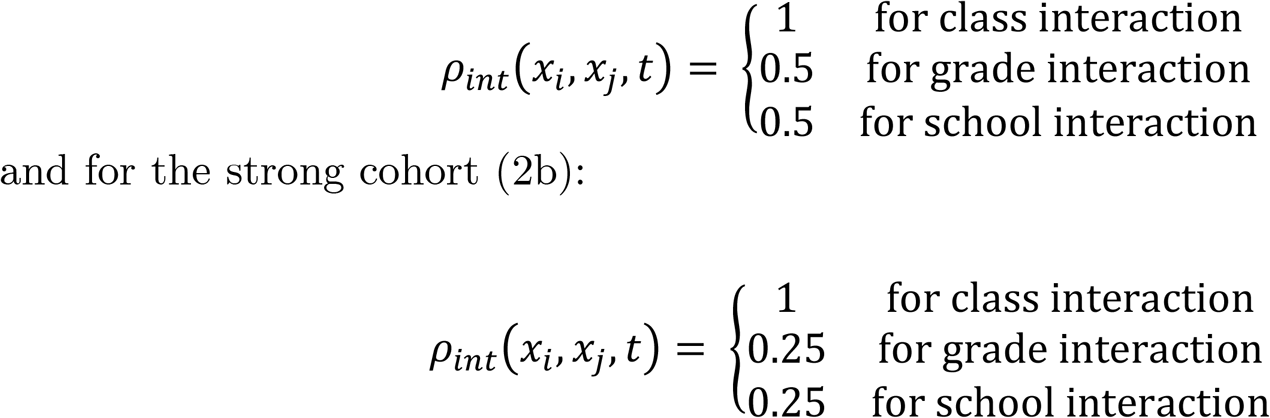
3. Staggered half classes: *Class sizes are cut in half, and each half attends two days a week* In this scenario, we assume that classes are halved, to average 10 students each. Half the class attends school two days a week, and the other half attends a different two days a week. Teachers and administrators attend four days a week. We group school, grade, and class interactions by whether or not they are within the same shift, and update *ρ_int_*(*x_i_, x_j_, t*) accordingly:for class interaction within shift, for grade interaction within shift

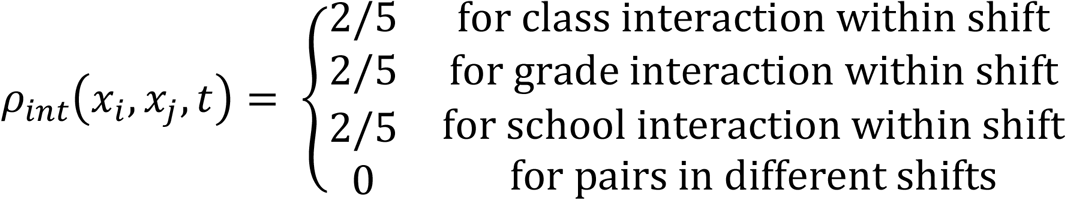
4. Staggered school days: *half the school attends two staggered days a week according to grade groups. Class size is maintained at regular levels* In this scenario, we assume that grades in a school attend two days a week, and the other half attends a different two days a week. For instance, in elementary schools, grades K-2 attend Mondays and Tuesdays, and grades 3-5 attend Thursdays and Fridays. In middle schools, grades 6-8 attend Mondayes and Tuesdays, and grade 8 attends Thursdays and Fridays. In high schools, grades 9-10 attend Mondays and Tuesdays and grade 11-12 attends Thursdays and Fridays. Teachers only attend the two days in which their classroom is present. School administrators attend all four days a week. We group school, grade, and class interactions by whether or not they are within the same shift, and update *ρ_int_*(*x_i_, x_j_, t*) accordingly:

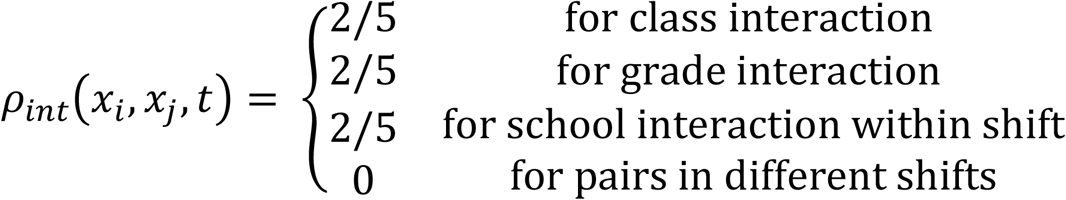
5. Students and faculty wear masks In this scenario, we assume that both students and teachers wear masks while at school. We assume that the masks both reduce the likelihood of acquiring COVID-19, as well as the likelihood of transmitting it. We assume that the effectiveness of masks for elementary school children is 15%, the effectiveness for middle school children is 25%, the effectiveness for high school children is 35% and the effectiveness for teachers is 50%. Accordingly, for each school, grade, or class pair, we have:

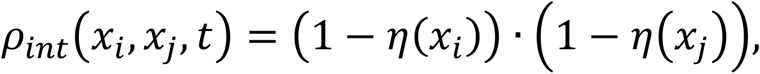

where *η*(*x_j_*) represents the effectiveness of the mask for individual *i*. such that *η*(*x_j_*) = 0.15 if the individual is an elementary school student, *η*(*x_j_*) = 0.25 if the individual is a middle school student, *η*(*x_j_*) = 0.35 if the individual is a high school student, and *η*(*x_j_*) = 0.5 if the individual is a teacher or staff member.
6. Monthly/weekly testing of teachers and students: *Faculty and students are tested with 85% sensitivity on a weekly or monthly basis^42^, and positive cases are isolated and their class quarantined for 14 days* In this scenario, every 7 or 30 days, the state of the non-hospitalized agents are ascertained through a simulated test. We assumed that the test would detect individuals in a symptomatic or asymptomatic or pre-symptomatic state with 85% sensitivity and 100% specificity. If a truly positive case was simulated to test positive, the case would reduce their school contacts by 100% for 14 days and their community contacts by 75% for 14 days. Additionally, the students or teacher in the same class as the case would reduce their school contacts by 100% and their community contacts by 75% for 14 days. This is implemented though updating *ρ_sch_*(*x_i_, x_j_, t*) and *ρ_comm_*(*x_i_, x_j_, t*) as described. If a school administrator tested positive, only the administrator isolated for 14 days.

## Choice of susceptibility parameters based on available literature

The impact of school closures depends critically on the relative susceptibility and infectiousness of children. What follows is a brief summary of key literature, emphasizing contact tracing studies where possible, as of July 17, 2020. We acknowledge that uncertainty remains in these parameters. While more studies from upper-income countries report a smaller role of children in the transmission of COVID-19 compared to adults, there is likely substantial selection bias owing to the increased likelihood of children to have less severe symptoms and the timing of studies during school closures when children had few non-household contacts. For these reasons we explore scenarios where children are half as susceptible to infection as adults, and scenarios where children are equally as susceptible to children as adults. A review by Goldstien, Lipsitch, and Cevik includes a more thorough discussion of this information.^12^

| Author | Journal | Country | Type of study | Findings |
| --- | --- | --- | --- | --- |
| Bunyavanich, et al.<sup>69</sup> | JAMA | -- | Laboratory | SARS-CoV2 uses the ACE2 receptor for host entry and the ACE2 gene expression increases linearly with age. Expression was lowest in younger children (2.40 mean log<sub>2</sub> counts per million), and increased in older children (2.77 mean log<sub>2</sub> counts per million), young adults (3.02 mean log<sub>2</sub> counts per million), and older adults (3.09 mean log<sub>2</sub> counts per million). |
| Zhang, et al.<sup>70</sup> | Preprint | China | Contact tracing | The odds of secondary attack in children was 0.34 (95% CI: 0.24 - 0.49) times that of adults. Children 0-14 years were 59% (95% CI 7-82%) less susceptible than individuals 65 years and over. |
| Mizumoto, et al.<sup>71</sup> | Preprint | Japan | Contact tracing | The odds of secondary attack in children was 0.21 (95% CI: 0.11 - 0.41) times that of adults. |
| Jing, et al.<sup>15</sup> | Preprint | China | Contact tracing | The secondary attack rate in children was 5.3%, compared to 12.6% overall; OR: 0.27 (0.13 - 0.55) |
| Li, et al.<sup>72</sup> | CID | China | Contact tracing | The secondary attack rate in children was 4%, compared to 20.5% in adults; OR = 0.16 (0.06-0.46) |
| Posfay-Barbe, et al.<sup>21</sup> | Pediatrics | Switzerland | Contact tracing | Adult household contacts had symptoms prior to or at the same time as the study child in 80% (31/39) of cases. In 8% (3/39) of households did the study child develop symptoms prior to any other household contact. 85% (75/88) of adult household contacts developed symptoms at some point, compared to 43% (10/23) of pediatric household contacts (p<0.001). |
| Danis, et al.<sup>73</sup> | CID | France | Contact tracing | A 9 year old child with co-occurring influenza and COVID 19 visited more than 80 children. Zero other students got COVID-19 but numerous other students had influenza |
| Wu, et al.<sup>74</sup> | Pediatrics | China | Contact tracing | Of 68 children with confirmed COVID-19 admitted to Qingdao Women's and Children's Hospital from January 20 to February 27, 2020, and with complete epidemiological data, 65 (95.59%) patients were household contacts of previously infected adults. |
| Dattner, et al.<sup>16</sup> | Preprint | Israel | Contact tracing | 25% of children infected over all households, compared to 44% of adults infected over all households, excluding index cases. Using a modelling approach, estimated that the susceptibility of children (under 20 years old) is 45% [40%, 55%] of the susceptibility of adults, and the infectivity of children 85% [65%, 110%] relative to adults. |
| Bi, et al.<sup>18</sup> | Lancet ID | China | Contact tracing | The household secondary attack rate in children (7.4%) was similar to that for adults (7.9%). Children were similarly susceptible to infection compared to adults, but were less likely to have severe symptoms. |
| Laxminarayan, et al.<sup>17</sup> | Preprint | India | Contact tracing | One-third of the 33,584 confirmed cases were <30 years of age in two Indian states. After adjusting for the fact that most contacts involving children occurred in the household setting, there was not strong evidence of differential risk of acquiring or transmitting infection across ages. |
| Park, et al.<sup>27</sup> | EID | South Korea | Contact tracing | A total of 11.8% (95% CI 11.2%–12.4%) household contacts of index patients had COVID-19; in households with an index patient 10–19 years of age, 18.6% (95% CI 14.0%–24.0%) of contacts had COVID-19; in households with an index patient 0-9 years of age, 5.3% (1.3–13.7) of contacts had COVID-19. |
| Fatah-Moghadam, et al.<sup>28</sup> | Pre-Print | Italy | Contact tracing | The secondary attack rate in children 0-14 was 8.4%, compared to 9.2% in adults 25-29 and >15% in adults 30 and older. Children were estimated to be the most infectious compared to other age groups. |
| Fontanet et al.<sup>33</sup> | Pre-Print | France | Retrospective cohort | Study evaluated students, teachers, staff, and family members who attended a high school linked to a cluster of COVID-19 and found an infection attack rate of 40.9% in high school affiliated students, staff, and faculty. |
| Fontanet et al.<sup>32</sup> | Pre-print | France | Retrospective cohort | Students, teachers, and staff who were exposed to SARS-CoV-2 before school closures did not develop COVID-19 following exposure to three SARS-CoV-2 positive students |
| Gudbjartsson, et al.<sup>75</sup> | NEJM | Iceland | Screening | Screened 6% of the population for COVID-19 through both targeted and random means. 13.3% and 0.8% of the target and general population were positive, respectively, and 6.3% and 0% of the targeted and general population of individuals 10 and younger. |
| Dong, et al.<sup>14</sup> | Pediatrics | China | Surveillance | 2135 pediatric patients with COVID-19 were analyzed. Children of all ages were represented in the data set as being equally susceptible, with 5.3% of cases severe. |
| Zhu, et al.<sup>76</sup> | Preprint | China, Singapore, US, Vietnam, South Korea | Systematic review of household clusters | In three (9.7%) of 31 published household clusters, the index case was a child compared to 30 (54%) of 56 published household clusters of influenza A (H5N1). |
| Davies, et al.<sup>13</sup> | Nature Medicine | China, South Korea, China, Italy, Singapore, and Canada | Modelling | SEIR models were fit to surveillance data from several countries. It is estimated that the susceptibility of children is half that of adults over 20 years old. |
| Jones et al.<sup>26</sup> | Preprint | Germany | Virology | Children and adults shed similar viral loads that likely represent infectivity. |
| Stoye <sup>77</sup> | Preprint | Germany | Virology | Re-analysis of Jones, et al. suggests that there is an increase in viral load by age, with higher viral load in older ages. |
| Lennon <sup>78</sup>, et al. | Preprint | USA | Virology | Researchers found similar distributions of viral load in patients with or without symptoms at the time of testing during the local peak of the epidemic; as the epidemic waned, individuals without symptoms at the time of testing had lower viral loads. |

**Figure S1.**
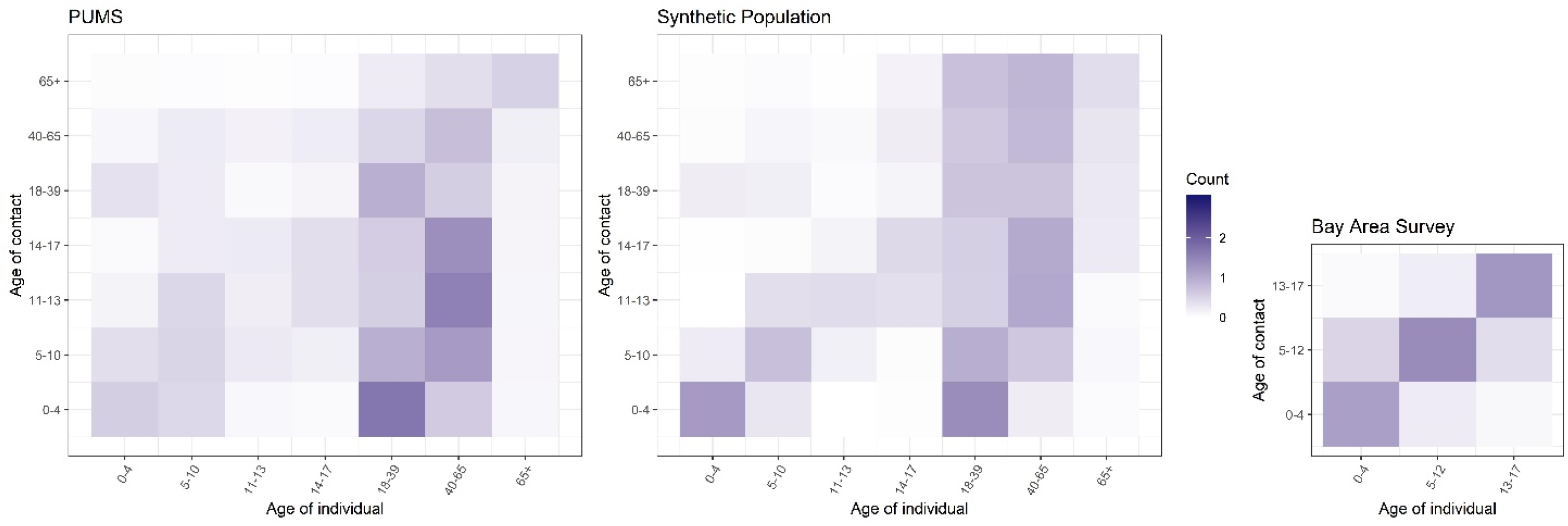
Validation of synthetic population. To validate the household composition in our synthetic population, we compared the household contact matrix for individuals represented in Public Use Microdata Sample (PUMS) from the 2018 1-year American Community Survey (left) for 9 Bay area Counties (Alameda, San Francisco, Contra Costa, Marin, Napa, San Mateo, Santa Clara, Solano, and Sonoma) and one random draw of the synthetic population (middle). Similar patterns are reflected. Compared to PUMS, the number of household contacts of the same age groups within the synthetic population are elevated, which follows the pattern seen among our household Bay Area survey (right).

**Figure S2.**
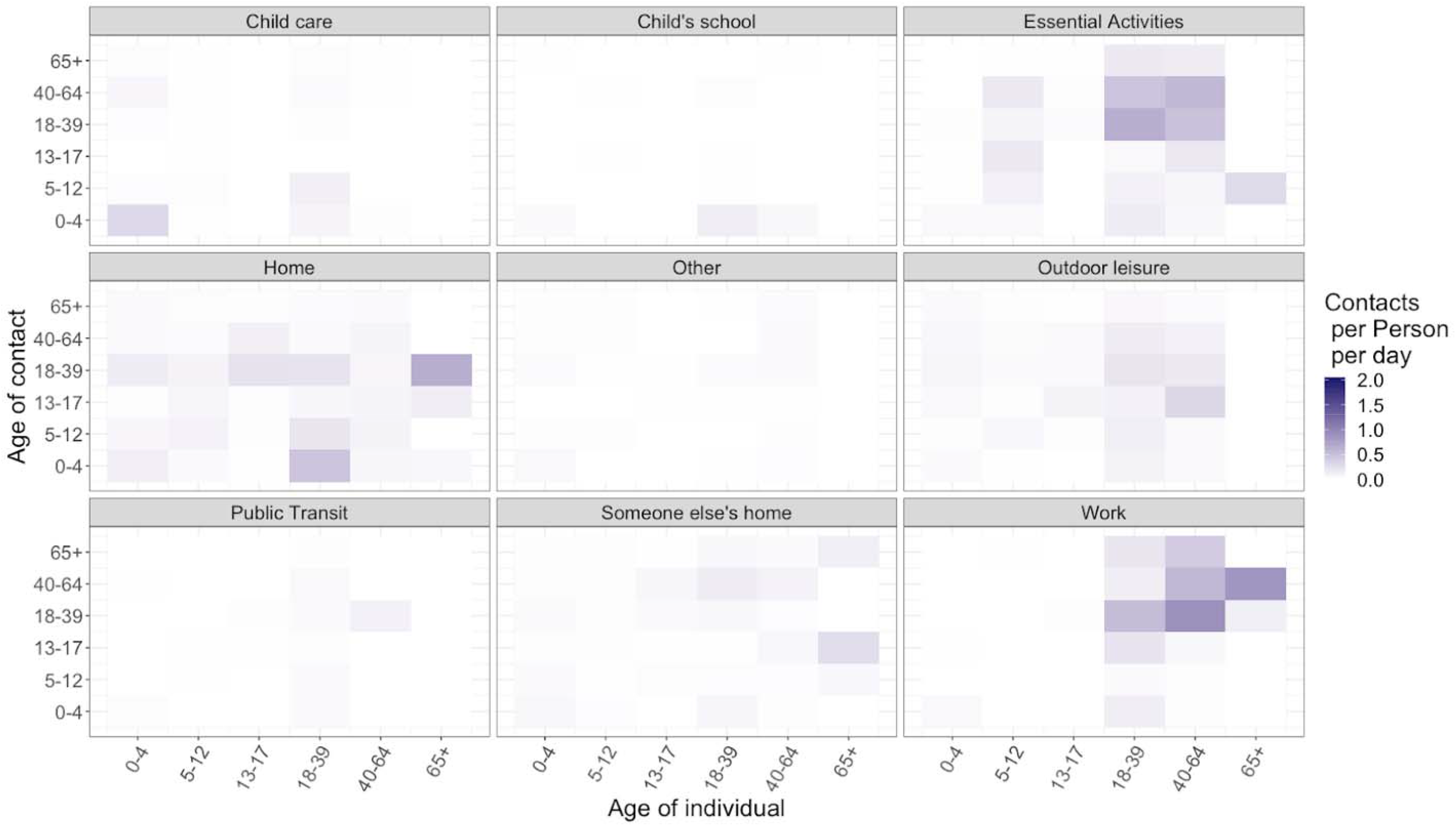
Location stratified contact matrices adjusted for non-response. We used inverse probability weighting to adjust for non-responses in location-specific contact rates. Weighted contact matrices did not differ substantially from unweighted matrices.

**Figure S3.**
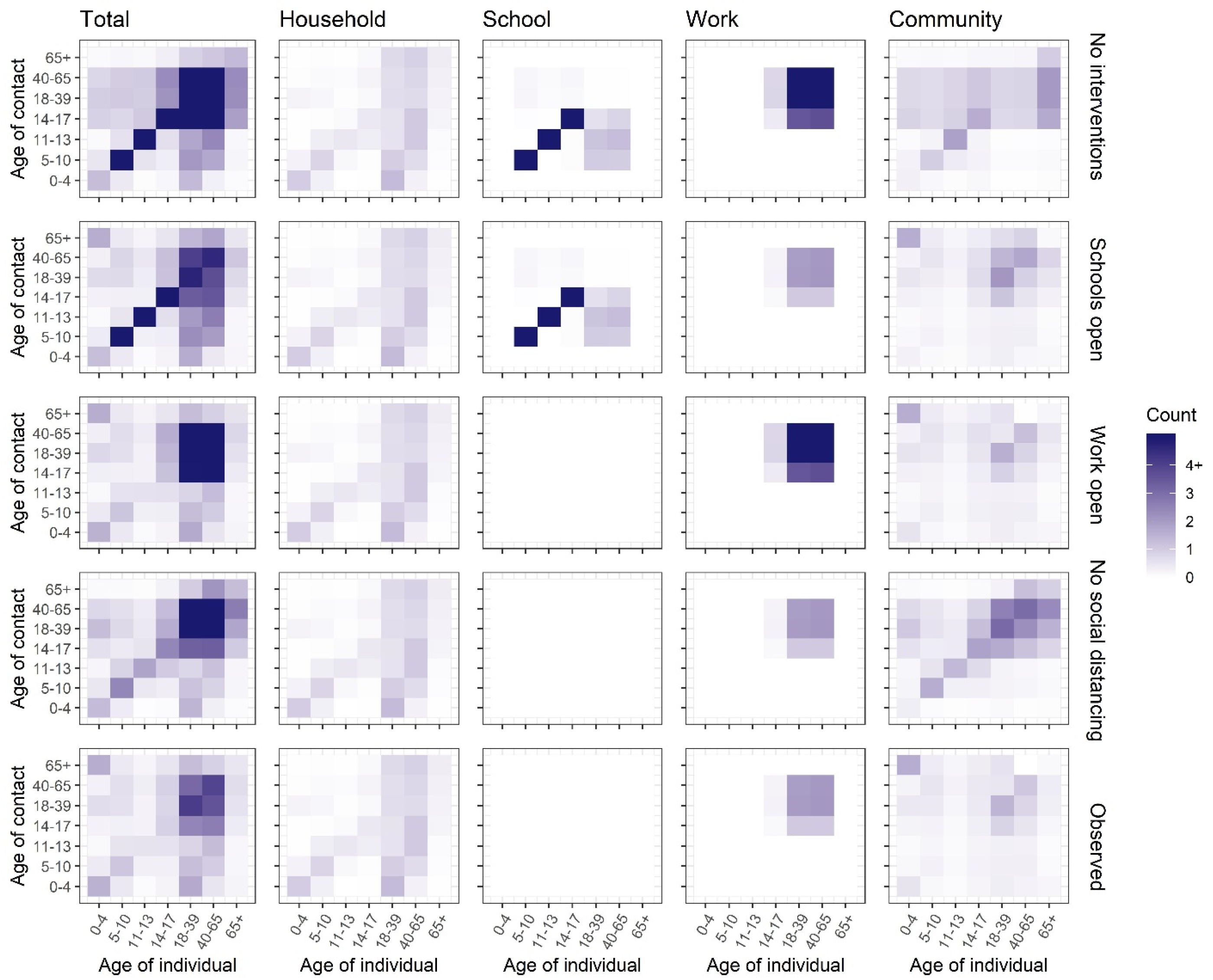
Age-specific contact matrices used for each counterfactual scenario. Synthetic age-specific contact patterns across all locations, at home, in the workplace, in school, and at other locations during normal circumstances (i.e., under no intervention) are presented in the top row. Age-specific and location-specific contact matrices under the various counterfactual physical distancing interventions are presented in rows 2-4. Observed contact patterns are presented in the bottom row. Darker color intensities indicate higher proclivity of making the age-specific contact.

**Figure S4.**
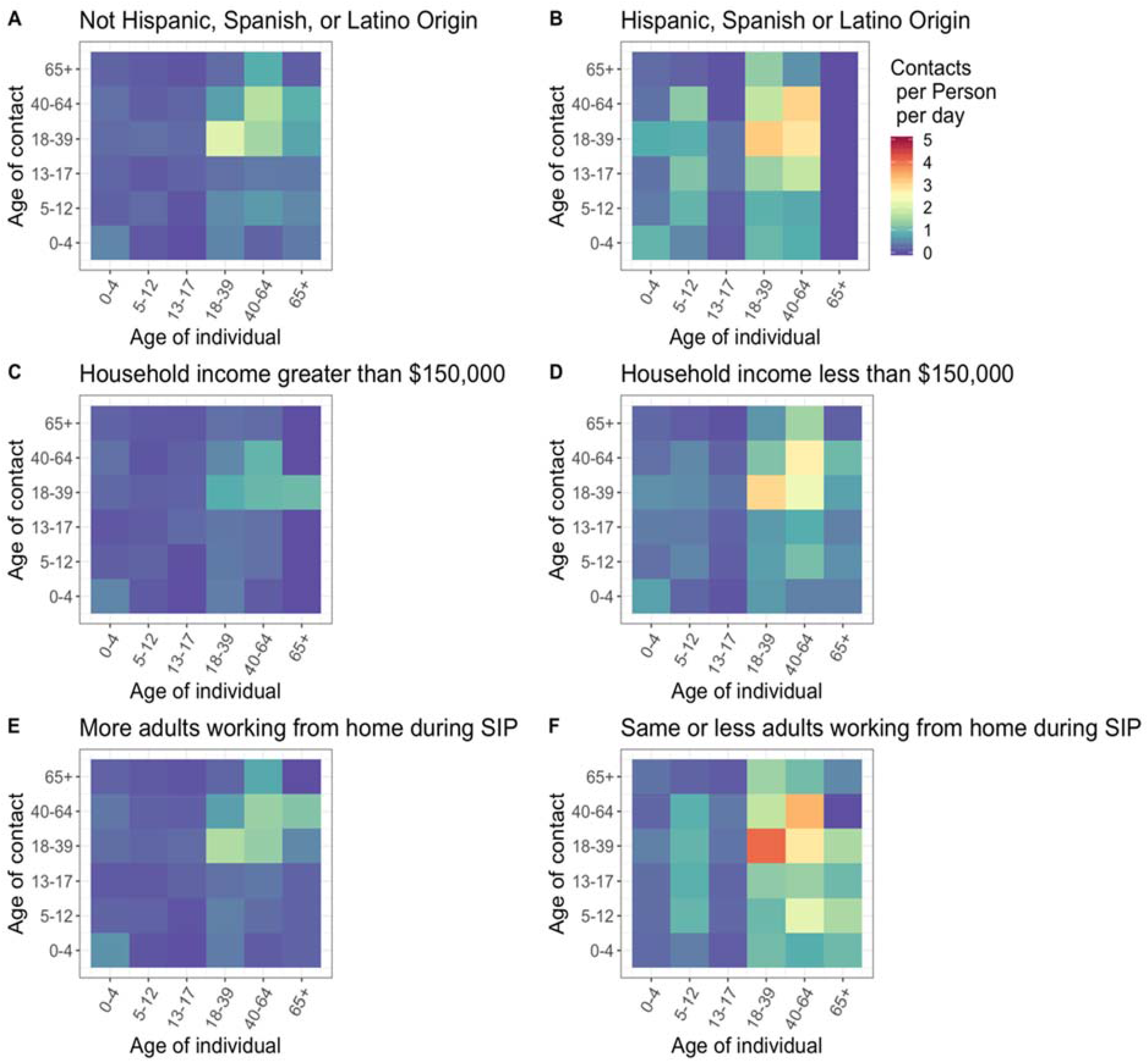
Community contact matrices by household characteristics. In multivariate adjusted regression modeling, Hispanic households had 2.32 (0.08, 4.5) more contacts than non-Hispanic households, households with an income less than $150,000 had 0.35 (−1.12, 1.8) less contacts compared to households with income less than $150,000, and households with less or the same number of adults working from home during Shelter in Place (SIP) had 1.85 (0.16, 3.52) more contacts than household with the same number or less adults working from home during SIP. SIP: shelter-in-place

**Figure S5.**
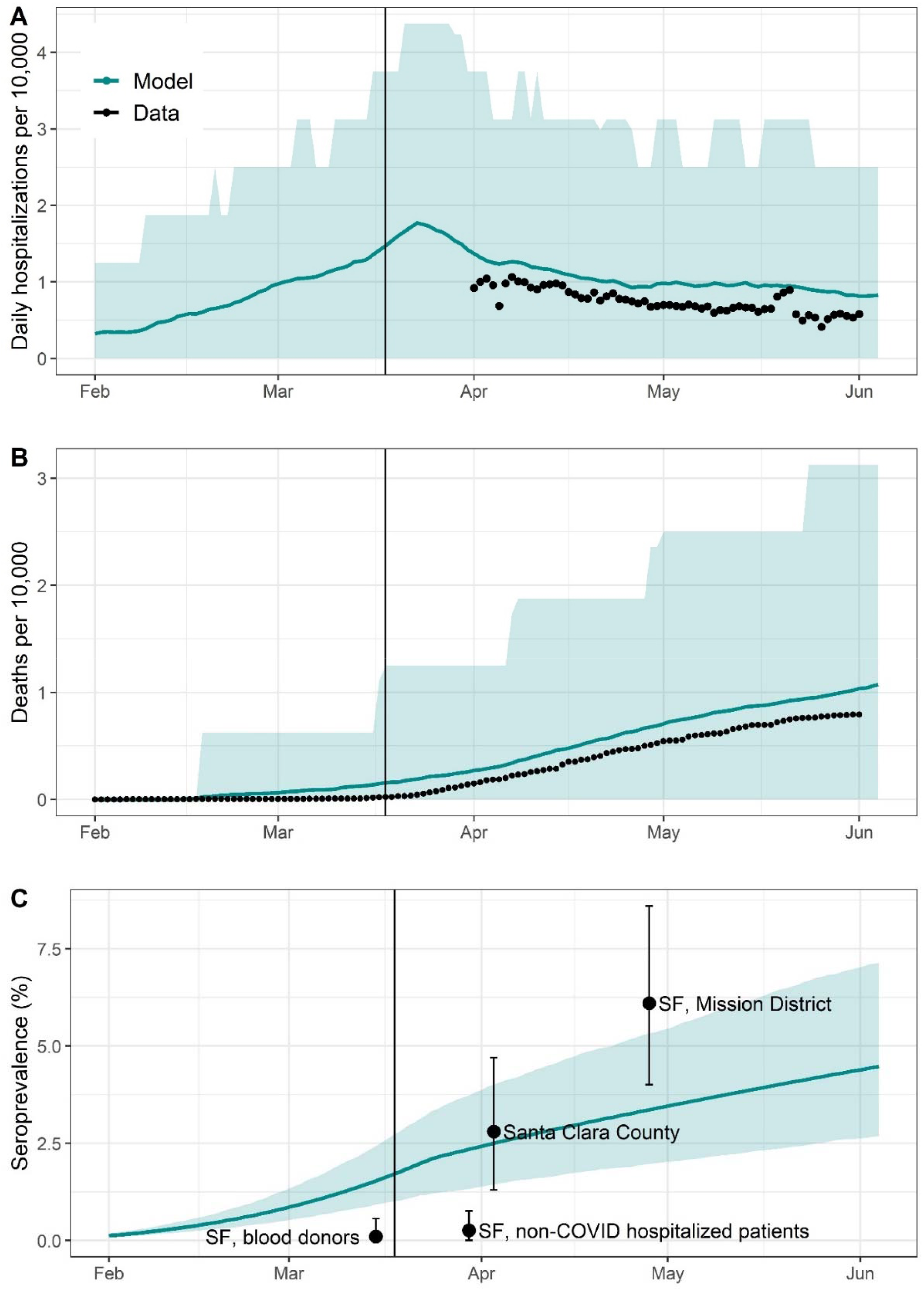
Comparison of model to observed data. Comparison of modelled (teal) to observed (black) data on daily hospitalizations per 10,000 population (A), cumulative deaths per 10,000 (B), and seroprevalence (C). The teal line represents the mean of 500 modelled simulations for the “observed” scenario, with the teal shaded region representing the 2.5th and 97.5th percentiles of model estimates. A) Data on confirmed and suspected COVID-19 hospitalizations are downloadable from the California Department of Public Health open data portal.^79^ B) Reported deaths, and population per county are available from usafacts.org.^58^ Since usafacts.org reports only confirmed deaths, we upweighted deaths by the time-varying ratio of confirmed to confirmed and suspected COVID-19 cases from ICU data. C) Estimates of the seroprevalence of infection are obtained from studies conducted in various populations from the Bay Area: blood donors from the San Francisco Bay Area, patients hospitalized at a San Francisco hospital with confirmed negative test for COVID-19,^80^ Santa Clara County,^81^ and La Mission District,^61^ a neighborhood in San Francisco with a high Latinx and essential worker population. We expect the seroprevalence of blood donors and patients hospitalized for non-COVID infections to be lower than in the source population given that blood donors tend to be healthier than the average population, and that the hospitalized population precluded capturing of current COVID-19 cases. We expect seroprevalence in La Mission District to be higher than the source population given the large proportion of essential workers in this neighborhood.

**Table S1:** Synthetic population parameters.

| Parameter | Synthetic city<br>(Oakland) | Source |
| --- | --- | --- |
| Total population | 429,082 | US Census Bureau<sup>82</sup> |
| Proportion <5 years old | 0.063 | US Census Bureau<sup>82</sup> |
| Proportion 5-17 years old | 0.135 | US Census Bureau<sup>82</sup> |
| Proportion >65 years old | 0.129 | US Census Bureau<sup>82</sup> |
| Proportion of HH with kids <18 years old | 0.251 | Bay Area Census<sup>83</sup> |
| Average size of HH with kids | 4.2 | Bay Area Census<sup>83</sup> |
| Average size of HH without kids | 2.5 | Bay Area Census<sup>83</sup> |
| Proportion of single parent HHs | 0.4 | Healthy Alameda County<sup>84</sup> |
| Proportion of HHs that are intergenerational | 0.04 | US Census Bureau<sup>85</sup> |
| Mean age of mother at first parity | 29 | New York Times<sup>86</sup> |
| Median Age | 33 | Bay Area Census<sup>83</sup> |
| Number of public elementary schools | 74 | CA Dept. of Education<sup>87</sup> |
| Number of public middle schools | 16 | CA Dept. of Education<sup>87</sup> |
| Number of public high schools | 18 | CA Dept. of Education<sup>87</sup> |
| Proportion of kids in public school | 0.87 | Ed-Data.org<sup>88</sup> |
| Average class size (elementary school) | 20 | CA Dept. of Education<sup>87</sup> |
| Average class size (middle school) | 19 | CA Dept. of Education<sup>87</sup> |
| Average class size (high school) | 19 | CA Dept. of Education<sup>87</sup> |
| Average total size (elementary school) | 383 | CA Dept. of Education<sup>87</sup> |
| Average total size (middle school) | 414 | CA Dept. of Education<sup>87</sup> |
| Average total size (high school) | 619 | CA Dept. of Education<sup>87</sup> |

**Table S2.** Representativeness of survey sample to Bay Area population. Number and proportion of individuals represented in the survey stratified by income and race sample before and after applying demographic weights. Public Use Microdata Sample (PUMS) from the 2018 1-year American Community Survey (ACMS) were used to calculate the expected distribution of income and race across for 9 Bay area Counties (Alameda, San Francisco, Contra Costa, Marin, Napa, San Mateo, Santa Clara, Solano, and Sonoma County). The expected distribution of race and income was compared against the demographic distribution of the 612 respondents in a web-based contact survey distributed across the Bay Area. Demographic weights were calculated by dividing the expected ACMS proportion by the proportion of the demographic represented in the survey sample.

|  | Un-weighted<br>N (%) | Weighted<br>N (%) | ACMS<br>N (%) |
| --- | --- | --- | --- |
| Income |  |  |  |
| Less than \$19,999 | 33 (5.4) | 27.7 (4.9) | 358345 (4.7) |
| \$20,000 to \$39,999 | 51 (8.4) | 46.3 (8.2) | 589603 (7.8) |
| \$40,000 to \$59,999 | 53 (8.7) | 50.0 (8.8) | 635775 (8.4) |
| \$60,000 to \$79,999 | 61 (10.0) | 49.8 (8.8) | 641278 (8.5) |
| \$80,000 to \$99,999 | 58 (9.5) | 50.2 (8.8) | 640969 (8.5) |
| \$100,000 to \$149,999 | 106 (17.4) | 96.2 (17) | 1392802 (18.4) |
| \$150,000 or more | 248 (40.7) | 247 (43.6) | 3124120 (41.3) |
| Race |  |  |  |
| White alone | 341 (55.9) | 286 (50.5) | 3679854 (48.6) |
| Black or African American alone | 56 (9.2) | 33.7 (5.9) | 460135 (6.1) |
| American Indian or Alaskan Native | 2 (0.3) | 0.4 (0.01) | 11094 (0.15) |
| Asian Alone | 159 (26.1) | 160 (28.2) | 2077052 (27.4) |
| Native Hawaiian or Pacific Islander Alone | 2 (0.3) | 0.39 (0.01) | 39793 (0.50) |
| Some other race alone | 23 (3.8) | 50.2 (8.9) | 811534 (10.0) |
| Two or more races | 27 (4.4) | 36.1 (6.4) | 463094 (6.1) |

**Table S3.**
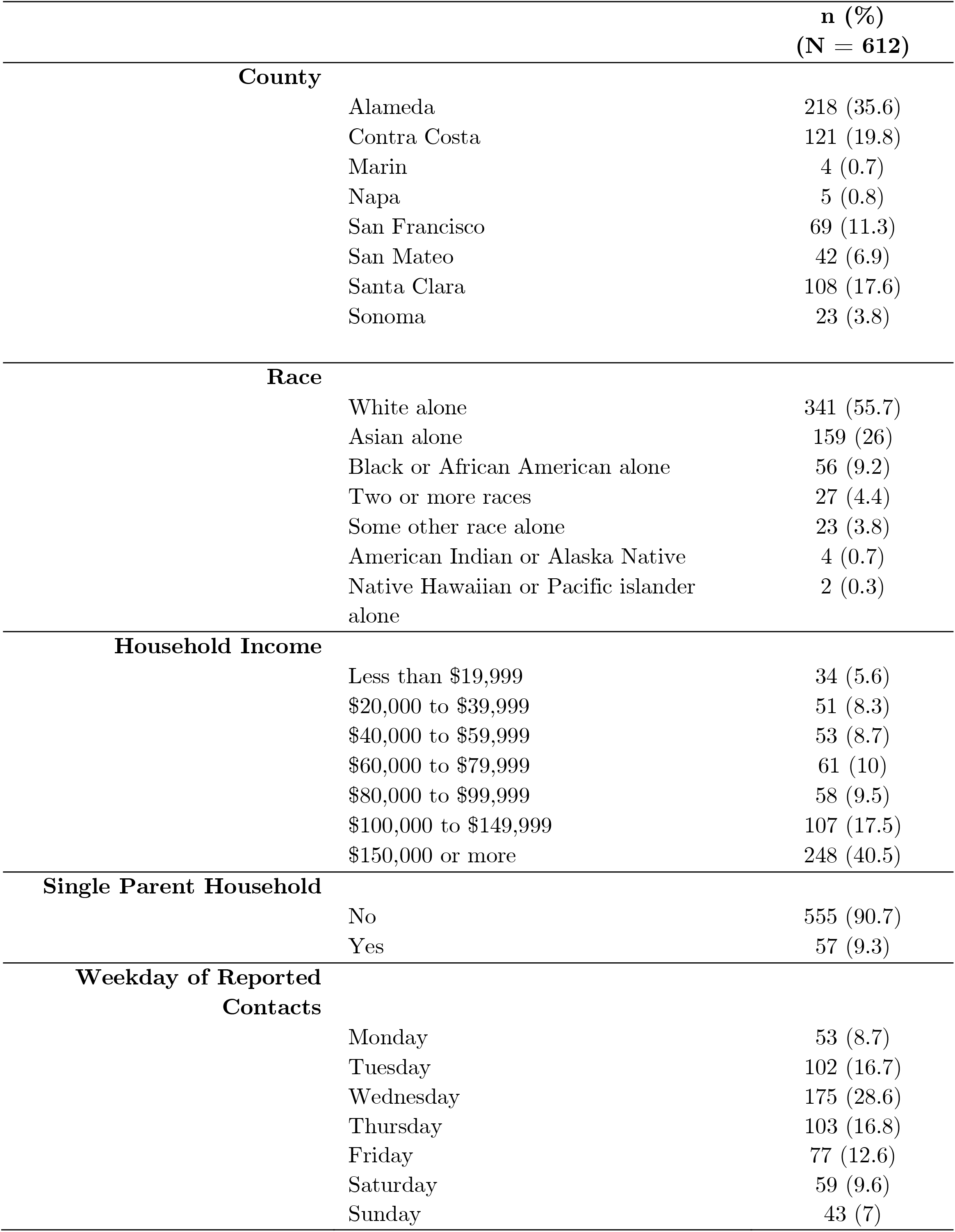
Characteristics of survey respondents. Characteristics of the 612 households who responded to the contact survey administered via Qualtrics between May 4, 2020 and June 1, 2020.

|  | n (%) |
| --- | --- |
|  | (N = 612) |
| **County** |  |
| Alameda | 218 (35.6) |
| Contra Costa | 121 (19.8) |
| Marin | 4 (0.7) |
| Napa | 5 (0.8) |
| San Francisco | 69 (11.3) |
| San Mateo | 42 (6.9) |
| Santa Clara | 108 (17.6) |
| Sonoma | 23 (3.8) |
| **Race** |  |
| White alone | 341 (55.7) |
| Asian alone | 159 (26) |
| Black or African American alone | 56 (9.2) |
| Two or more races | 27 (4.4) |
| Some other race alone | 23 (3.8) |
| American Indian or Alaska Native | 4 (0.7) |
| Native Hawaiian or Pacific islander alone | 2 (0.3) |
| **Household Income** |  |
| Less than \$19,999 | 34 (5.6) |
| \$20,000 to \$39,999 | 51 (8.3) |
| \$40,000 to \$59,999 | 53 (8.7) |
| \$60,000 to \$79,999 | 61 (10) |
| \$80,000 to \$99,999 | 58 (9.5) |
| \$100,000 to \$149,999 | 107 (17.5) |
| \$150,000 or more | 248 (40.5) |
| **Single Parent Household** |  |
| No | 555 (90.7) |
| Yes | 57 (9.3) |
| **Weekday of Reported Contacts** |  |
| Monday | 53 (8.7) |
| Tuesday | 102 (16.7) |
| Wednesday | 175 (28.6) |
| Thursday | 103 (16.8) |
| Friday | 77 (12.6) |
| Saturday | 59 (9.6) |
| Sunday | 43 (7) |

**Table S4.**
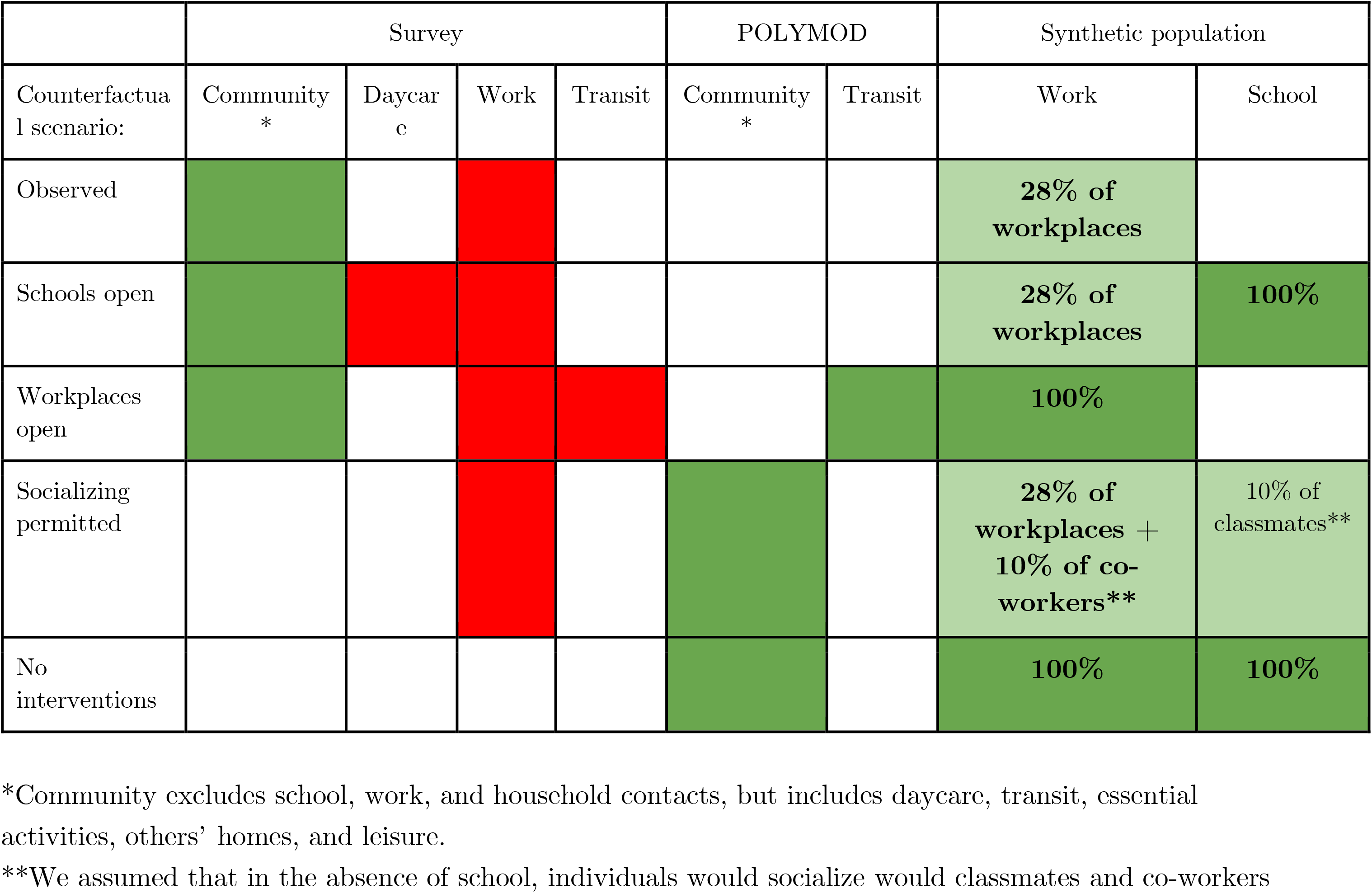
Composition of community matrices. The age-structured community matrix for analyses examining the effect of the spring semester closure (March 17 - June 1) was generated through a combination of POLYMOD and survey location data. Green boxes indicate the contact matrix was added to the overall community matrix, and red boxes indicate the contact matrix was subtracted from the overall community matrix.

**Table S5.** Effect of fall semester reopening strategies: children half as susceptible, moderate community transmission. Excess proportion of infections, symptomatic infections, hospitalizations, or deaths attributable to school reopenings, that would be experienced by each sub-group if schools were allowed to open under certain circumstances compared to if schools remained closed.

| Intervention | Subgroup | Excess percent affected, % (95% CI) |  | Excess rate per 10,000 sub-pop. (95% CI) |  |
| --- | --- | --- | --- | --- | --- |
|  |  | Infection | Symptomatic infection | Hospitalizations | Deaths |
| None | Teachers/staff (all) | 14.83 (0.93, 29.25) | 10.27 (0.47, 20.66) | 40.5 (-46.95, 146.64) | 2.97 (0, 47.17) |
|  | High sch teachers/staff | 30.5 (0, 64.17) | 20.95 (0, 46.01) | 76.53 (0, 384.62) | 4.94 (0, 181.82) |
|  | Middle sch teachers/staff | 19.53 (-2.33, 52.18) | 13.44 (-2.27, 38.65) | 55.01 (0, 444.44) | 5.04 (0, 5.04) |
|  | Elementary sch teachers/staff | 5.75 (-1.72, 15.79) | 4.09 (-1.75, 11.97) | 18.14 (-86.96, 172.45) | 1.26 (0, 1.26) |
|  | Students (all) | 14.18 (1.63, 26.77) | 2.98 (0.33, 5.83) | 0.08 (0, 0.08) | 0.01 (0, 0.01) |
|  | High school students | 25.75 (0.78, 55.99) | 5.4 (0, 12.17) | 0.17 (0, 0.17) | 0 (0, 0) |
|  | Middle school students | 17.21 (0, 46.34) | 3.62 (0, 10.11) | 0.11 (0, 0.11) | 0.03 (0, 0.03) |
|  | Elementary sch students | 4.93 (0.22, 13.48) | 1.04 (-0.1, 2.96) | 0.01 (0, 0.01) | 0.01 (0, 0.01) |
|  | HH member of students | 2.04 (-0.77, 5.07) | 1.38 (-0.51, 3.73) | 6.86 (-14.32, 30.11) | 0.87 (-3.8, 7.48) |
|  | Community member | 1.16 (-0.9, 3.28) | 0.79 (-0.7, 2.35) | 4.2 (-7.33, 16.32) | 0.54 (-2.73, 3.66) |
| Stable cohorts (weak) | Teachers/staff (all) | 3.16 (-1.42, 8.74) | 2.18 (-1.41, 6.19) | 8.46 (-47.39, 91.76) | 0.61 (0, 0.61) |
|  | High sch teachers/staff | 5.65 (-3.85, 21.06) | 3.88 (-3.7, 15.38) | 16.62 (-185.19, 196.08) | 0.2 (0, 0.2) |
|  | Middle sch teachers/staff | 3.92 (-4.65, 17.03) | 2.68 (-4.55, 13.04) | 9.27 (0, 222.22) | 0.85 (0, 0.85) |
|  | Elementary sch teachers/staff | 1.72 (-3.51, 7.69) | 1.21 (-2.7, 5.98) | 4.32 (-86.96, 87.74) | 0.71 (0, 0.71) |
|  | Students (all) | 2.92 (0.19, 6.96) | 0.61 (-0.05, 1.62) | 0.03 (0, 0.03) | 0 (0, 0) |
|  | High school students | 4.58 (-0.46, 14.09) | 0.96 (-0.17, 3.28) | 0.08 (0, 0.08) | 0 (0, 0) |
|  | Middle school students | 3.41 (-0.6, 12.89) | 0.71 (-0.4, 2.97) | 0 (0, 0) | 0 (0, 0) |
|  | Elementary sch students | 1.56 (-0.23, 4.53) | 0.34 (-0.22, 1.22) | 0.01 (0, 0.01) | 0 (0, 0) |
|  | HH member of students | 0.5 (-1.23, 2.5) | 0.35 (-0.94, 1.8) | 2.19 (-15.29, 22.34) | 0.32 (-3.83, 7.33) |
|  | Community member | 0.29 (-1.18, 1.8) | 0.2 (-0.89, 1.27) | 0.92 (-9.08, 11.86) | 0.16 (-2.75, 2.75) |
| Stable cohorts (strong) | Teachers/staff (all) | 1.25 (-2.77, 5.16) | 0.92 (-1.88, 4.25) | 2.14 (-47.39, 47.85) | 0.24 (0, 0.24) |
|  | High sch teachers/staff | 1.9 (-5.66, 10.72) | 1.39 (-3.92, 9.44) | 3.31 (-185.19, 188.68) | 0.77 (0, 0.77) |
|  | Middle sch teachers/staff | 1.65 (-6.52, 9.56) | 1.2 (-4.55, 8.7) | 2.34 (-222.22, 222.22) | 0 (0, 0) |
|  | Elementary sch teachers/staff | 0.79 (-3.51, 5.22) | 0.6 (-2.7, 4.42) | 1.59 (-86.96, 87.72) | 0.46 (0, 0.46) |
|  | Students (all) | 1.3 (0.05, 3.41) | 0.27 (-0.1, 0.81) | 0 (0, 0) | 0 (0, 0) |
|  | High school students | 1.64 (-0.65, 6.08) | 0.35 (-0.32, 1.48) | 0 (0, 0) | 0 (0, 0) |
|  | Middle school students | 1.53 (-0.65, 5.36) | 0.31 (-0.41, 1.42) | 0 (0, 0) | 0 (0, 0) |
|  | Elementary sch students | 0.95 (-0.42, 3.12) | 0.2 (-0.31, 0.88) | 0 (0, 0) | 0 (0, 0) |
|  | HH member of students | 0.22 (-1.55, 2.08) | 0.15 (-1.19, 1.53) | 0.73 (-17.97, 18.49) | 0.17 (-3.8, 3.97) |
|  | Community member | 0.15 (-1.33, 1.54) | 0.1 (-0.96, 1.14) | 0.49 (-9.94, 10.04) | 0.06 (-2.73, 2.76) |
| 2-day half class shifts | Teachers/staff (all) | 0.7 (-2.38, 3.85) | 0.49 (-2.34, 3.27) | -0.01 (-47.39, 47.85) | -0.18 (-0.18, 0) |
|  | High sch teachers/staff | 1.03 (-5.66, 9.09) | 0.63 (-5.66, 6) | -0.83 (-185.19, 172.49) | -0.49 (-0.49, 0) |
|  | Middle sch teachers/staff | 0.85 (-6.38, 8.89) | 0.54 (-4.65, 6.67) | -1.47 (-222.22, 217.39) | -0.58 (-0.58, 0) |
|  | Elementary sch teachers/staff | 0.48 (-3.6, 5.08) | 0.4 (-2.72, 3.54) | 0.9 (-87.72, 87.72) | 0.11 (0, 0.11) |
|  | Students (all) | 0.4 (-0.44, 1.31) | 0.09 (-0.19, 0.39) | 0.01 (0, 0.01) | 0 (0, 0) |
|  | High school students | 0.52 (-0.94, 2.24) | 0.12 (-0.35, 0.75) | 0 (0, 0) | 0 (0, 0) |
|  | Middle school students | 0.45 (-0.95, 2.4) | 0.09 (-0.45, 0.67) | 0.03 (0, 0.03) | 0 (0, 0) |
|  | Elementary sch students | 0.29 (-0.71, 1.3) | 0.06 (-0.32, 0.5) | 0 (0, 0) | 0 (0, 0) |
|  | HH member of students | 0.09 (-1.59, 1.8) | 0.06 (-1.14, 1.3) | -0.05 (-18.38, 18.29) | 0.06 (-3.8, 4.01) |
|  | Community member | 0.04 (-1.42, 1.55) | 0.03 (-0.96, 1.08) | -0.03 (-10.03, 9.87) | 0.01 (-2.74, 2.75) |
| 2-day staggered grades | Teachers/staff (all) | 0.68 (-2.78, 4.13) | 0.46 (-2.3, 3.24) | 2.12 (-47.62, 47.85) | -0.05 (-0.05, 0) |
|  | High sch teachers/staff | 1.12 (-5.56, 9.43) | 0.75 (-3.92, 5.88) | 5.91 (-185.19, 192.31) | 0.17 (0, 0.17) |
|  | Middle sch teachers/staff | 0.92 (-6.53, 9.09) | 0.6 (-4.65, 6.98) | 4.02 (0, 222.22) | -0.01 (-0.01, 0) |
|  | Elementary sch teachers/staff | 0.39 (-4.2, 4.43) | 0.27 (-3.42, 4.27) | -0.35 (-87.72, 87.72) | -0.17 (-0.17, 0) |
|  | Students (all) | 0.55 (-0.32, 1.66) | 0.12 (-0.19, 0.44) | 0.01 (0, 0.01) | 0 (0, 0) |
|  | High school students | 0.8 (-0.75, 3.04) | 0.18 (-0.33, 0.82) | 0 (0, 0) | 0 (0, 0) |
|  | Middle school students | 0.77 (-0.88, 3.42) | 0.17 (-0.43, 1) | 0.02 (0, 0.02) | 0 (0, 0) |
|  | Elementary sch students | 0.26 (-0.67, 1.31) | 0.05 (-0.33, 0.49) | 0 (0, 0) | 0 (0, 0) |
|  | HH member of students | 0.15 (-1.65, 1.92) | 0.1 (-1.23, 1.35) | 0.9 (-18.34, 18.7) | 0.12 (-3.79, 7.24) |
|  | Community member | 0.09 (-1.48, 1.46) | 0.06 (-0.98, 1.08) | 0.18 (-9.98, 9.96) | 0.04 (-2.76, 3.6) |
| Masks | Teachers/staff (all) | 1.73 (-2.32, 6.29) | 1.22 (-1.89, 4.76) | 4.2 (-47.39, 48.09) | 0.44 (0, 0.44) |
|  | High sch teachers/staff | 4.38 (-3.92, 18.19) | 2.94 (-3.85, 13.46) | 10.2 (-181.82, 192.31) | 0.77 (0, 0.77) |
|  | Middle sch teachers/staff | 1.84 (-6.39, 11.63) | 1.31 (-4.65, 8.89) | 3.41 (-217.51, 222.22) | -0.29 (-0.29, 0) |
|  | Elementary sch teachers/staff | 0.45 (-3.54, 5.08) | 0.38 (-3.42, 4.31) | 1.72 (-87.72, 87.74) | 0.58 (0, 0.58) |
|  | Students (all) | 2.51 (0.05, 6.95) | 0.53 (-0.05, 1.65) | 0.07 (0, 0.01) | 0 (0, 0) |
|  | High school students | 5.42 (-0.45, 18.37) | 1.13 (-0.17, 4.21) | 0.16 (0, 0.16) | 0 (0, 0) |
|  | Middle school students | 2.37 (-0.62, 9.43) | 0.5 (-0.41, 2.36) | 0.05 (0, 0.05) | 0 (0, 0) |
|  | Elementary sch students | 0.64 (-0.55, 2.2) | 0.14 (-0.31, 0.63) | 0.01 (0, 0.01) | 0 (0, 0) |
|  | HH member of students | 0.35 (-1.45, 2.34) | 0.24 (-1.15, 1.72) | 0.88 (-18.12, 18.47) | 0.19 (-3.8, 3.83) |
|  | Community member | 0.21 (-1.42, 2.01) | 0.14 (-1, 1.36) | 0.8 (-9.91, 11.01) | 0.06 (-2.73, 2.75) |
| Monthly testing of teachers & students with isolation of cases and 14 day quarantine of whole class | Teachers/staff (all) | 12.9 (0.48, 26.64) | 8.94 (0.47, 19.25) | 34.98 (-46.95, 141.53) | 3.28 (0, 47.17) |
|  | High sch teachers/staff | 27.06 (-1.79, 61.54) | 18.51 (-1.82, 44.24) | 72.19 (0, 384.8) | 5.52 (0, 185.19) |
|  | Middle sch teachers/staff | 16.61 (-2.33, 48.78) | 11.64 (-2.27, 35.57) | 46.7 (0, 434.78) | 4.12 (0, 4.12) |
|  | Elementary sch teachers/staff | 4.93 (-1.77, 14.17) | 3.48 (-1.75, 11.02) | 13.3 (-86.96, 90.09) | 1.95 (0, 1.95) |
|  | Students (all) | 12.22 (1.25, 25.17) | 2.55 (0.19, 5.51) | 0.11 (0, 0.11) | 0 (0, 0) |
|  | High school students | 22.57 (0.16, 52.71) | 4.73 (0, 11.85) | 0.25 (0, 0.25) | 0 (0, 0) |
|  | Middle school students | 14.62 (-0.2, 41.63) | 3.04 (0, 9.33) | 0.17 (0, 0.17) | 0 (0, 0) |
|  | Elementary sch students | 4.17 (0.19, 11.22) | 0.86 (-0.11, 2.57) | 0 (0, 0) | 0 (0, 0) |
|  | HH member of students | 1.76 (-0.83, 4.51) | 1.19 (-0.6, 3.17) | 6.6 (-14.82, 29.26) | 0.92 (-3.76, 7.44) |
|  | Community member | 1.01 (-0.78, 2.97) | 0.69 (-0.57, 2.08) | 3.68 (-7.27, 15.54) | 0.5 (-2.72, 3.68) |
| Stable cohorts (strong), masks, monthly testing | Teachers/staff (all) | 0.28 (-2.86, 3.32) | 0.2 (-2.39, 2.86) | 0.31 (-47.4, 47.62) | 0 (0, 0) |
|  | High sch teachers/staff | 0.47 (-5.66, 7.55) | 0.28 (-5.77, 5.88) | 2.26 (-185.19, 185.27) | 0 (0, 0) |
|  | Middle sch teachers/staff | 0.39 (-6.52, 6.82) | 0.26 (-4.65, 6.67) | -0.71 (-222.22, 212.88) | 0 (0, 0) |
|  | Elementary sch teachers/staff | 0.15 (-3.65, 4.35) | 0.13 (-3.45, 3.51) | -0.22 (-86.98, 87.72) | 0.23 (0, 0.23) |
|  | Students (all) | 0.45 (-0.38, 1.39) | 0.09 (-0.2, 0.42) | 0.02 (0, 0.02) | 0 (0, 0) |
|  | High school students | 0.68 (-0.92, 2.97) | 0.13 (-0.34, 0.82) | 0.04 (0, 0.04) | 0 (0, 0) |
|  | Middle school students | 0.56 (-0.99, 2.93) | 0.12 (-0.43, 0.83) | 0.03 (0, 0.03) | 0 (0, 0) |
|  | Elementary sch students | 0.25 (-0.65, 1.3) | 0.05 (-0.41, 0.43) | 0 (0, 0) | 0 (0, 0) |
|  | HH member of students | 0.07 (-1.7, 1.79) | 0.03 (-1.25, 1.33) | 0.17 (-15.32, 18.61) | 0.14 (-3.8, 3.82) |
|  | Community member | 0.05 (-1.4, 1.51) | 0.03 (-1.08, 1.06) | 0.37 (-10.81, 10.04) | 0.07 (-2.74, 2.77) |
| 2-day half class shifts + stable cohorts | Teachers/staff (all) | 0.24 (-2.84, 3.37) | 0.19 (-2.37, 2.84) | -0.01 (-47.39, 47.85) | 0.43 (0, 0.43) |
|  | High sch teachers/staff | 0.35 (-5.77, 7.41) | 0.13 (-5.66, 5.77) | -0.83 (-185.19, 172.49) | 0.45 (0, 0.45) |
|  | Middle sch teachers/staff | 0.46 (-6.52, 6.98) | 0.36 (-4.65, 6.67) | -1.47 (-222.22, 217.39) | -0.29 (-0.29, 0) |
|  | Elementary sch teachers/staff | 0.1 (-4.31, 4.39) | 0.14 (-2.68, 3.54) | 0.9 (-87.72, 87.72) | 0.69 (0, 0.69) |
|  | Students (all) | 0.17 (-0.47, 0.84) | 0.04 (-0.2, 0.33) | 0.01 (0, 0.01) | 0 (0, 0) |
|  | High school students | 0.2 (-0.92, 1.4) | 0.03 (-0.47, 0.53) | 0 (0, 0) | 0 (0, 0) |
|  | Middle school students | 0.21 (-1.08, 1.71) | 0.06 (-0.44, 0.64) | 0.03 (0, 0.03) | 0 (0, 0) |
|  | Elementary sch students | 0.14 (-0.71, 1.12) | 0.03 (-0.32, 0.42) | 0 (0, 0) | 0 (0, 0) |
|  | HH member of students | 0.04 (-1.65, 1.7) | 0.02 (-1.22, 1.26) | -0.05 (-18.38, 18.29) | 0.16 (-3.8, 7.26) |
|  | Community member | 0.02 (-1.41, 1.44) | 0.02 (-1, 1.03) | -0.03 (-10.03, 9.87) | -0.02 (-2.74, 2.74) |
| 2-day staggered grades + stable cohorts | Teachers/staff (all) | 0.3 (-2.83, 3.32) | 0.18 (-2.37, 2.8) | 0.93 (-47.62, 47.62) | 0.05 (0, 0.05) |
|  | High sch teachers/staff | 0.39 (-5.66, 7.41) | 0.26 (-5.45, 5.77) | 1.43 (-185.19, 188.68) | 0.56 (0, 0.56) |
|  | Middle sch teachers/staff | 0.45 (-6.52, 6.98) | 0.28 (-4.65, 4.91) | 3.36 (0, 222.22) | 0.21 (0, 0.21) |
|  | Elementary sch teachers/staff | 0.2 (-4.31, 4.35) | 0.12 (-3.48, 3.48) | -0.3 (-87.72, 86.96) | -0.26 (-0.26, 0) |
|  | Students (all) | 0.29 (-0.49, 1.05) | 0.06 (-0.23, 0.38) | 0 (0, 0) | 0 (0, 0) |
|  | High school students | 0.4 (-0.83, 2.15) | 0.09 (-0.34, 0.68) | 0.02 (0, 0.02) | 0 (0, 0) |
|  | Middle school students | 0.38 (-1, 2.01) | 0.09 (-0.44, 0.7) | 0 (0, 0) | 0 (0, 0) |
|  | Elementary sch students | 0.17 (-0.78, 1.23) | 0.03 (-0.34, 0.41) | 0 (0, 0) | 0 (0, 0) |
|  | HH member of students | 0.06 (-1.75, 1.74) | 0.04 (-1.29, 1.34) | 0.47 (-18.55, 18.74) | 0.06 (-3.82, 3.83) |
|  | Community member | 0.03 (-1.45, 1.49) | 0.02 (-1, 1.06) | -0.04 (-10.01, 9.11) | 0 (-2.75, 2.74) |

**Table S6.**
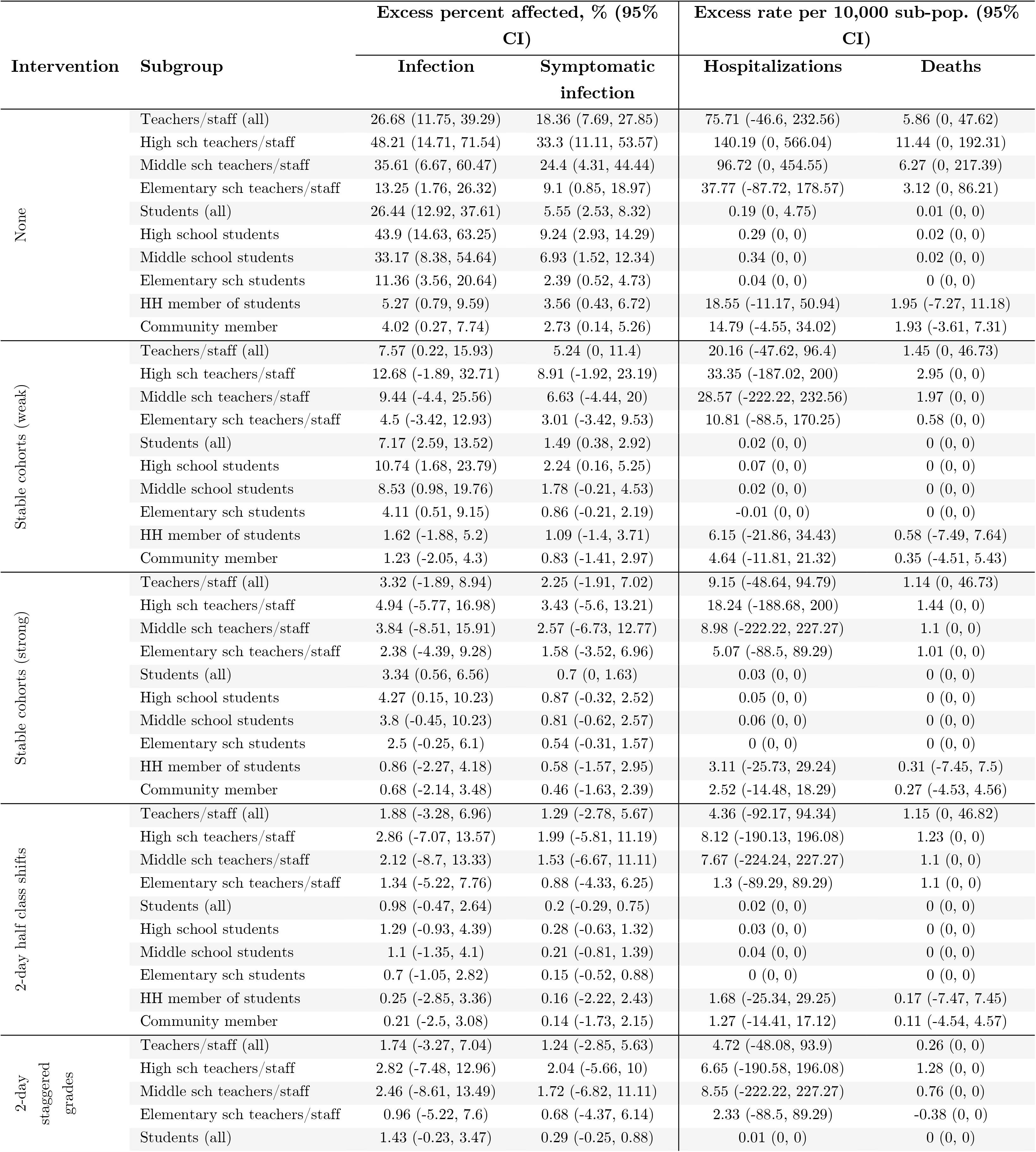
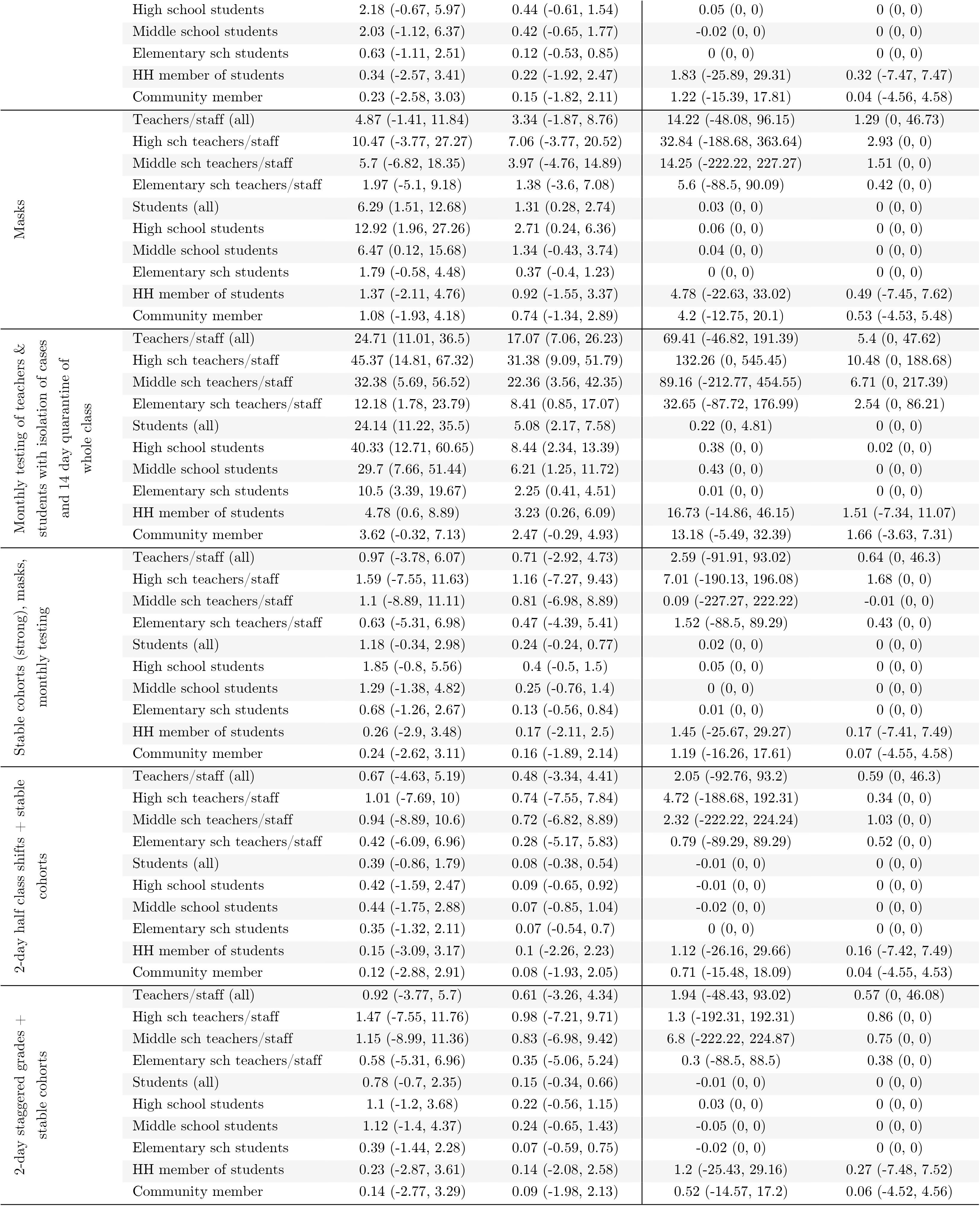
Effect of fall semester reopening strategies: children half as susceptible, high community transmission. Excess proportion of infections, symptomatic infections, hospitalizations, or deaths attributable to school re-openings, that would be experienced by each sub-group if schools were allowed to open under certain circumstances compared to if schools remained closed.

**Table S7.**
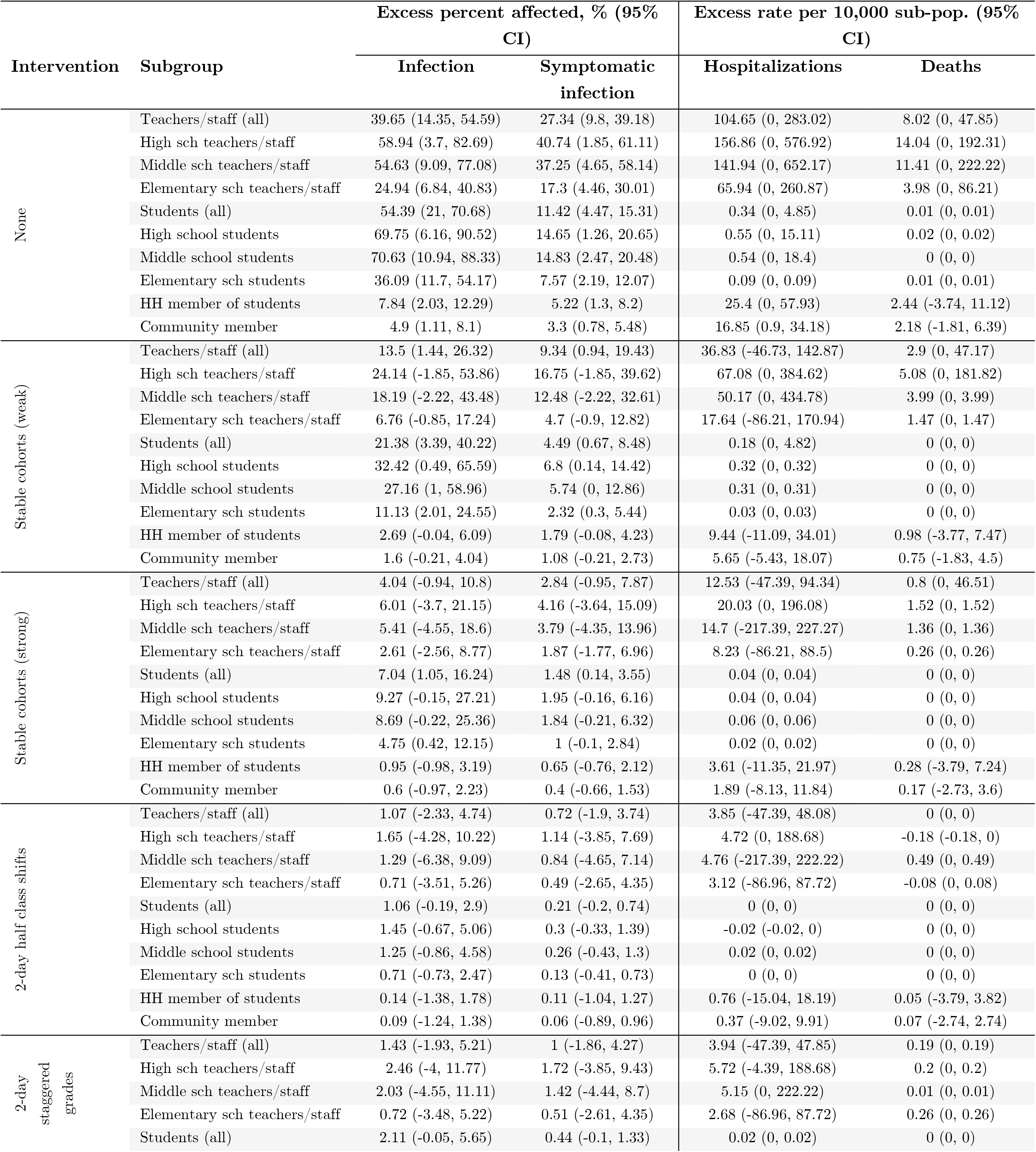
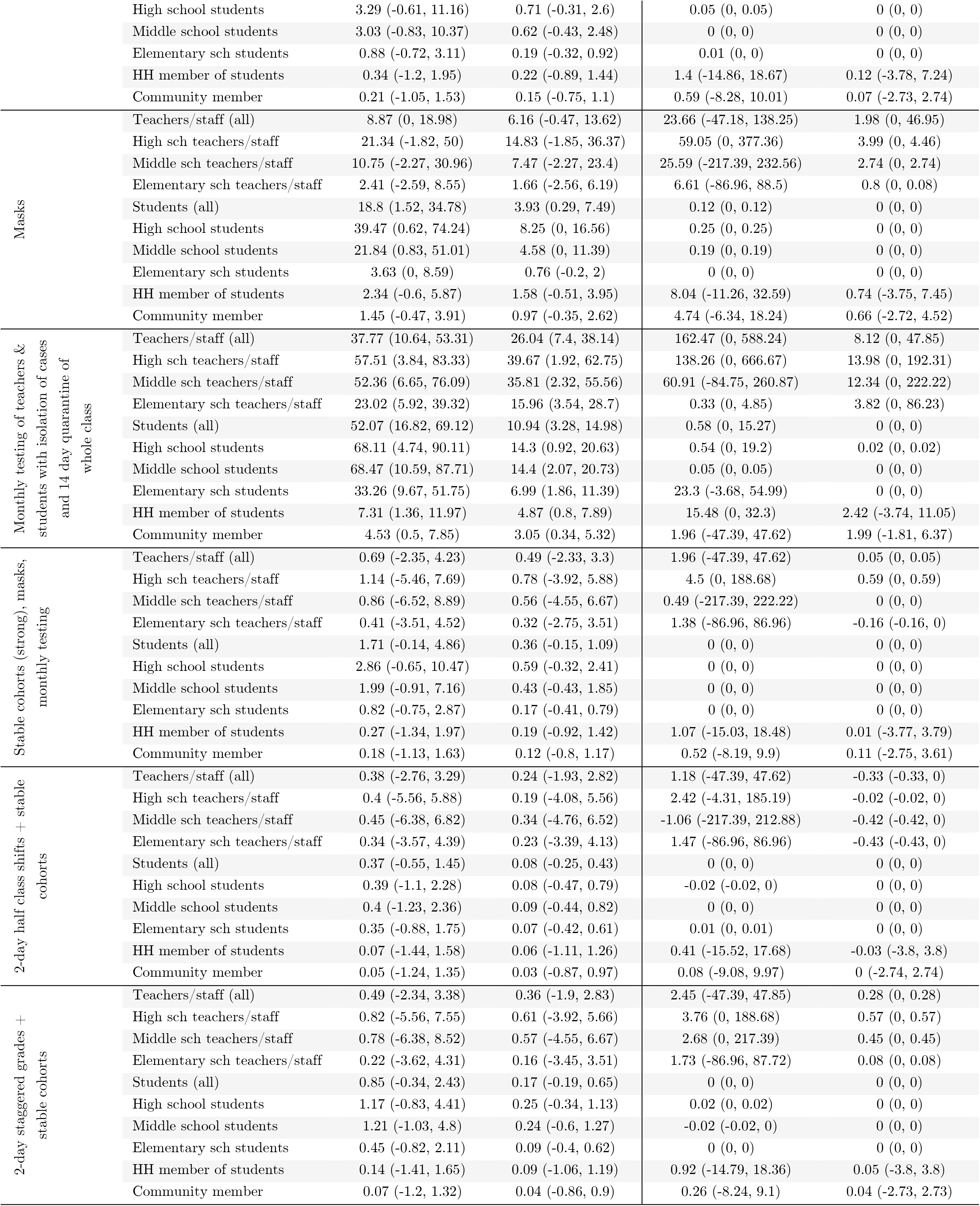
Effect of fall semester reopening strategies: children equally as susceptible, moderate community transmission. Excess proportion of infections, symptomatic infections, hospitalizations, or deaths attributable to school re-openings, that would be experienced by each sub-group if schools were allowed to open under certain circumstances compared to if schools remained closed.

**Table S8.** Effect of fall semester reopening strategies: children equally as susceptible, high community transmission. Excess proportion of infections, symptomatic infections, hospitalizations, or deaths attributable to school re-openings, that would be experienced by each sub-group if schools were allowed to open under certain circumstances compared to if schools remained closed.

| Intervention | Subgroup | Excess percent affected, % (95% CI) |  | Excess rate per 10,000 sub-pop. (95% CI) |  |
| --- | --- | --- | --- | --- | --- |
|  |  | Infection | Symptomatic infection | Hospitalizations | Deaths |
| None | Teachers/staff (all) | 46.81 (26.94, 59.43) | 32.16 (18.05, 42.59) | 119.83 (0, 285.75) | 9.39 (0, 47.85) |
|  | High sch teachers/staff | 63.32 (18, 83.93) | 43.55 (12.96, 62.51) | 159.84 (0, 576.92) | 12.38 (0, 192.31) |
|  | Middle sch teachers/staff | 59.55 (19.15, 80) | 40.85 (12.76, 58.7) | 159.64 (0, 666.67) | 11.78 (0, 222.22) |
|  | Elementary sch teachers/staff | 34.24 (17.94, 47.86) | 23.53 (11.71, 34.49) | 85.71 (-84.75, 333.47) | 7.13 (0, 87.72) |
|  | Students (all) | 61.92 (36.75, 73.92) | 13 (7.69, 16.16) | 0.37 (0, 4.86) | 0.01 (0, 0.01) |
|  | High school students | 74.17 (21.16, 90.05) | 15.62 (4, 20.55) | 0.58 (0, 15.36) | 0 (0, 0) |
|  | Middle school students | 75.05 (27.13, 88.76) | 15.68 (5.31, 20.61) | 0.59 (0, 19.27) | 0.04 (0, 0.04) |
|  | Elementary sch students | 47.25 (27.47, 60.33) | 9.92 (5.63, 13.59) | 0.11 (0, 0.11) | 0 (0, 0) |
|  | HH member of students | 12.02 (5.25, 16.85) | 7.97 (3.48, 11.33) | 38.49 (3.75, 74.46) | 3.92 (-3.75, 14.82) |
|  | Community member | 9.47 (3.93, 13.35) | 6.38 (2.78, 9.13) | 32.53 (11.71, 52.83) | 4.44 (-0.91, 10.04) |
| Stable cohorts (weak) | Teachers/staff (all) | 22.28 (6.1, 35.24) | 15.31 (3.72, 24.89) | 58.21 (-46.73, 189.57) | 3.99 (0, 47.39) |
|  | High sch teachers/staff | 34.22 (1.92, 59.26) | 23.73 (0, 44.23) | 87.61 (0, 392.16) | 5.5 (0, 185.19) |
|  | Middle sch teachers/staff | 28.88 (2.27, 52.4) | 19.88 (0, 38.31) | 76.09 (0, 454.55) | 6.11 (0, 217.39) |
|  | Elementary sch teachers/staff | 14.2 (2.7, 26.72) | 9.66 (0.87, 18.59) | 37.64 (-86.96, 178.57) | 2.51 (0, 85.47) |
|  | Students (all) | 33.75 (11.89, 49.03) | 7.08 (2.31, 10.59) | 0.25 (0, 4.83) | 0 (0, 0) |
|  | High school students | 45.21 (4.89, 70.06) | 9.43 (0.91, 15.86) | 0.48 (0, 14.62) | 0 (0, 0) |
|  | Middle school students | 42.2 (6.93, 65.67) | 8.88 (1.09, 14.76) | 0.33 (0, 0.33) | 0 (0, 0) |
|  | Elementary sch students | 21.89 (8.81, 35.15) | 4.62 (1.5, 7.64) | 0.05 (0, 0.05) | 0 (0, 0) |
|  | HH member of students | 6.21 (1.18, 10.9) | 4.12 (0.7, 7.39) | 20.15 (-7.63, 55.17) | 1.98 (-3.94, 11.14) |
|  | Community member | 4.91 (0.63, 8.84) | 3.3 (0.5, 6.01) | 16.72 (-0.92, 34.89) | 2.09 (-2.73, 7.38) |
| Stable cohorts (strong) | Teachers/staff (all) | 7.71 (0.47, 16.36) | 5.26 (-0.47, 11.59) | 19.08 (-47.62, 96.15) | 1.5 (0, 46.73) |
|  | High sch teachers/staff | 11.35 (-2, 29.42) | 7.77 (-2, 21.16) | 28.9 (-181.82, 200) | 2.07 (0, 2.07) |
|  | Middle sch teachers/staff | 9.52 (-2.27, 26.68) | 6.53 (-2.33, 20.01) | 28.26 (0, 232.56) | 1.53 (0, 1.53) |
|  | Elementary sch teachers/staff | 5.34 (-1.75, 13.56) | 3.61 (-1.77, 10.26) | 10.92 (-87.72, 90.09) | 1.21 (0, 1.21) |
|  | Students (all) | 12.65 (2.99, 23.01) | 2.64 (0.56, 5.17) | 0.07 (0, 0.07) | 0 (0, 0) |
|  | High school students | 16.38 (0.9, 37.02) | 3.41 (0, 8.13) | 0.19 (0, 0.19) | 0 (0, 0) |
|  | Middle school students | 15.36 (0.8, 34.32) | 3.19 (0, 7.79) | 0.02 (0, 0.02) | 0 (0, 0) |
|  | Elementary sch students | 8.82 (1.53, 17.24) | 1.85 (0.11, 4.09) | 0.02 (0, 0.02) | 0 (0, 0) |
|  | HH member of students | 2.23 (-0.66, 5.45) | 1.47 (-0.52, 3.67) | 7.4 (-14.8, 30.64) | 0.81 (-7.21, 7.53) |
|  | Community member | 1.75 (-0.77, 4.35) | 1.18 (-0.58, 3) | 6.06 (-7.21, 20.14) | 0.85 (-2.75, 5.42) |
| 2-day half class shifts | Teachers/staff (all) | 2.35 (-1.88, 7.11) | 1.56 (-1.9, 5.26) | 5.61 (-47.62, 93.47) | 1.03 (0, 46.08) |
|  | High sch teachers/staff | 3.57 (-5.36, 14.01) | 2.35 (-5.09, 10.72) | 7.3 (-181.82, 192.31) | 0.57 (0, 0.57) |
|  | Middle sch teachers/staff | 2.56 (-6.67, 12.77) | 1.73 (-4.88, 9.3) | 10.29 (-204.19, 227.27) | 2.2 (0, 2.2) |
|  | Elementary sch teachers/staff | 1.71 (-3.48, 7.08) | 1.13 (-3.39, 5.26) | 3.04 (-87.72, 88.5) | 0.78 (0, 0.78) |
|  | Students (all) | 1.98 (-0.19, 4.65) | 0.41 (-0.23, 1.18) | 0 (0, 0) | 0 (0, 0) |
|  | High school students | 2.63 (-0.82, 7.74) | 0.55 (-0.49, 1.96) | 0.02 (0, 0.02) | 0 (0, 0) |
|  | Middle school students | 2.32 (-1.05, 7.04) | 0.49 (-0.65, 1.92) | 0 (0, 0) | 0 (0, 0) |
|  | Elementary sch students | 1.38 (-0.86, 3.76) | 0.28 (-0.49, 1.15) | 0 (0, 0) | 0 (0, 0) |
|  | HH member of students | 0.41 (-1.87, 2.68) | 0.29 (-1.31, 1.94) | 1.47 (-18.65, 22.2) | 0.09 (-7.37, 7.38) |
|  | Community member | 0.34 (-1.8, 2.45) | 0.23 (-1.24, 1.74) | 1.2 (-10.94, 12.67) | 0.23 (-3.64, 3.66) |
| 2-day staggered grades | Teachers/staff (all) | 3.35 (-1.88, 9.26) | 2.34 (-1.88, 7.14) | 7.8 (-48.08, 94.34) | 0.85 (0, 46.73) |
|  | High sch teachers/staff | 5.92 (-5.26, 19.64) | 4.18 (-3.85, 14.81) | 15.3 (-188.68, 196.08) | 0.81 (0, 0.81) |
|  | Middle sch teachers/staff | 4.56 (-6.52, 17.78) | 3.15 (-4.65, 13.64) | 11.94 (-217.39, 232.56) | 1.56 (0, 1.56) |
|  | Elementary sch teachers/staff | 1.7 (-4.31, 8.62) | 1.19 (-3.45, 6.2) | 2.83 (-87.72, 88.5) | 0.63 (0, 0.63) |
|  | Students (all) | 4.88 (0.59, 10.04) | 1.02 (0, 2.3) | 0.05 (0, 0.05) | 0 (0, 0) |
|  | High school students | 7.33 (0, 18.82) | 1.53 (-0.18, 4.4) | 0.06 (0, 0.06) | 0 (0, 0) |
|  | Middle school students | 6.73 (-0.19, 15.63) | 1.41 (-0.42, 3.68) | 0.08 (0, 0.08) | 0 (0, 0) |
|  | Elementary sch students | 2.33 (-0.65, 5.99) | 0.48 (-0.42, 1.51) | 0.02 (0, 0.02) | 0 (0, 0) |
|  | HH member of students | 1.07 (-1.7, 3.79) | 0.72 (-1.08, 2.73) | 2.99 (-21.93, 29.39) | 0.47 (-7.34, 7.5) |
|  | Community member | 0.86 (-1.62, 3.47) | 0.58 (-1.18, 2.34) | 2.49 (-12.63, 17.28) | 0.38 (-3.67, 4.57) |
| Masks | Teachers/staff (all) | 13.58 (2.38, 24.64) | 9.33 (1.42, 17.21) | 33.77 (-47.17, 142.86) | 2.63 (0, 46.95) |
|  | High sch teachers/staff | 28.25 (1.82, 51.92) | 19.5 (0, 38.18) | 70 (0, 384.62) | 4.52 (0, 4.52) |
|  | Middle sch teachers/staff | 16.76 (0, 39.13) | 11.58 (-2.13, 28.27) | 45.05 (0, 260.2) | 1.8 (0, 1.8) |
|  | Elementary sch teachers/staff | 5.6 (-0.88, 13.68) | 3.78 (-1.71, 10.34) | 12.64 (-87.72, 91.26) | 2.08 (0, 84.75) |
|  | Students (all) | 25.95 (4.89, 40.5) | 5.43 (1.01, 8.9) | 0.17 (0, 4.75) | 0 (0, 0) |
|  | High school students | 49.56 (3.23, 76.01) | 10.39 (0.8, 16.91) | 0.33 (0, 0.33) | 0 (0, 0) |
|  | Middle school students | 31.72 (3.4, 57.43) | 6.6 (0.42, 12.84) | 0.21 (0, 0.21) | 0.02 (0, 0.02) |
|  | Elementary sch students | 7.47 (1.86, 13.7) | 1.57 (0.21, 3.33) | 0.05 (0, 0.05) | 0 (0, 0) |
|  | HH member of students | 4.51 (0.25, 8.8) | 3.01 (0.04, 5.93) | 14.86 (-11.08, 44.85) | 1.53 (-3.88, 11.04) |
|  | Community member | 3.55 (0, 7.28) | 2.39 (-0.06, 4.85) | 12.4 (-3.61, 29.2) | 1.71 (-2.7, 7.17) |
| Monthly testing of teachers & students with isolation of cases and 14 day quarantine of whole class | Teachers/staff (all) | 45.53 (24.07, 58.02) | 31.37 (15.96, 41.4) | 116.18 (0, 284.39) | 9.67 (0, 47.85) |
|  | High sch teachers/staff | 62.92 (16.98, 82.69) | 43.26 (10.9, 61.11) | 158.2 (0, 576.92) | 13.09 (0, 192.31) |
|  | Middle sch teachers/staff | 58.63 (19.13, 80.43) | 40.45 (10.64, 59.09) | 153.32 (0, 666.67) | 11.12 (0, 222.22) |
|  | Elementary sch teachers/staff | 32.42 (13.63, 47.37) | 22.35 (9.31, 34.19) | 82.27 (-84.75, 265.49) | 7.57 (0, 87.72) |
|  | Students (all) | 60.35 (31, 72.52) | 12.67 (6.98, 15.89) | 0.33 (0, 4.84) | 0 (0, 0) |
|  | High school students | 73.13 (19.49, 89.55) | 15.39 (3.76, 20.29) | 0.53 (0, 15.2) | 0 (0, 0) |
|  | Middle school students | 74.16 (28.31, 87.63) | 15.54 (5.97, 20.48) | 0.55 (0, 0.55) | 0.02 (0, 0.02) |
|  | Elementary sch students | 44.97 (22.03, 58.52) | 9.43 (4.46, 13.1) | 0.08 (0, 0.08) | 0 (0, 0) |
|  | HH member of students | 11.5 (4.09, 16.22) | 7.63 (2.68, 11.01) | 37.35 (6.97, 72.22) | 3.64 (-3.77, 14.79) |
|  | Community member | 9.02 (2.92, 12.92) | 6.08 (1.94, 8.75) | 31.08 (9.09, 50.67) | 4.14 (-0.92, 9.98) |
| Stable cohorts (strong), masks, monthly testing | Teachers/staff (all) | 1.45 (-2.36, 5.69) | 0.97 (-2.29, 4.29) | 3.74 (-47.85, 92.6) | 0.57 (0, 0.57) |
|  | High sch teachers/staff | 2.3 (-5.66, 11.11) | 1.6 (-5.46, 9.26) | 5.36 (-185.19, 192.31) | -0.19 (-0.19, 0) |
|  | Middle sch teachers/staff | 1.65 (-6.82, 11.11) | 1.19 (-6.52, 8.89) | 6.75 (-217.39, 222.35) | 0.89 (0, 0.89) |
|  | Elementary sch teachers/staff | 0.98 (-3.54, 6.14) | 0.59 (-3.45, 5.17) | 1.84 (-87.72, 88.5) | 0.78 (0, 0.78) |
|  | Students (all) | 3.18 (0.2, 7.16) | 0.67 (-0.05, 1.66) | 0.03 (0, 0.03) | 0 (0, 0) |
|  | High school students | 5.13 (-0.33, 13.71) | 1.07 (-0.32, 3.3) | 0.06 (0, 0.06) | 0 (0, 0) |
|  | Middle school students | 3.76 (-0.79, 10.31) | 0.8 (-0.45, 2.65) | 0.02 (0, 0.02) | 0 (0, 0) |
|  | Elementary sch students | 1.6 (-0.72, 4.27) | 0.35 (-0.42, 1.22) | 0.02 (0, 0.02) | 0 (0, 0) |
|  | HH member of students | 0.6 (-1.73, 2.95) | 0.39 (-1.32, 2.03) | 1.99 (-18.52, 22.91) | 0.28 (-7.34, 7.44) |
|  | Community member | 0.48 (-1.51, 2.82) | 0.33 (-1.02, 1.95) | 1.96 (-10.22, 14.52) | 0.29 (-3.61, 3.65) |
| 2-day half class shifts + stable cohorts | Teachers/staff (all) | 0.82 (-2.87, 4.74) | 0.56 (-2.76, 3.85) | 1.4 (-47.62, 48.08) | 0.51 (0, 0.51) |
|  | High sch teachers/staff | 0.96 (-5.89, 9.1) | 0.62 (-5.56, 7.55) | 0.06 (-185.19, 185.19) | -0.01 (-0.01, 0) |
|  | Middle sch teachers/staff | 0.84 (-6.98, 10.64) | 0.62 (-6.67, 8.33) | 4.7 (-217.39, 222.22) | 0.67 (0, 0.67) |
|  | Elementary sch teachers/staff | 0.75 (-4.27, 6.09) | 0.5 (-3.48, 5.17) | 0.72 (-87.72, 88.5) | 0.69 (0, 0.69) |
|  | Students (all) | 0.69 (-0.83, 2.38) | 0.14 (-0.34, 0.65) | 0 (0, 0) | 0 (0, 0) |
|  | High school students | 0.7 (-1.41, 3.33) | 0.15 (-0.62, 1.04) | 0.02 (0, 0.02) | 0 (0, 0) |
|  | Middle school students | 0.8 (-1.51, 3.76) | 0.17 (-0.68, 1.24) | 0 (0, 0) | 0 (0, 0) |
|  | Elementary sch students | 0.63 (-1.16, 2.72) | 0.12 (-0.53, 0.84) | 0 (0, 0) | 0 (0, 0) |
|  | HH member of students | 0.17 (-1.97, 2.3) | 0.11 (-1.38, 1.61) | 0.59 (-18.77, 21.36) | 0.05 (-7.34, 7.33) |
|  | Community member | 0.15 (-1.76, 2.25) | 0.1 (-1.28, 1.56) | 0.71 (-11.86, 12.72) | 0.14 (-3.64, 3.65) |
| 2-day staggered grades + stable cohorts | Teachers/staff (all) | 1.48 (-3.21, 6.25) | 1 (-2.75, 5.16) | 4.17 (-47.85, 93.47) | 0.15 (0, 0.15) |
|  | High sch teachers/staff | 2.56 (-6, 12.73) | 1.73 (-4.11, 9.44) | 7.63 (-188.68, 192.31) | -0.36 (-0.36, 0) |
|  | Middle sch teachers/staff | 2.23 (-6.82, 13.05) | 1.58 (-6.52, 10.87) | 4.41 (-217.39, 227.27) | 0.49 (0, 0.49) |
|  | Elementary sch teachers/staff | 0.69 (-5.22, 6.9) | 0.45 (-4.35, 5.31) | 2.55 (-87.72, 88.5) | 0.27 (0, 0.27) |
|  | Students (all) | 2.27 (-0.1, 5.17) | 0.46 (-0.24, 1.28) | 0 (0, 0) | 0 (0, 0) |
|  | High school students | 3.07 (-0.76, 8.43) | 0.62 (-0.5, 2.09) | 0.03 (0, 0.03) | 0 (0, 0) |
|  | Middle school students | 3.23 (-0.67, 8.99) | 0.67 (-0.61, 2.4) | -0.02 (-0.02, 0) | 0 (0, 0) |
|  | Elementary sch students | 1.25 (-1.11, 3.98) | 0.25 (-0.54, 1.16) | 0 (0, 0) | 0 (0, 0) |
|  | HH member of students | 0.53 (-2.13, 3.32) | 0.35 (-1.58, 2.26) | 1.32 (-21.93, 25.03) | 0.06 (-7.36, 7.37) |
|  | Community member | 0.41 (-2.06, 2.92) | 0.28 (-1.46, 2.02) | 1.43 (-13.44, 15.47) | 0.15 (-3.67, 4.5) |

